# Genetic Architecture of Heart Failure with Preserved versus Reduced Ejection Fraction

**DOI:** 10.1101/2021.12.01.21266829

**Authors:** Jacob Joseph, Qin Hui, Chang Liu, Krishna Aragam, Zeyuan Wang, Brian Charest, Jennifer E. Huffman, Serkalem Demissie, Luc Djousse, Juan P. Casas, J. Michael Gaziano, Kelly Cho, Peter W.F. Wilson, Lawrence S. Phillips, VA Million Veteran Program, Christopher J. O’Donnell, Yan V. Sun

## Abstract

**Background:** Pharmacologic clinical trials for heart failure (HF) with preserved ejection fraction (HFpEF) have been largely unsuccessful as compared to those for heart failure with reduced ejection fraction (HFrEF). Whether differences in the genetic underpinnings of these HF subtypes may provide insights into the disparate outcomes of these clinical trials remains unknown.

**Objectives:** We pursued genetic association analyses to compare the genetic architecture of HFpEF with that of HFrEF.

**Methods:** We created a non-Hispanic White cohort including 19,495 HFrEF and 19,589 HFpEF cases among 43,344 unclassified HF cases, and 258,943 controls without HF in the Veterans Health Administration Million Veteran Program. We then conducted genome-wide association studies of unclassified HF, HFrEF and HFpEF, followed by genetic correlation analyses and Mendelian randomization analyses of established HF risk factors with HFrEF and HFpEF.

**Results:** We found 13 loci associated with HFrEF at genome-wide significance, but only one associated with HFpEF. Among genome-wide significant loci for HFrEF, four loci were not associated with any HF risk factor. The single locus identified for HFpEF (*FTO*) is a known marker for obesity. Genetically determined associations were widely different between HFrEF and HFpEF for several risk factors including coronary artery disease, lipid levels, and pulse pressure.

**Conclusions:** The modest genetic discovery for HFpEF compared to HFrEF despite a robust sample size indicates that HFpEF, as currently defined, likely represents the amalgamation of several, distinct pathobiological entities. Development of consensus sub-phenotyping of HFpEF is paramount to better dissect the underlying genetic signals and contributors to HFpEF.

**Condensed Abstract:** We utilized a large, uniformly phenotyped, single cohort of heart failure sub-classified into heart failure with reduced (HFrEF) and preserved ejection fraction (HFpEF) based on current clinical definitions, to conduct detailed genetic analyses of the two HF sub-types. We found different genetic architectures and distinct genetic association profiles of HFrEF and HFpEF suggesting differences in underlying pathobiology. Furthermore, the low yield of HFpEF genome-wide association study (GWAS) compared to similarly powered HFrEF GWAS underscores the heterogeneity of HFpEF and the urgent need for developing consensus sub-phenotyping of HFpEF to improve the discovery in genetic mechanisms and therapeutic interventions.

## Introduction

Heart failure (HF) affects approximately 64 million people worldwide and 6.2 million adults in the United States.(1,2) While major advances in therapy have reduced the morbidity and mortality due to heart failure with reduced ejection fraction (HFrEF), there is significant residual risk of adverse outcomes.(3) Therapeutic options are limited for heart failure with preserved ejection fraction (HFpEF), which accounts for approximately half of all cases of HF, with large scale clinical trials largely failing to demonstrate conclusive benefits. (4,5) Agents that have reduced the progression of myocardial remodeling and reduced adverse outcomes in HFrEF have not demonstrated comparable benefit in HFpEF.

Genomic analyses of large cohorts represent promising approaches to better understand the pathobiology of HFrEF and HFpEF. (6,7) A recent GWAS meta-analysis of multiple cohorts of European ancestry has identified several genomic loci associated with unclassified HF, although similar genomic analyses focused on HFrEF and HFpEF are lacking.(8) The Million Veteran Program (MVP) is a large biobank linked to extensive national Veterans Affairs (VA) electronic health record (EHR) databases. Using algorithms developed to curate HFrEF and HFpEF phenotypes in the national VA databases based on current consensus definitions,(9) we extensively explored the genetic architecture of each HF subtype in a single large cohort in the MVP. In addition to demonstrating the disparate genetic underpinnings of HFrEF and HFpEF, our results highlight the marked heterogeneity of the HFpEF phenotype, and the urgent need to develop consensus approaches to sub-phenotype HFpEF to enable pathophysiological and therapeutic discovery.

## Methods

### Datasets

#### Million Veteran Program

The design of MVP has been previously described.(10) Veterans were recruited from over 60 Veterans Health Administration medical centers nationwide since 2011. A unique feature of MVP is the linkage of a large biobank to an extensive, national, database from 2003 onward that integrates multiple elements such as diagnosis codes, procedure codes, laboratory values, and imaging reports, which permits detailed phenotyping of this large cohort. MVP has received ethical and study protocol approval by the Veterans Affairs Central Institutional Review Board in accordance with the principles outlined in the Declaration of Helsinki.

#### UK Biobank

UK Biobank is a prospective study with over 500,000 participants aged 40– 69 years recruited in 2006–2010 with extensive phenotypic and genotypic data.(11)

### Phenotyping of Heart Failure, HFrEF, and HFpEF

Our HF phenotyping algorithms utilize both structured and unstructured data to ensure accuracy of the HF diagnosis, and natural language processing to ascertain all measurements of left ventricular function from imaging studies (i.e. echocardiograms) and from clinical notes, with the latter permitting capture of left ventricular ejection fractions (LVEF) measured outside the VA system. (12-14) Capture of all LVEFs ensured that we truly obtained the LVEF measured at the time of diagnosis of HF to allow proper identification of HFpEF and exclude any veteran with recovered LVEF from the HFpEF cohort. For this study, to increase the number of HFpEF patients included in the study, we utilized a less restrictive definition recently utilized in a study (15) that did not require that all LVEFs recorded after the baseline measurement also be ≥50%, or the use of diuretics and/or measurement of B-type natriuretic peptide at baseline (**Figure 1**). To ensure adequacy of this definition, we compared the genetic associations obtained in the cohort to genetic associations obtained in a cohort curated with the more restrictive definition used for our previous epidemiological studies. (12,13,16,17) Comorbid conditions were curated using International Classification of Diseases (ICD)-10 or ICD-9 codes as in our previous studies and described in the Supplementary Materials. (13)

**Figure 1.**
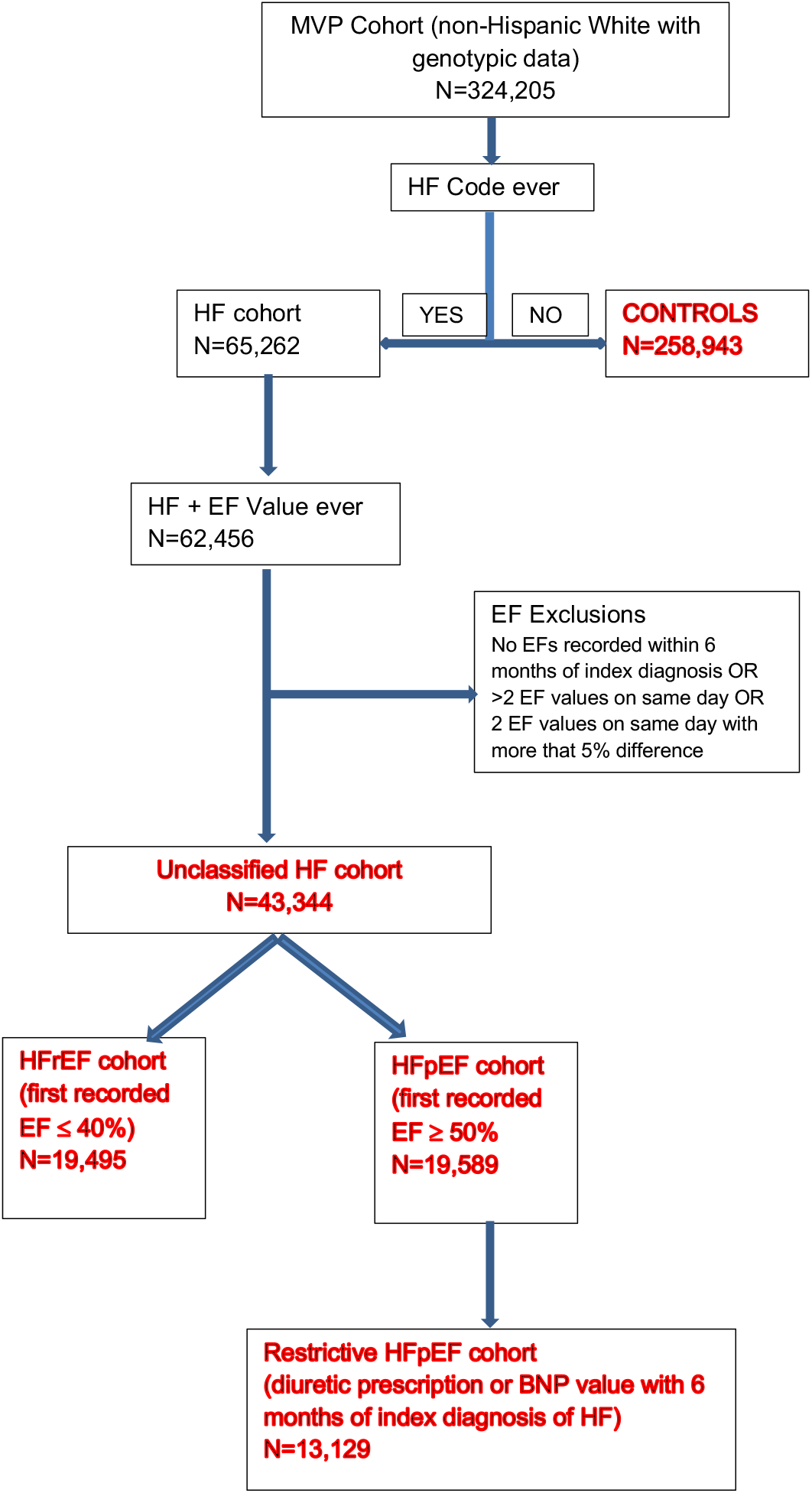
Algorithm for phenotyping of cohorts for genetic analyses. Consort diagram detailing the phenotyping of cases (unclassified HF, HFrEF, HFpEF, restrictive case definition of HFpEF) and controls.

In the UK Biobank, we defined HF as the presence of self-reported HF/pulmonary edema or cardiomyopathy at any visit; or an ICD-10 or ICD-9 billing code indicative of heart/ventricular failure or a cardiomyopathy of any cause, as described and validated previously, and consistent with that used in a recent, international collaborative effort (8,18) Assessments of LVEF were not available in the majority of UK Biobank participants to permit classification into HFpEF and HFrEF.

### Genetic Data Production, Quality Control and Imputation

DNA extracted from participants’ blood was genotyped using a customized Affymetrix Axiom® biobank array, the MVP 1.0 Genotyping Array. The array was enriched for both common and rare genetic variants of clinical significance in different ethnic backgrounds. Quality-control procedures used to assign ancestry, remove low-quality samples and variants, and perform genotype imputation were previously described. (19) We excluded: duplicate samples, samples with more heterozygosity than expected, an excess (>2.5%) of missing genotype calls, or discordance between genetically inferred sex and phenotypic gender.(19) In addition, one individual from each pair of related individuals (more than second degree relatedness as measured by the KING software)(20) were removed. Prior to imputation, variants that were poorly called (genotype missingness > 5%) or that deviated from their expected allele frequency observed in the 1000 Genomes reference data were excluded. After pre-phasing using EAGLE v2.4(21), we then imputed to the 1000 Genomes phase 3 version 5 reference panel (1000G) using Minimac4.(22) Genotyped SNPs after quality control were interpolated into the imputation file. Imputed variants with poor imputation quality (r^2^<0.3) were excluded from further analyses.

### Assignment of Racial/Ethnic Groups in the MVP

The MVP participants were assigned to mutually exclusive racial/ethnic groups using HARE (Harmonized Ancestry and Race/Ethnicity), a machine learning algorithm that integrates genetically inferred ancestry (GIA) with self-identified race/ethnicity (SIRE) as previously described. (23)

### Genome-wide Association Analysis

**Figure 2** demonstrates our study schema. Imputed and directly measured single nucleotide polymorphisms (SNPs) with minor allele frequency >1% were tested for association with HF, HFrEF, and HFpEF assuming an additive genetic model using PLINK2 (24) and adjusting for age, sex, and the top ten genotype-derived principal components. In UK Biobank analyses, genotyping array was included as an additional covariate. We meta-analyzed GWAS results of HF from MVP and UK Biobank using inverse-variance weighted fixed-effects model implemented in METAL.(25) Joint meta-analysis results were reported for unclassified HF to improve the power for GWAS discovery.(26) GWAS results were summarized using FUMA, a platform that annotates, prioritizes, visualizes and interprets GWAS results.(27) Genome-wide significant SNPs (P<5×10^−8^) were grouped into a genomic locus based on either r^2^ > 0.1 or distance between loci of < 500kb using the 1000 Genomes European reference panel. Lead SNPs were defined within each locus if they were independent (r^2^ < 0.1). We considered loci as novel if the sentinel SNP was of genome-wide significance (P<5×10^−8^) and located > 1 Mb from previously reported GWS SNPs associated with HF. (8,18) For novel loci, we used the genomic base-pair position of each sentinel SNP to map to the closest gene within a 500 kb region as the candidate gene. The physical base-pair location (GRCh37/hg19) and alleles were used to uniquely identify a genetic variant to replicate previous reported genetic associations with HF, and with HF risk factors.

**Figure 2.**
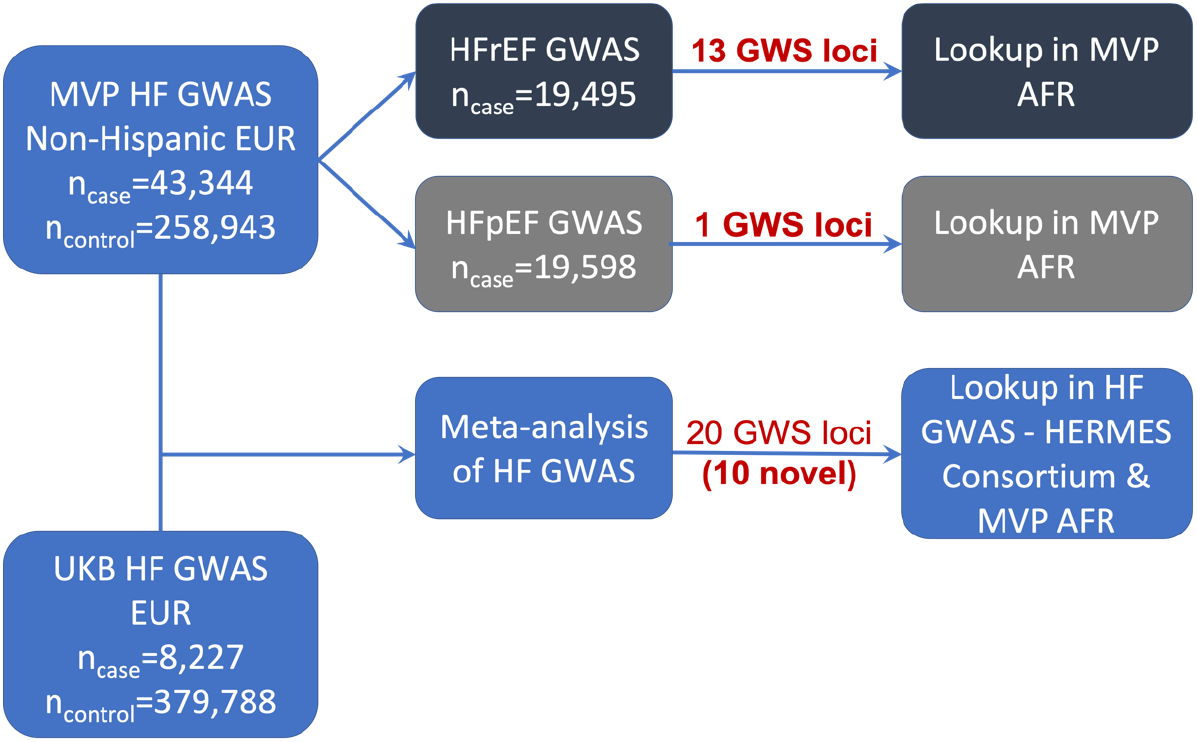
Study Schema.

For replication of unclassified HF, we conducted genome-wide association testing among UK Biobank participants passing sample quality control, comparing unclassified HF cases with non-HF controls. Procedures for genotyping and genotype imputation in the UK Biobank have been described previously.(11) For genetic association testing, we included SNPs with minor allele frequency (MAF) > 1% available in the Haplotype Reference Consortium (HRC), and imputation quality (INFO) > 0.3. We restricted analyses to samples of European genetic ancestry, defined by a combination of self-reported race and genetic principal components of ancestry. Specifically, we selected samples with genetic data who self-reported as white (British, Irish, or Other) and applied an outlier detection protocol (R package aberrant) to three pairs of principal components (PC1/PC2, PC3/PC4, and PC5/PC6), as generated centrally by the UK Biobank. Outliers in any of the three pairs of PCs were excluded from analysis to ensure that the study population was relatively homogenous in terms of genetic ancestry. Additional sample exclusions were implemented for 2nd-degree or closer relatedness (Kinship coefficient > 0.0884), sex chromosome aneuploidy, and excess missingness or heterozygosity, as defined by the UK Biobank. Association analyses were performed using PLINK2 (https://www.cog-genomics.org/plink/2.0/)25 on imputed genotype dosages, and a logistic regression model was used adjusting for age at enrollment, sex, genotyping array, and the first 10 principal components of ancestry. After merging with the phenotypic data, a total of 8,227 unclassified HF cases were comparted to 379,788 non-HF controls. Test statistic inflation was investigated by genomic control and inspection of quantile-quantile plots.

### Genetic Correlation

We estimated genetic correlations between these complex traits using cross-trait LD Score Regression and European ancestry-based GWAS results of HFpEF and HFrEF. (28,29) A reference panel consisting of 1.2 million HapMap3 variants was used to merge with GWAS summary statistics filtered to variants with MAF > 0.01, Hardy-Weinberg equilibrium P>10^−20^ and imputation R^2^ > 0.5. Using LD Score Regression and GWAS summary statistics, we also estimated the inflation factor of unclassified HF, HFpEF and HFrEF.

### Mendelian Randomization Analysis of HF Risk Factors

To assess differential causal associations of risk factors with HFrEF and HFpEF, we conducted two-sample Mendelian Randomization (MR). For MR, we utilized genetic instrumental variables reported in previous GWAS of the following traditional HF risk factors: coronary artery disease (CAD),(30) atrial fibrillation (AF),(31) type 2 diabetes (T2D),(32) body mass index (BMI),(33) lipids,(34) blood pressure(35) and estimated glomerular filtration rate (eGFR).(36) The GWS sentinel SNPs from each GWAS were selected as the genetic instrumental variables (GIVs) for each HF risk factor. We estimated the MR association of each risk factor using three complementary methods: inverse-variance-weighted, median weighted, and MR-Egger regression, as implemented in the R package TwoSampleMR.(37) MR-Egger regression was used to identify the horizontal pleiotropy measured by the intercept of the regression. Random-effects model was used to estimate the MR association between HF risk factors and HF outcomes for IVW and MR-Egger regression. To avoid sample overlap in the two-sample MR design, we used summary statistics of unclassified HF, HFrEF and HFpEF from the MVP study, and summary statistics of risk factors in previous GWAS without the MVP, all from studies of European ancestry. We considered nominal p-value of 0.05 as suggestive evidence for MR association for each HF risk factor. We applied a stringent Bonferroni correction for 12 tested factors (p-value<0.05/12=0.0042) acknowledging that some factors are not independent.

## RESULTS

The primary study population for the GWAS consisted of 258,943 controls, and cases of unclassified HF (n=43,344), HFpEF (n=19,589), and HFrEF (n=19,495) from the MVP cohort, and 8,227 HF cases and 379,788 controls from the UK Biobank cohort, all of European genetic ancestry. The GWS associations of unclassified HF, HFrEF and HFpEF were then examined in the MVP non-Hispanic African Americans and a recent HF GWAS in Europeans from the HERMES consortium (**Figure 2**). The MVP control and HF cohorts were predominantly male. In both MVP and UK Biobank, the HF cohorts tended to be older with a higher prevalence of cardiometabolic risk factors and comorbidities than the control populations (**Table 1** and **Supplementary Table 1**, and **Supplementary Table 2**).

**Table 1.** Characteristics of HF Patients and non-HF Controls in the MVP Participants of European Ancestry.

| Group | Control<br>(N=258,943) | HFpEF<br>(N=19,589) | HFrEF<br>(N=19,495) | Unclassified HF<br>(N=43,344) |
| --- | --- | --- | --- | --- |
| Age (years), mean±SD | 62.74±13.76 | 69.88±9.77 | 69.29±9.74 | 69.61±9.74 |
| Male (%) | 92.14 | 95.74 | 97.85 | 96.92 |
| Body mass index (kg/m<sup>2</sup>), mean±SD | 29.20±5.53 | 31.95±6.98 | 30.20±6.38 | 31.08±6.73 |
| Underweight (< 18.5) % | 0.56 | 0.47 | 0.59 | 0.52 |
| Normal (18.5-24.9) % | 20.25 | 13.43 | 18.79 | 16.05 |
| Overweight (25.0-29.9) % | 40.66 | 29.71 | 35.09 | 32.44 |
| Obese (30.0-34.9) % | 24.70 | 27.08 | 25.62 | 26.37 |
| Morbidly obese (≥ 35.0) % | 13.84 | 29.31 | 19.91 | 24.62 |
| LVEF, mean±SD | NA | 56.97±5.65 | 29.33±9.36 | 43.36±15.05 |
| Atrial fibrillation (%) | 6.33 | 30.80 | 37.83 | 34.44 |
| Coronary artery disease (%) | 22.47 | 63.87 | 74.63 | 69.72 |
| Chronic kidney disease (%) | 9.54 | 37.21 | 35.75 | 36.43 |
| Diabetes (%) | 20.61 | 48.54 | 45.06 | 46.76 |
| Hyperlipidemia (%) | 66.9 | 87.75 | 88.20 | 88.04 |
| Hypertension (%) | 62.97 | 93.22 | 91.69 | 92.51 |
| Peripheral vascular disease (%) | 15.18 | 42.47 | 42.27 | 42.47 |
| Stroke/TIA (%) | 8.26 | 25.29 | 24.33 | 24.93 |

Note: HFpEF: heart failure with preserved ejection fraction; HFrEF: heart failure with reduced ejection fraction; HF: heart failure; SD: standard deviation; LVEF: left ventricular ejection fraction; TIA: transient ischemic attack.

### GWAS of Unclassified HF

In unclassified HF, the meta-analysis of MVP and UKB GWAS results (**Supplementary Figures 1 and 2**) identified 20 genome-wide significant (GWS) loci including 10 novel loci (**Table 2 and Supplementary Tables 3 and 4**). The regional association plots of each GWS locus are shown in **Supplementary Figure 3A-3T**. We replicated all 12 GWS independent SNPs associated with HF from a recent HF GWAS publication,(8) (Bonferroni-corrected p-value < 0.05; **Supplementary Table 5**).

**Table 2.** Sentinel SNPs significantly associated with heart failure.

| rsID | Position | Closest Gene<br>(* denotes novel association) | Genomic Region | Risk allele/Ref. allele | Risk allele frequency | META HF GWAS |  | MVP HF GWAS |  |
| --- | --- | --- | --- | --- | --- | --- | --- | --- | --- |
|  |  |  |  |  |  | OR (95% CI) | p-value | OR (95% CI) | p-value |
| rs371236917 | 1:16310737 | *SRARP/HSPB7/ZBTB17* | flanking | C/CT | 0.70 | 1.06 (1.05, 1.08) | $4.97 \times 10^{-15}$ | 1.06 (1.04, 1.08) | $1.50 \times 10^{-12}$ |
| rs1277930 | 1:109822143 | *CELSR2* | flanking | A/G | 0.77 | 1.05 (1.04, 1.07) | $1.10 \times 10^{-10}$ | 1.05 (1.03, 1.07) | $7.20 \times 10^{-8}$ |
| rs7595697 | 2:11568158 | *E2F6\** | flanking | T/C | 0.37 | 1.04 (1.02, 1.05) | $4.98 \times 10^{-8}$ | 1.04 (1.02, 1.05) | $1.05 \times 10^{-6}$ |
| rs6795366 | 3:44005735 | *ABHD5\** | intergenic | C/T | 0.74 | 1.05 (1.03, 1.06) | $1.95 \times 10^{-8}$ | 1.04 (1.02, 1.06) | $5.36 \times 10^{-6}$ |
| rs2634073 | 4:111665783 | *PITX2* | intergenic | T/C | 0.20 | 1.08 (1.06, 1.10) | $7.42 \times 10^{-19}$ | 1.07 (1.05, 1.09) | $1.63 \times 10^{-11}$ |
| rs3176326 | 6:36647289 | *CDKN1A* | intron | G/A | 0.80 | 1.08 (1.06, 1.10) | $1.08 \times 10^{-18}$ | 1.08 (1.06, 1.10) | $1.00 \times 10^{-15}$ |
| rs10455872 | 6:161010118 | *LPA* | intron | G/A | 0.07 | 1.11 (1.08, 1.14) | $9.34 \times 10^{-17}$ | 1.11 (1.08, 1.14) | $7.73 \times 10^{-13}$ |
| rs4717903 | 7:74068167 | *GTF2I\** | flanking | C/T | 0.25 | 1.04 (1.03, 1.06) | $3.55 \times 10^{-8}$ | 1.04 (1.02, 1.06) | $1.33 \times 10^{-5}$ |
| rs4977575 | 9:22124744 | *CDKN2B-AS* | intergenic | G/C | 0.49 | 1.07 (1.06, 1.09) | $6.92 \times 10^{-23}$ | 1.06 (1.05, 1.08) | $3.87 \times 10^{-16}$ |
| rs579459 | 9:136154168 | *ABO* | flanking | C/T | 0.22 | 1.05 (1.03, 1.06) | $1.26 \times 10^{-8}$ | 1.04 (1.02, 1.06) | $3.43 \times 10^{-6}$ |
| rs59693993 | 10:75583034 | *CAMK2G* | intron | C/T | 0.86 | 1.06 (1.04, 1.08) | $3.08 \times 10^{-8}$ | 1.05 (1.03, 1.08) | $1.79 \times 10^{-6}$ |
| rs61869036 | 10:121422836 | *BAG3* | intron | G/C | 0.79 | 1.06 (1.04, 1.08) | $1.76 \times 10^{-11}$ | 1.04 (1.03, 1.06) | $3.13 \times 10^{-6}$ |
| rs12149832 | 16:53842908 | *FTO* | intron | A/G | 0.41 | 1.07 (1.05, 1.08) | $3.40 \times 10^{-21}$ | 1.07 (1.06, 1.09) | $9.05 \times 10^{-20}$ |
| rs12933292 | 16:69566309 | *NFAT5\** | intergenic | C/G | 0.59 | 1.04 (1.03, 1.06) | $5.25 \times 10^{-9}$ | 1.04 (1.03, 1.06) | $2.75 \times 10^{-7}$ |
| rs1002135 | 17:2097583 | *SMG6\** | intron | G/T | 0.38 | 1.04 (1.03, 1.06) | $6.33 \times 10^{-9}$ | 1.04 (1.02, 1.06) | $5.89 \times 10^{-7}$ |
| rs12150603 | 17:37834715 | *PNMT/PGAP3\** | intron | G/A | 0.35 | 1.04 (1.03, 1.06) | $5.21 \times 10^{-9}$ | 1.05 (1.03, 1.06) | $5.78 \times 10^{-8}$ |
| rs150947345 | 17:57486425 | *YPEL2\** | flanking | A/T | 0.02 | 1.16 (1.10, 1.22) | $1.67 \times 10^{-8}$ | 1.16 (1.09, 1.23) | $1.62 \times 10^{-6}$ |
| rs34432450 | 17:65880259 | *BPTF\** | intron | C/T | 0.21 | 1.06 (1.04, 1.08) | $1.93 \times 10^{-12}$ | 1.06 (1.04, 1.08) | $3.30 \times 10^{-10}$ |
| rs79329549 | 18:36560942 | *18q12.2\** | intergenic | T/G | 0.91 | 1.07 (1.05, 1.10) | $4.60 \times 10^{-9}$ | 1.08 (1.05, 1.11) | $5.24 \times 10^{-8}$ |
| rs1999323 | 21:30534128 | *MAP3K7CL\** | intron | T/C | 0.15 | 1.07 (1.05, 1.09) | $5.26 \times 10^{-11}$ | 1.05 (1.03, 1.07) | $4.04 \times 10^{-6}$ |

Note: chromosomal position is based on GRCh37/hg19 reference. The sentinel SNPs were mapped to the closed refseq genes based on chromosomal base-pair position. All genetic associations were aligned to effects of the risk alleles (i.e., increased risk for unclassified
HF). Ref: reference; OR: odds ratio; CI: confidence interval; GWAS: genome-wide association study. MVP - Million Veteran Program cohort ( $n_{\text{cases}}=43,344$ ) META – meta-analysis of MVP and UK Biobank cohorts).

### GWAS of HFrEF and HFpEF

We conducted GWAS of our established definitions of HFrEF and HFpEF. First, we compared the output of GWAS for the more and less restrictive HFpEF definitions and observed high, overall genetic correlation (r=0.981, p<2×10-16) between these phenotypes, including among the top 110 HFpEF-associated SNPs (r=0.995, p<2×10-16; **Supplementary Figure 4**). We therefore used the less restrictive (and better-powered) HFpEF definition as the primary HFpEF phenotype for all subsequent analyses.

In the GWAS among the MVP participants of European ancestry, we identified 13 GWS loci associated with HFrEF and one GWS locus (*FTO*) associated with HFpEF (**Figure 3**; **Table 3; Supplementary Figure 5A and 5B**). The regional association plots of each GWS locus are shown in **Supplementary Figure 6A-6N**. Two lead SNPs in the *FTO* locus for HFrEF (rs7188250) and HFpEF (rs11642015) were in linkage disequilibrium (r^2^=0.873). Among these thirteen loci associated with HF subtypes, seven loci (*NFIA, E2F6, MITF, PHACTR1, METTL7A, PNMT and BPTF*) have not been reported in previous HF-related GWAS, of which four loci (*NFIA, MITF, PHACTR1 and METTL7A*) were GWS only in GWAS of HFrEF cases.

**Figure 3.**
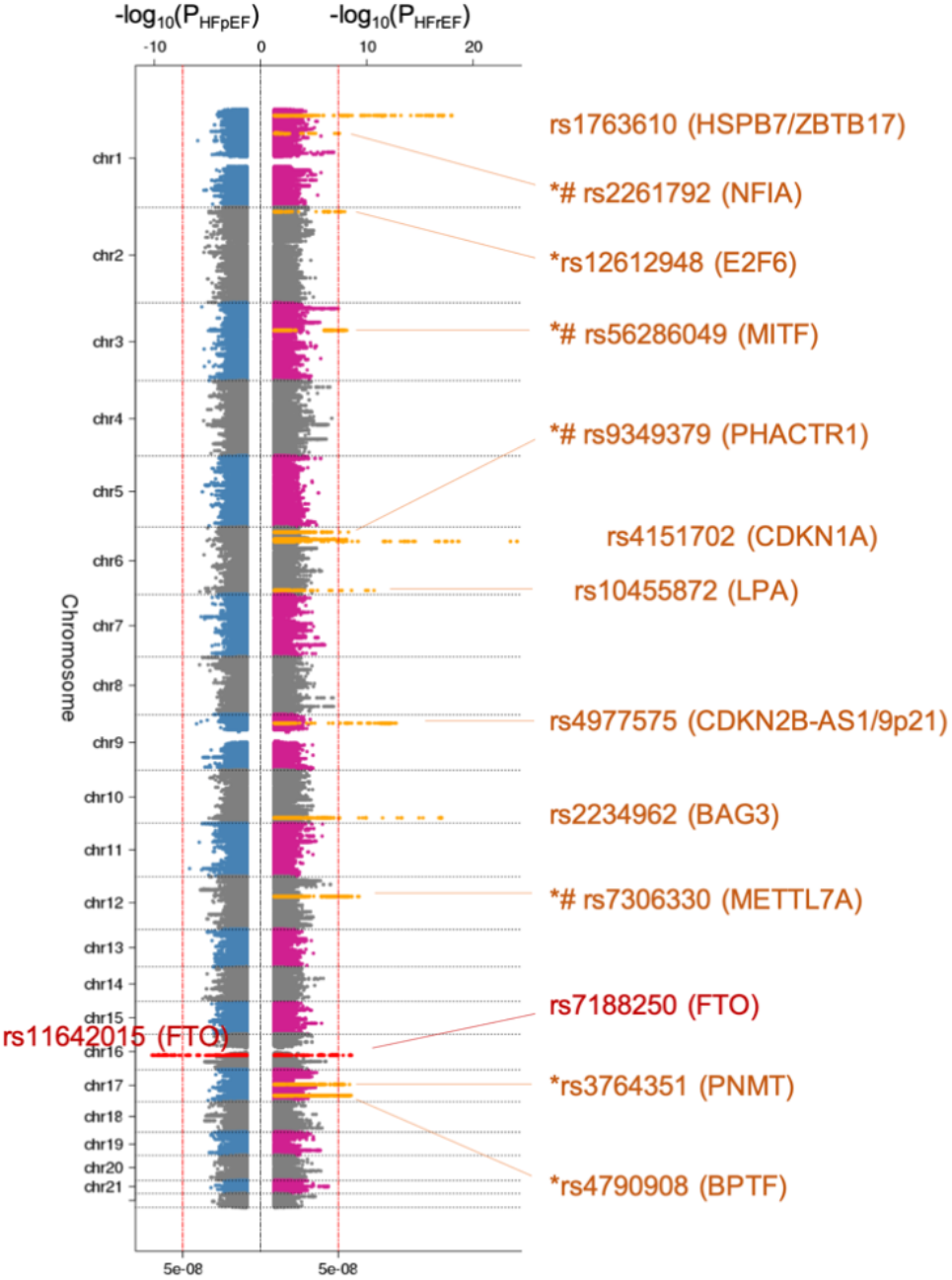
Genome-wide associations of HFrEF and HFpEF. Genome-wide significant loci association studies of HFpEF and HFrEF among non-Hispanic White veterans. Sentinel SNPs and the nearest mapped genes are shown. Y-axis shows chromosomal position. Sentinel SNPs and their nearest genes are shown. *: novel HF locus; #: unique locus in the HFrEF GWAS but not in the HF meta-analysis; dashed vertical line indicates genome-wide significance threshold (P=5×10^−8^).

**Table 3.**
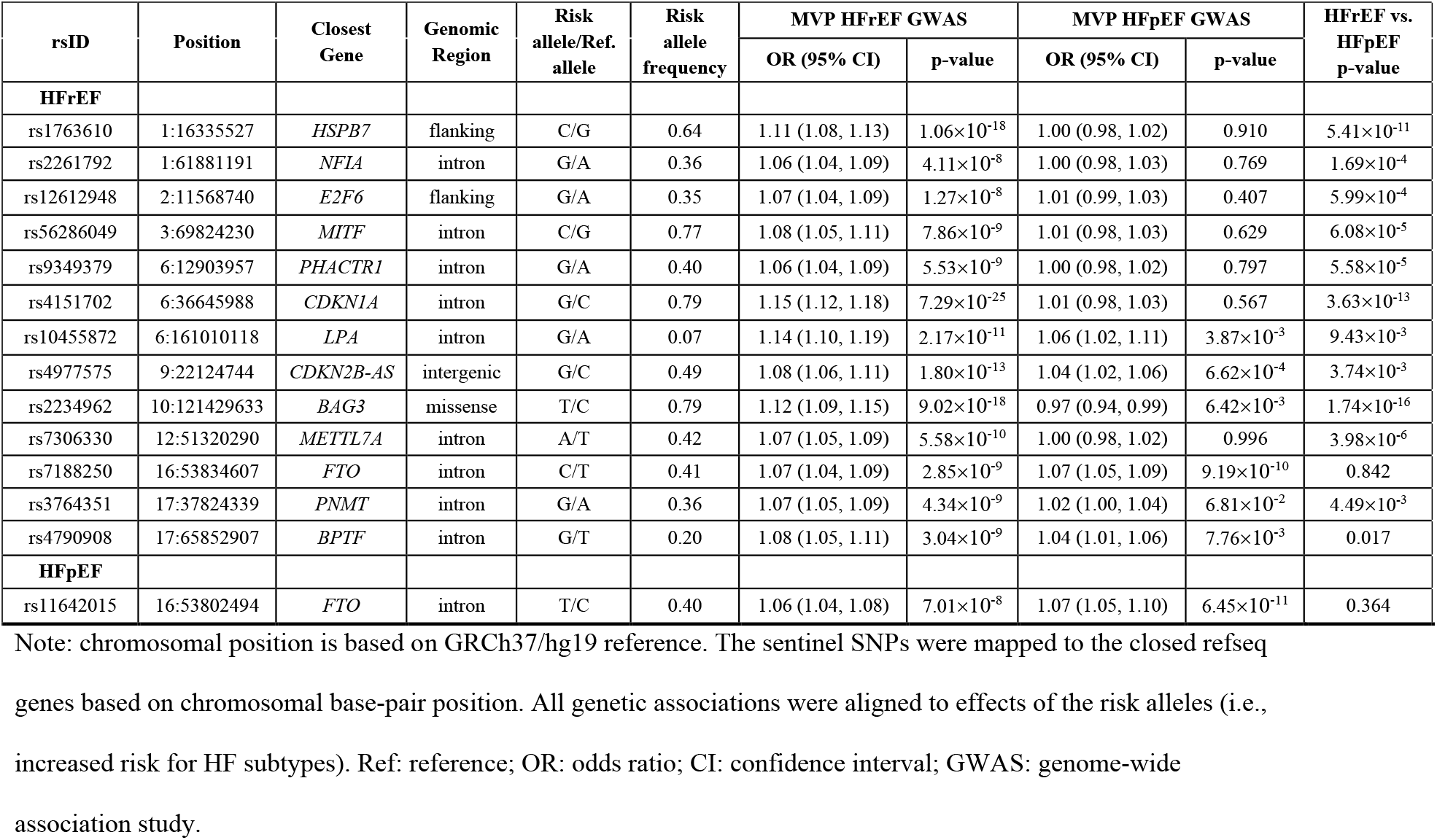
Sentinel SNPs significantly associated with HFrEF (19,495 cases) and HFpEF (19,589 cases).

| rsID | Position | Closest Gene | Genomic Region | Risk allele/Ref. allele | Risk allele frequency | MVP HFrEF GWAS |  | MVP HFpEF GWAS |  | HFrEF vs. HFpEF p-value |
| --- | --- | --- | --- | --- | --- | --- | --- | --- | --- | --- |
|  |  |  |  |  |  | OR (95% CI) | p-value | OR (95% CI) | p-value |  |
| **HFrEF** |  |  |  |  |  |  |  |  |  |  |
| rs1763610 | 1:16335527 | *HSPB7* | flanking | C/G | 0.64 | 1.11 (1.08, 1.13) | $1.06 \times 10^{-18}$ | 1.00 (0.98, 1.02) | 0.910 | $5.41 \times 10^{-11}$ |
| rs2261792 | 1:61881191 | *NFIA* | intron | G/A | 0.36 | 1.06 (1.04, 1.09) | $4.11 \times 10^{-8}$ | 1.00 (0.98, 1.03) | 0.769 | $1.69 \times 10^{-4}$ |
| rs12612948 | 2:11568740 | *E2F6* | flanking | G/A | 0.35 | 1.07 (1.04, 1.09) | $1.27 \times 10^{-8}$ | 1.01 (0.99, 1.03) | 0.407 | $5.99 \times 10^{-4}$ |
| rs56286049 | 3:69824230 | *MITF* | intron | C/G | 0.77 | 1.08 (1.05, 1.11) | $7.86 \times 10^{-9}$ | 1.01 (0.98, 1.03) | 0.629 | $6.08 \times 10^{-5}$ |
| rs9349379 | 6:12903957 | *PHACTR1* | intron | G/A | 0.40 | 1.06 (1.04, 1.09) | $5.53 \times 10^{-9}$ | 1.00 (0.98, 1.02) | 0.797 | $5.58 \times 10^{-5}$ |
| rs4151702 | 6:36645988 | *CDKN1A* | intron | G/C | 0.79 | 1.15 (1.12, 1.18) | $7.29 \times 10^{-25}$ | 1.01 (0.98, 1.03) | 0.567 | $3.63 \times 10^{-13}$ |
| rs10455872 | 6:161010118 | *LPA* | intron | G/A | 0.07 | 1.14 (1.10, 1.19) | $2.17 \times 10^{-11}$ | 1.06 (1.02, 1.11) | $3.87 \times 10^{-3}$ | $9.43 \times 10^{-3}$ |
| rs4977575 | 9:22124744 | *CDKN2B-AS* | intergenic | G/C | 0.49 | 1.08 (1.06, 1.11) | $1.80 \times 10^{-13}$ | 1.04 (1.02, 1.06) | $6.62 \times 10^{-4}$ | $3.74 \times 10^{-3}$ |
| rs2234962 | 10:121429633 | *BAG3* | missense | T/C | 0.79 | 1.12 (1.09, 1.15) | $9.02 \times 10^{-18}$ | 0.97 (0.94, 0.99) | $6.42 \times 10^{-3}$ | $1.74 \times 10^{-16}$ |
| rs7306330 | 12:51320290 | *METTL7A* | intron | A/T | 0.42 | 1.07 (1.05, 1.09) | $5.58 \times 10^{-10}$ | 1.00 (0.98, 1.02) | 0.996 | $3.98 \times 10^{-6}$ |
| rs7188250 | 16:53834607 | *FTO* | intron | C/T | 0.41 | 1.07 (1.04, 1.09) | $2.85 \times 10^{-9}$ | 1.07 (1.05, 1.09) | $9.19 \times 10^{-10}$ | 0.842 |
| rs3764351 | 17:37824339 | *PNMT* | intron | G/A | 0.36 | 1.07 (1.05, 1.09) | $4.34 \times 10^{-9}$ | 1.02 (1.00, 1.04) | $6.81 \times 10^{-2}$ | $4.49 \times 10^{-3}$ |
| rs4790908 | 17:65852907 | *BPTF* | intron | G/T | 0.20 | 1.08 (1.05, 1.11) | $3.04 \times 10^{-9}$ | 1.04 (1.01, 1.06) | $7.76 \times 10^{-3}$ | 0.017 |
| **HFpEF** |  |  |  |  |  |  |  |  |  |  |
| rs11642015 | 16:53802494 | *FTO* | intron | T/C | 0.40 | 1.06 (1.04, 1.08) | $7.01 \times 10^{-8}$ | 1.07 (1.05, 1.10) | $6.45 \times 10^{-11}$ | 0.364 |

Note: chromosomal position is based on GRCh37/hg19 reference. The sentinel SNPs were mapped to the closed refseq genes based on chromosomal base-pair position. All genetic associations were aligned to effects of the risk alleles (i.e., increased risk for HF subtypes). Ref: reference; OR: odds ratio; CI: confidence interval; GWAS: genome-wide association study.

Among 13 HFrEF-associated loci, nine loci had different associations with HFrEF and HFpEF (p-value<0.0038, corrected for 13 tests, **Table 3**). For example, the risk allele of the *BAG3* missense variant (rs2234962) was associated with higher risk for HFrEF (OR 1.12, 95% CI 1.09-1.15, p-value 9.02×10^−18^), but was associated with lower risk for HFpEF (OR 0.97, 95% CI 0.94-0.99, p-value 6.42×10^−3^). Only four loci, including *LPA, FTO, PNMT* and *BPTF*, were not differentially associated with HF subtypes.

We observed moderate genomic inflation (λ) for unclassified HF (λ= 1.263), HFrEF (λ= 1.152) and HFpEF (λ= 1.118), on par in with GWAS of phenotypes with similarly large sample sizes. The LDSC intercepts were 1.044 (SE 0.010), 1.013 (SE 0.008) and 1.028 (SE 0.008) for unclassified HF, HFrEF and HFpEF, respectively, indicating that most of the inflation was due to polygenicity of HF and subtypes.

### Replication in MVP African Americans and other HF GWAS

Among MVP African Americans, all but two of the SNPs identified in the GWAS of unclassified HF in the European ancestry had genetic associations with unclassified HF in the same direction, and two (rs3176326-CDKN1A and rs12150603-PNMT) were significant after Bonferroni correction (**Supplementary Table 4**); four (rs4717903-GTF2I, rs12933292-NFAT5, rs1002135-SMG6, and rs1999323-MAP3K7CL) were replicated in the recent HF GWAS (8) after Bonferroni correction.

Among 13 GWS loci associated with HFrEF, 11 had genetic effects in the same direction in the MVP African American cohort (**Supplementary Table 6**), including three (rs1763610-HSPB7, rs4151702-CDKN1A, and rs2234962-BAG3) which were test-wise significant after Bonferroni correction. Interestingly, the sentinel SNP of the *FTO* locus was significantly associated with HFpEF (rs11642015, OR 1.10, 95% CI 1.03-1.17, p-value 6.30×10^−3^), but not associated with HFrEF (rs7188250, OR 1.06, 95% CI 0.99-1.12, p-value 0.11).

### Genetic Associations with HFrEF and HFpEF in Candidate Genes and Loci

Out of 12 GWS loci reported in the recent HERMES study of unclassified HF, all were associated with HFrEF, but only four were significantly associated with HFpEF including the *FTO* locus (**Supplementary Table 5**). Other loci replicated in HFrEF were *ZBTB17/HSPB7* locus (closest gene of *SRARP* discovered in our study) and *HCG22* locus (38) (OR 1.05, CI 1.03-1.08, P=7.83×10^−5^). We did not replicate previously reported associations of *FRMD4B* or *USP3* region with HF.(6,39) Among 15 autosomal genes related to cardiomyopathy,(40) we found significant associations in HFrEF with only *TMEM43* (**Supplementary Table 7, Supplementary Figure 7**).

### Associations of HFrEF- and HFpEF Loci with Cardiovascular Risk Factors

As shown in **Figure 4** and **Supplementary Table 8**, several of the 13 loci associated with HFrEF and HFpEF also demonstrated genetic associations with risk factors as previously reported (*PHACTR1, LPA*, and *CDKN2B-AS* with CAD; *CDKN1A* with AF); and *FTO* with BMI, T2D, and HDL cholesterol. Although most loci were associated with multiple risk factors, the *BAG3* locus was only associated with blood pressure traits, and the *MITF* and *METTL7A* loci were associated with eGFR. Three novel loci, *SRARP, NFIA* and *E2F6*, were not significantly associated with any tested HF risk factors.

**Figure 4.**
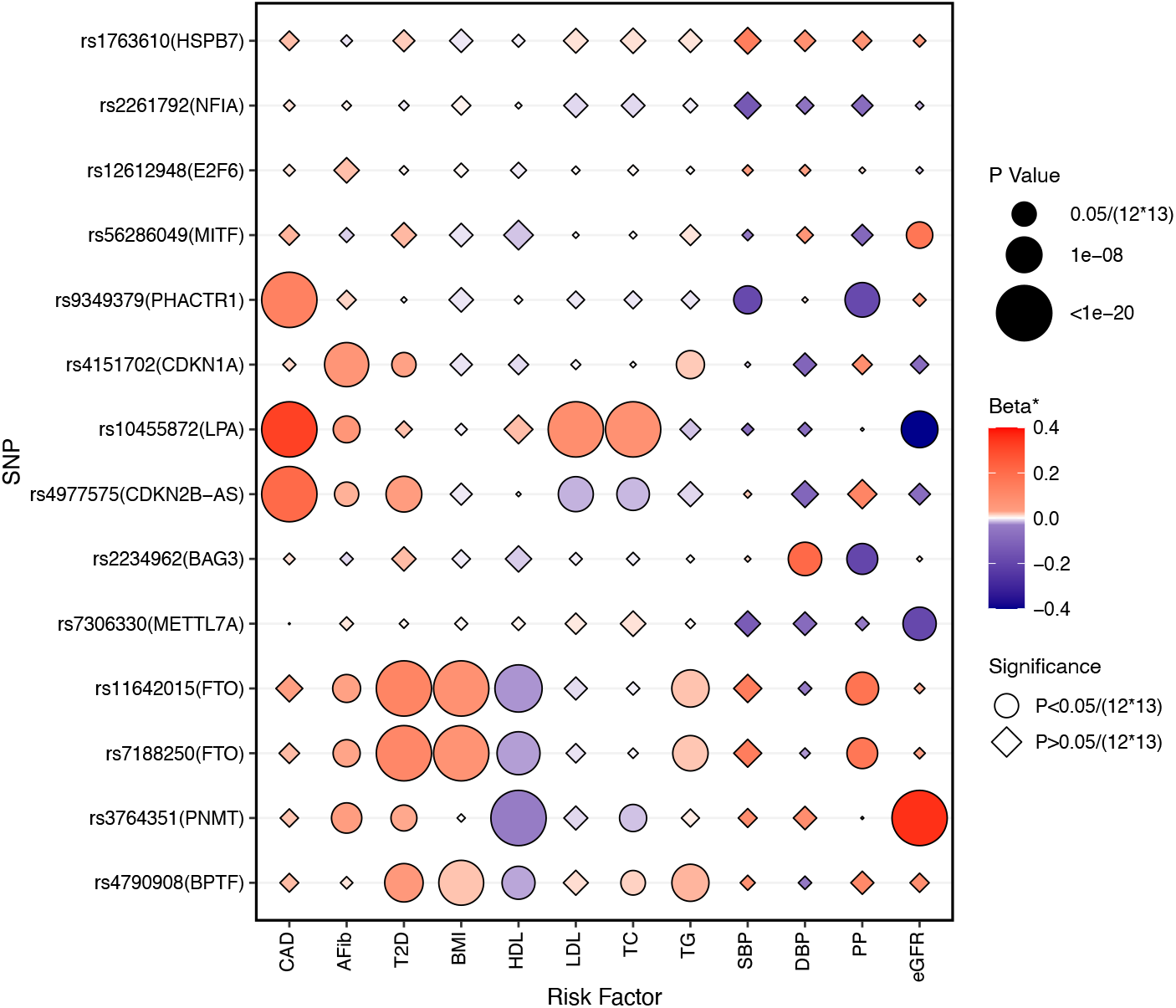
Genetic associations between HFrEF/HFpEF risk variants and HF risk factors. The genetic associations were identified from published GWAS of HF risk factors. Beta: beta coefficients for continuous risk factors, log(odds ratio) for binary risk factors, percent change in eGFR. CAD: coronary artery disease; AFib: atrial fibrillation; T2D: type 2 diabetes; BMI: body mass index; HDL: high-density lipoprotein cholesterol; LDL: low-density lipoprotein cholesterol; TC: total cholesterol; TG: triglycerides; SBP: systolic blood pressure; DBP: diastolic blood pressure; PP: pulse pressure; eGFR: estimated glomerular filtration rate.

### Genetic Correlation Between HFrEF and HFpEF and Heritability

Using LDSC and the MVP GWAS summary statistics, we estimated the heritability (*h*^*2*^) of unclassified HF, HFpEF and HFrEF as 3.7% (SE 0.3%), 1.9% (SE 0.2%) and 3.1% (0.3%), respectively. Heritability of HFpEF was substantially lower than that of unclassified HF and HFrEF. We also identified a modest positive genetic correlation between HFrEF and HFpEF (0.57±0.07).

### Mendelian Randomization Association Analysis of HF Risk Factors

We present the MR association results from the inverse-variance-weighted method (**Figure 5**) since the assumption of zero-intercept was not violated in the Egger regression (**Supplementary Table 9** shows results of all 3 MR methods). In primary MR analyses (inverse-variance-weighted estimates), CAD had a stronger causal association with HFrEF, and all lipid parameters as well as T2D and DBP had a significant causal association only with HFrEF. While AF, BMI, and SBP demonstrated similar causal associations with both HF subtypes, PP was significantly associated with HFpEF only. Similar results were observed from the median weighted method (**Supplementary Table 9**). Sensitivity analysis using Egger regression showed consistent effect estimates but larger confidence intervals (**Supplementary Table 9**).

**Figure 5.**
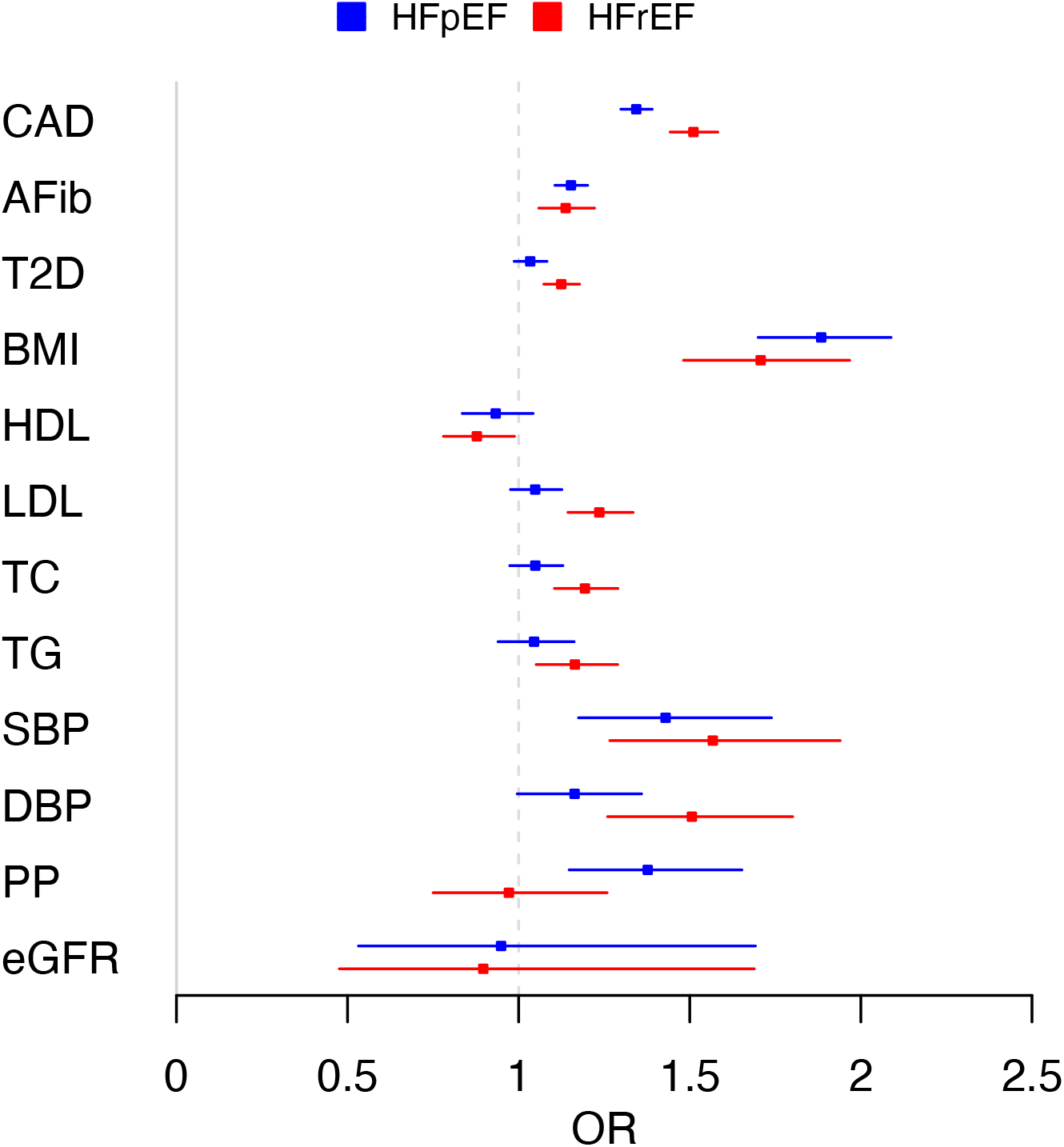
Mendelian randomization analysis of HF risk factors in relation to HFpEF and HFrEF. CAD: coronary artery disease; AFib: atrial fibrillation; T2D: type 2 diabetes; BMI: body mass index; HDL: high-density lipoprotein cholesterol; LDL: low-density lipoprotein cholesterol; TC: total cholesterol; TG: triglycerides; SBP: systolic blood pressure; DBP: diastolic blood pressure; PP: pulse pressure; eGFR: estimated glomerular filtration rate.

## DISCUSSION

In our large-scale genetic association analysis of clinical HF subtypes, we found pronounced differences in the genetic architectures of HFrEF and HFpEF. The very limited genetic discovery in HFpEF in spite of a large cohort size similar to HFrEF, suggests that HFpEF as currently clinically defined is a heterogenous phenotype with varying underlying pathobiology across the phenotype (**Central Illustration**).

Our study of HFpEF as universally defined based on current guidelines(9) suggests underlying pathophysiological heterogeneity as a plausible explanation for the neutral results of clinical trials using therapies targeting specific pathophysiologic mechanisms.(5) Clinical trials have used the universal definition but have frequently changed the LVEF cutoff to below 50%; yet the overall success of these trials is low in terms of demonstrating reductions in mortality and morbidity.(4) Even the recently reported benefits of sodium-glucose co-transporter-2 inhibitors were not specific to HFpEF, but rather were seen across the spectrum of HF (both HFrEF and HFpEF).(41) The paucity of genetic discovery in HFpEF is like that observed in stroke, another condition with heterogenous pathogenesis. For example, in a GWAS of 72,147 stroke patients from two large biobanks, only 3 novel loci were discovered. (42) While previous small studies have examined both *a priori* grouping into sub-phenotypes(43), and unbiased clustering using machine learning approaches(44), our findings suggest an urgent need to develop consensus sub-phenotyping strategies to resolve the heterogeneity of HFpEF as currently defined, as will be the focus of the recently initiated National Institutes of Health HeartShare Program (https://grants.nih.gov/grants/guide/rfa-files/RFA-HL-21-015.html).

Our genetic analyses of the associations between HF risk factors and HF subtypes, and causal relations of HF risk factors to HFrEF and HFpEF confirmed current epidemiologic data and the validity of our cohorts. For example, we found strong genetic associations of CAD and lipid with HFrEF. Conversely, genetically-determined pulse pressure was more associated with HFpEF. Atrial fibrillation and BMI were causally related to both HFrEF and HFpEF. At the level for individual variants, for e.g., in case of the myocardial variant BAG3, different associations were seen with HFpEF and HFrEF. Our finding that the direct genetic correlation between HFrEF and HFpEF was modest (r^2^ approximately 32%) reinforces our findings at the genomic level that HFrEF and HFpEF have different genetic architecture.

### Study Limitations

Our findings should be interpreted in the context of the strengths and limitations of the study. This study is the first large-scale genomic analyses of HFpEF and HFrEF. Our HFpEF cohort had less women compared to epidemiologic studies and recent clinical trials; however, the genetic and causal associations of risk factors with HFpEF as compared to HFrEF mirrored associations seen in epidemiologic studies. Since we utilized natural language processing to capture all recorded LVEFs including measurements performed outside the VA, our cohort of HFpEF excluded any participants with previously reduced and currently normal LVEF. We ensured adequate power by increasing the size of our HFpEF cohort with a less restrictive algorithm for curation, which demonstrated very high genetic correlation with a smaller cohort curated utilizing the more restrictive algorithm. Hence our findings indicate that the issue with reduced genetic discovery in our cohort was not secondary to impurity of the phenotype due to EMR-based curation, but rather that HFpEF as currently defined may be a collection of sub-phenotypes with multiple independent disease mechanisms. Our case and control cohorts, since they were recruited from a hospital setting, had a higher prevalence of comorbidities compared to a population-based cohort. We could not externally replicate our findings since currently there are no other large adequately phenotyped cohorts of HFpEF and HFrEF.

## Conclusions

The genetic architectures of HFpEF and HFrEF differ significantly. HFpEF as currently clinically defined is a pathophysiologically heterogenous disease that requires further characterization into consensus sub-phenotypes to enhance genetic discovery. Better genetic understanding of HF subtypes will lead to precise diagnosis, accurate risk assessment, and effective treatment and management of the global pandemic of heart failure.

### Perspectives

#### Competency in Medical Knowledge

Large-scale genetic studies help in understanding pathobiology of complex diseases. While heart failure with reduced and preserved ejection fraction qualitatively share similar risk factors, their genetic underpinnings are different. Heart failure with preserved ejection fraction is a heterogenous condition with likely different disease mechanisms underlying its genesis.

#### Translational Outlook 1

Consensus sub-phenotyping strategies are urgently needed for therapeutic advances in heart failure with preserved ejection fraction.

#### Translational Outlook 2

Multi-omics studies may be useful in sub-phenotyping heart failure with preserved ejection fraction to enable discovery of novel therapeutic targets.

## Data Availability

All data produced in the present study are available upon reasonable request to the authors

## Acknowledgements

We are grateful to all the MVP investigators; a list of MVP investigators can be found in Supplementary Materials.

## Abbreviations

GWAS: Genome-wide association study
GWS: Genome wide significant
HF: Heart Failure
HFpEF: Heart failure with preserved ejection fraction
HFrEF: Heart failure with reduced ejection fraction
MVP: Million Veteran Program
SNP: Single nucleotide polymorphism
VA: Veterans Affairs

## Figure Titles and Captions

**Central Illustration. Limited genetic discovery in HFpEF due to pathophysiological heterogeneity**

