## Supplementary Methods and Figures for "Genetic Architecture of Heart Failure with Preserved versus Reduced Ejection Fraction"

**Supplementary Information**

**Supplementary Methods**

Comorbidities were curated from the national databases using the International Classification of Diseases (ICD)-10 and ICD-9 codes. The codes used to extract each comorbidity are listed below (% sign indicates that any code with the preceding number was utilized).
Atrial Fibrillation:
ICD-9: 427.31%
ICD-10: I48.2%, I48.0%, I48.91%, I48.1%
Coronary Artery Disease:
ICD-9: 410%, 411%, 412%, 413%, 414%
ICD-10: I20%, I21%, I22%, I23%, I24%, I25%
Chronic Kidney Disease:
ICD-9: 403.01%, 404.02%, 404.10%, 585.00%, 403.11%, 404.03%, 404.90%, 586.00%, 403.91%, 404.10%, 584.00%, 792.50%
ICD-10: V42.0%, V56.0%, V56.3%, N18%, V45.1%, V56.1%, V56.8%, V56.2%
Diabetes Mellitus:
ICD-9: 250%
ICD-10: E08%, E09%y, E10%, E11%, E12%, E13%
Hyperlipidemia:
ICD-9: 272%
ICD-10: E78.0, E75.249, E78.01, E77.0, E78.1, E77.1, E78.2, E78.81, E78.3, E78.89, E78.4, E78.9, E78.5, E88.1, E78.6, E88.89, E75.21, E75.22
Hypertension:
ICD-9: 401%, 402%, 403%, 404%, 405%
ICD-10: I10%, I11%, I12%, I13%, I15%, I16%, I67.4%
Peripheral Vascular Disease:
ICD-9: 440%, 441%, 442%, 443%, 444%, 447%, 451%, 452%, 453%, 557%
ICD-10: E08.51, E08.52, E09.51, E09.52, E10.51, E10.52, E11.51, E11.52, E13.51, E13.52, I67.0, I70.0, I70%, I71%, I72%, I73.01, I73.1, I73.81, I73.89, I73.9, I74.01, I74.09, I74.10, I74.11, I74.19, I74.2, I74.3, I74.4, I74.5, I74.8, I74.9, I77%, I79.0, I79.1, I79.8, I80%, I81, I82%, K55%, K76.5, M31.8, M31.9
Stroke or Transient Ischemic Attack:
ICD-9: 430%, 431%, 433%, 434%, 436%, 437.0%, 437.6%
ICD-10: I63%, I65%, I66%, I67.2%, I67.8%, I67.6%, I60%, I61%, G45%

### Supplementary Figures

**Supplementary Figure 1.** Quantile-quantile plot of genome-wide meta-analysis of heart failure.

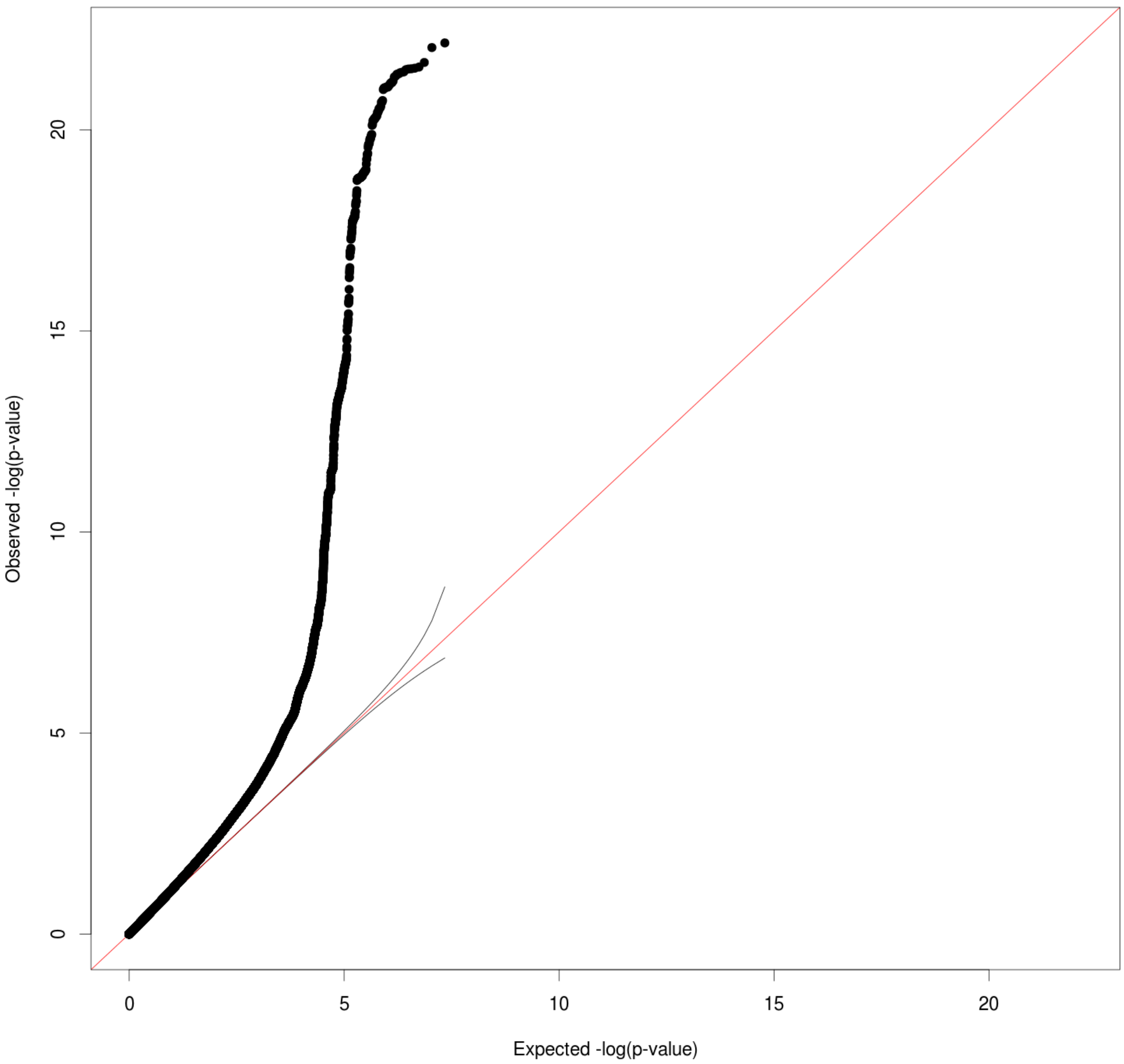

**Supplementary Figure 2.** Manhattan plot of genome-wide meta-analysis of unclassified heart failure.

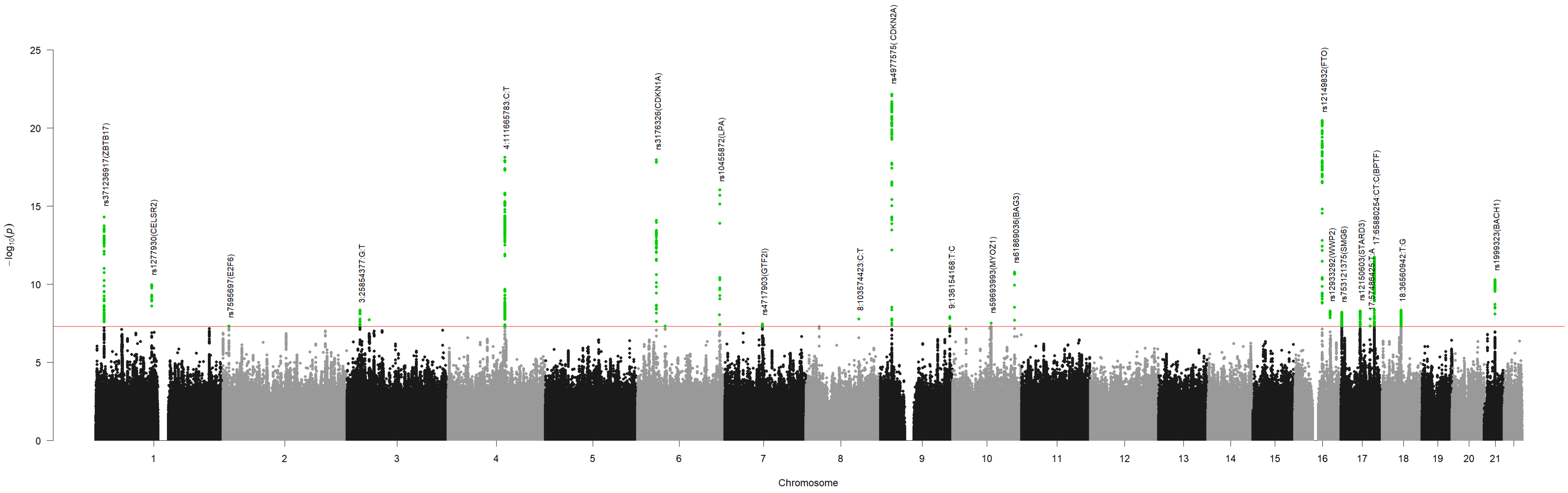

**Supplementary Figure 3A-T**. Genome-wide significant loci (n=20) associated with heart failure.

3A

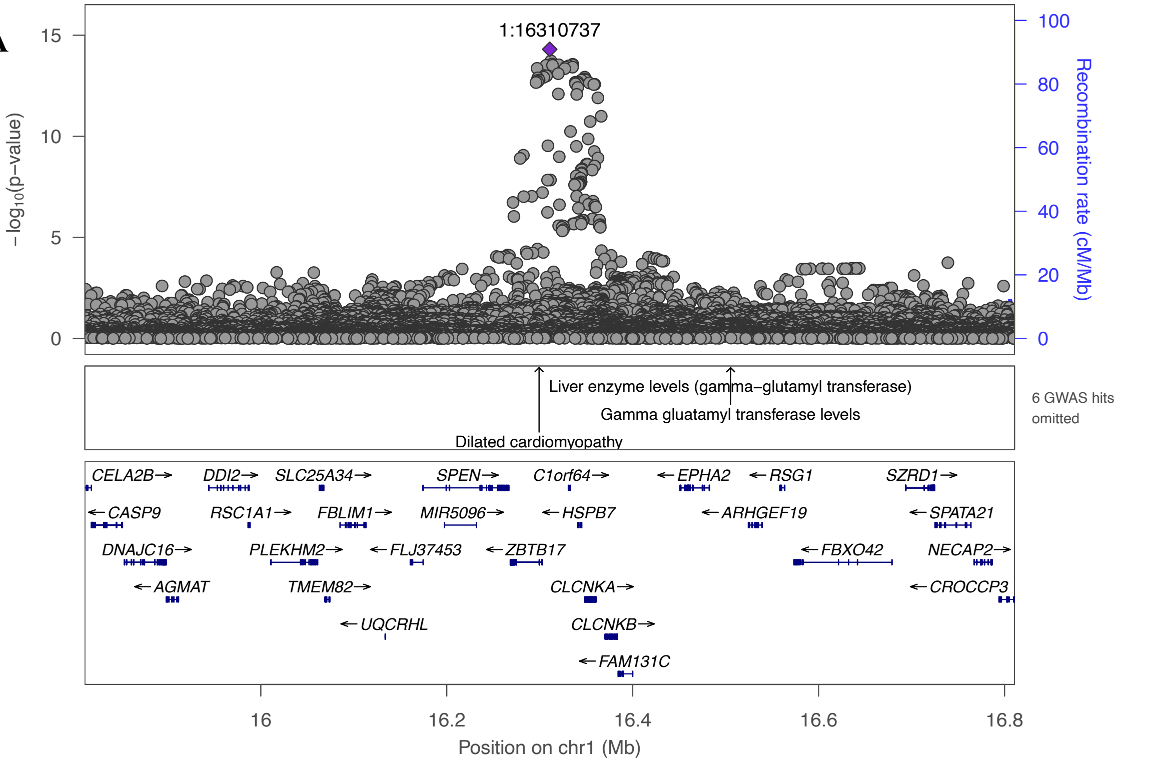


3B

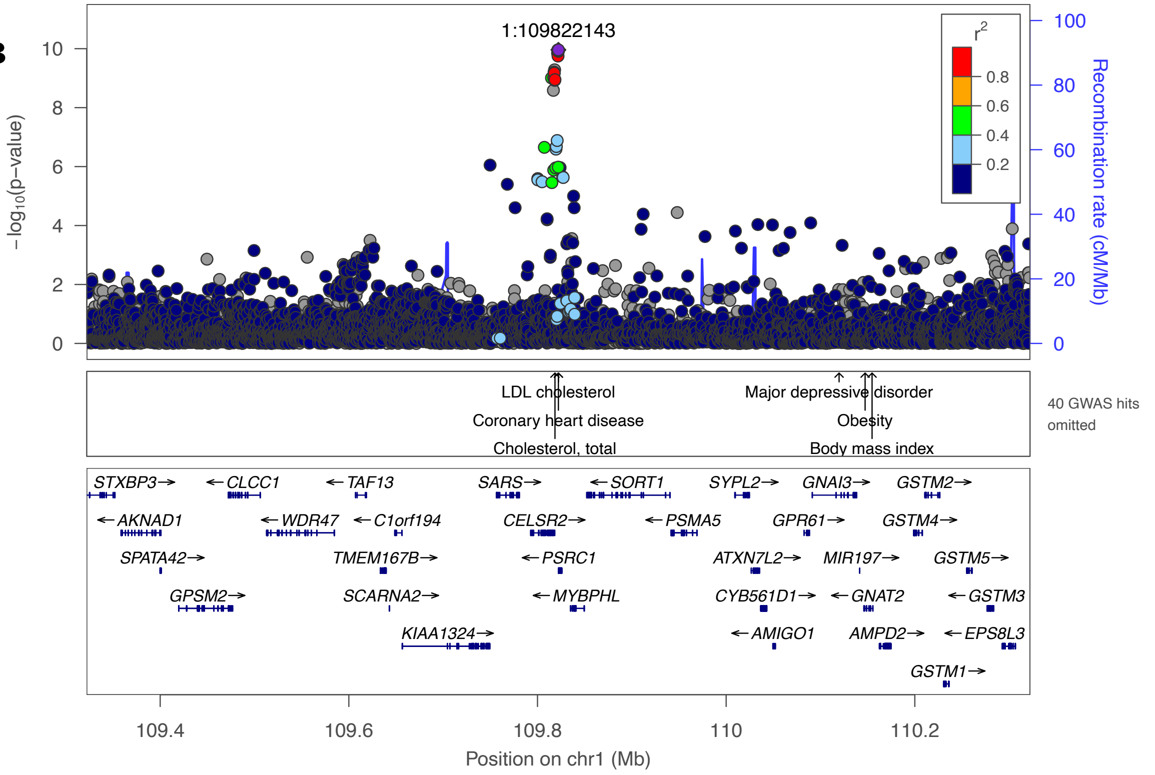

3C

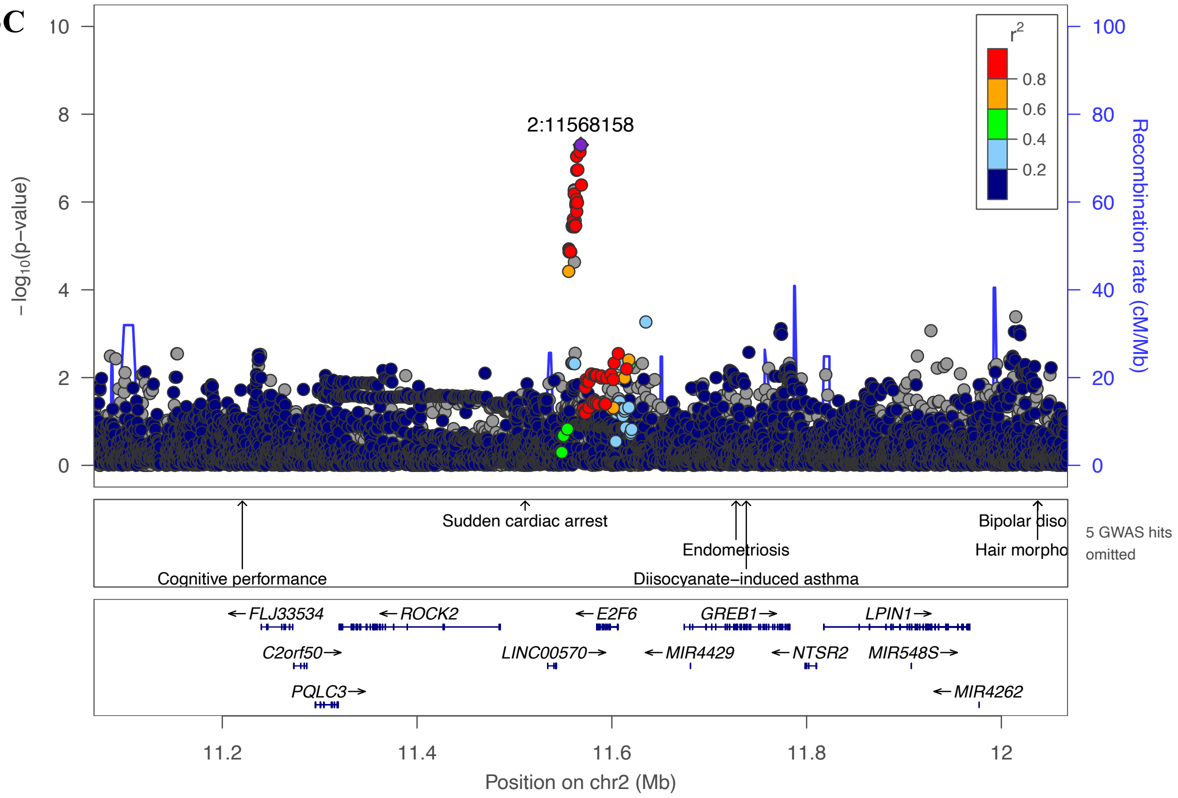

2:11568158
−log10(p−value)
Recombination rate (cM/Mb)
r2
0.8
0.6
0.4
0.2
Cognitive performance
Sudden cardiac arrest
Endometriosis
Diisocyanate−induced asthma
Bipolar diso
Hair morpho
5 GWAS hits omitted
FLJ33534
ROCK2
E2F6
GREB1
LPIN1
C2orf50
LINC00570
MIR4429
NTSR2
MIR548S
PQLC3
MIR4262
11.2
11.4
11.6
11.8
12
Position on chr2 (Mb)


3D

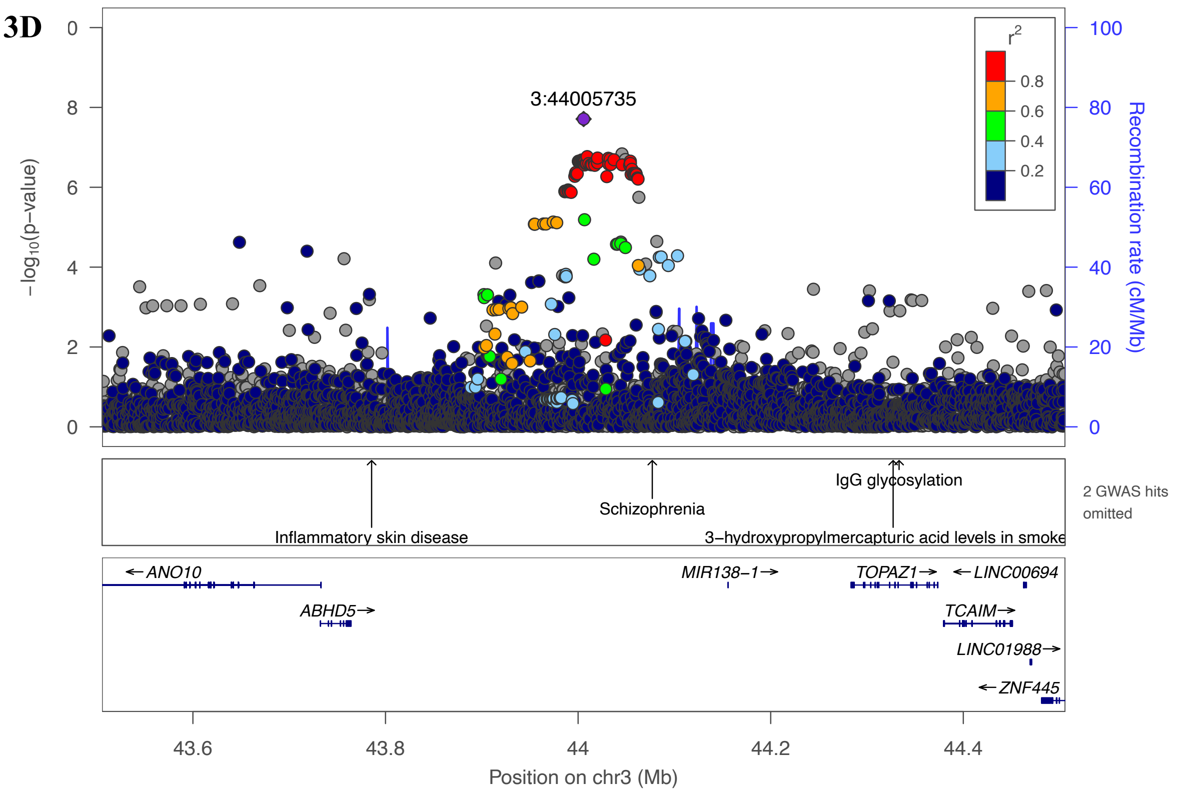

3:44005735
−log10(p−value)
Recombination rate (cM/Mb)
r2
0.8
0.6
0.4
0.2
Inflammatory skin disease
Schizophrenia
IgG glycosylation
3−hydroxypropylmercapturic acid levels in smoke
2 GWAS hits omitted
ANO10
ABHD5
MIR138−1
TOPAZ1
LINC00694
TCAIM
LINC01988
ZNF445
43.6
43.8
44
44.2
44.4
Position on chr3 (Mb)

3E

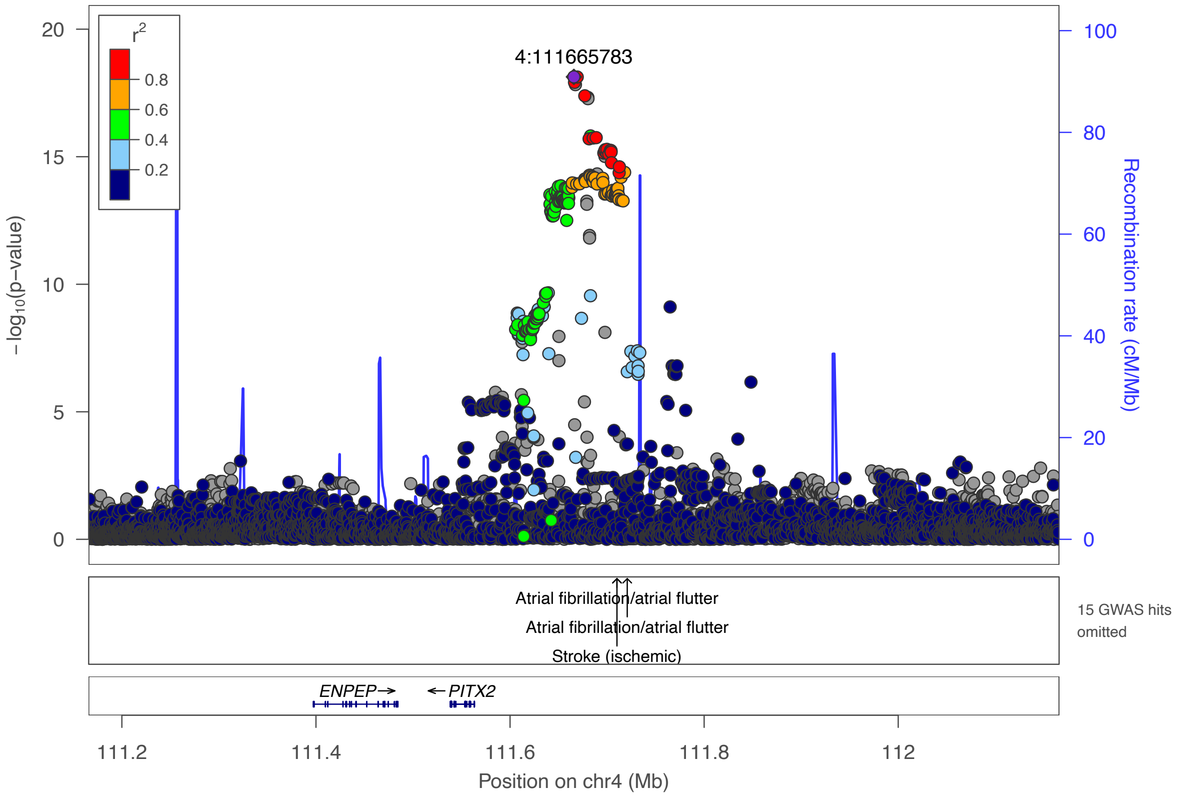

r2
0.8
0.6
0.4
0.2
4:111665783
−log10(p−value)
Recombination rate (cM/Mb)
Atrial fibrillation/atrial flutter
Atrial fibrillation/atrial flutter
Stroke (ischemic)
15 GWAS hits omitted
ENPEP→
←PITX2
111.2
111.4
111.6
111.8
112
Position on chr4 (Mb)


3F

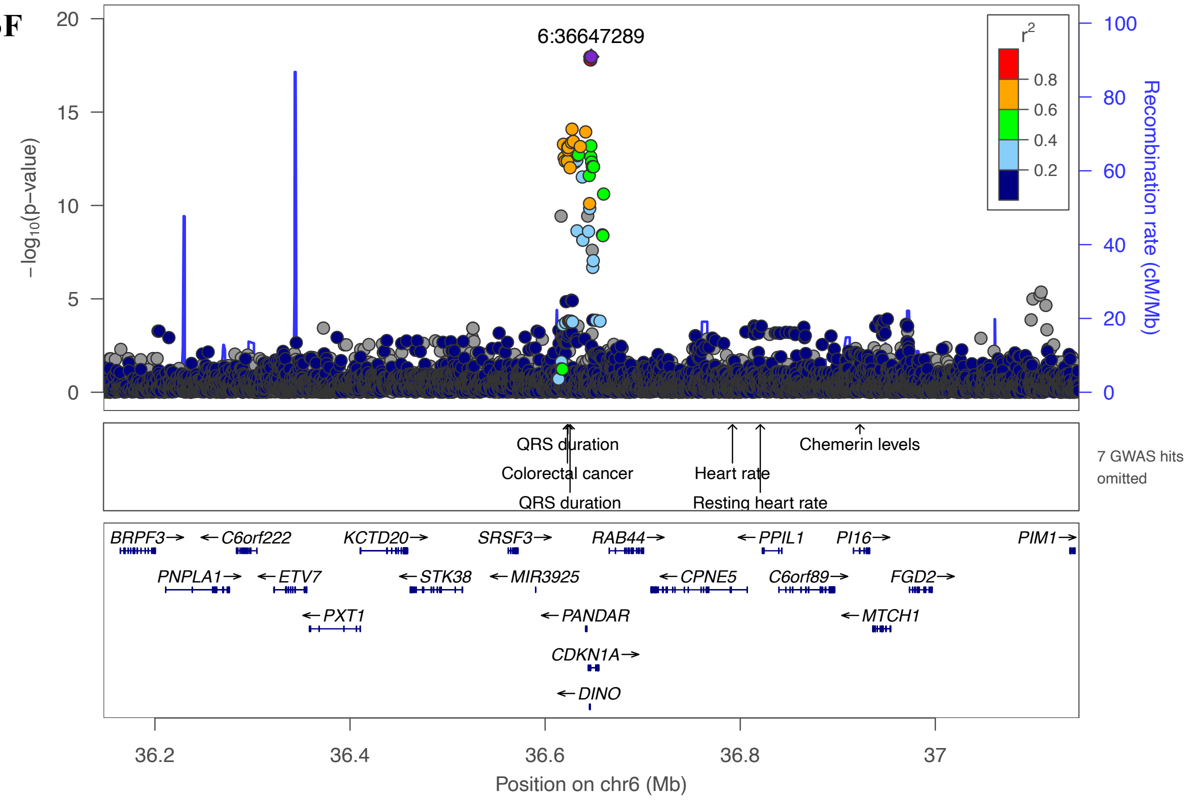

6:36647289
r2
0.8
0.6
0.4
0.2
−log10(p−value)
Recombination rate (cM/Mb)
QRS duration
Colorectal cancer
QRS duration
Heart rate
Resting heart rate
Chemerin levels
7 GWAS hits omitted
BRPF3→
←C6orf222
KCTD20→
SRSF3→
RAB44→
←PPIL1
PI16→
PIM1→
PNPLA1→
←ETV7
←STK38
←MIR3925
←CPNE5
C6orf89→
FGD2→
←PXT1
←PANDAR
←MTCH1
CDKN1A→
←DINO
36.2
36.4
36.6
36.8
37
Position on chr6 (Mb)

3G

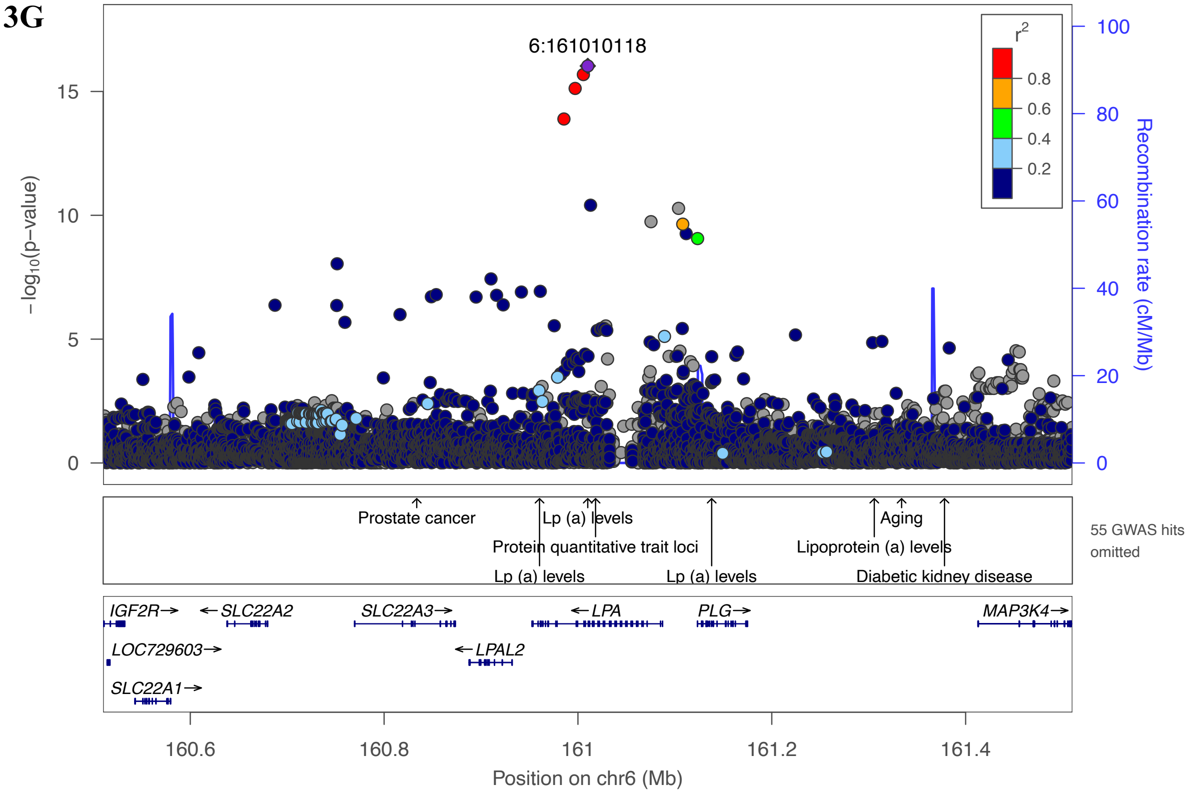
6:161010118
−log10(p−value)
Recombination rate (cM/Mb)
r2
0.8
0.6
0.4
0.2
Prostate cancer
Lp (a) levels
Protein quantitative trait loci
Lp (a) levels
Lp (a) levels
Aging
Lipoprotein (a) levels
Diabetic kidney disease
55 GWAS hits omitted
IGF2R
SLC22A2
SLC22A3
LPA
PLG
MAP3K4
LOC729603
LPAL2
SLC22A1
160.6
160.8
161
161.2
161.4
Position on chr6 (Mb)

3H

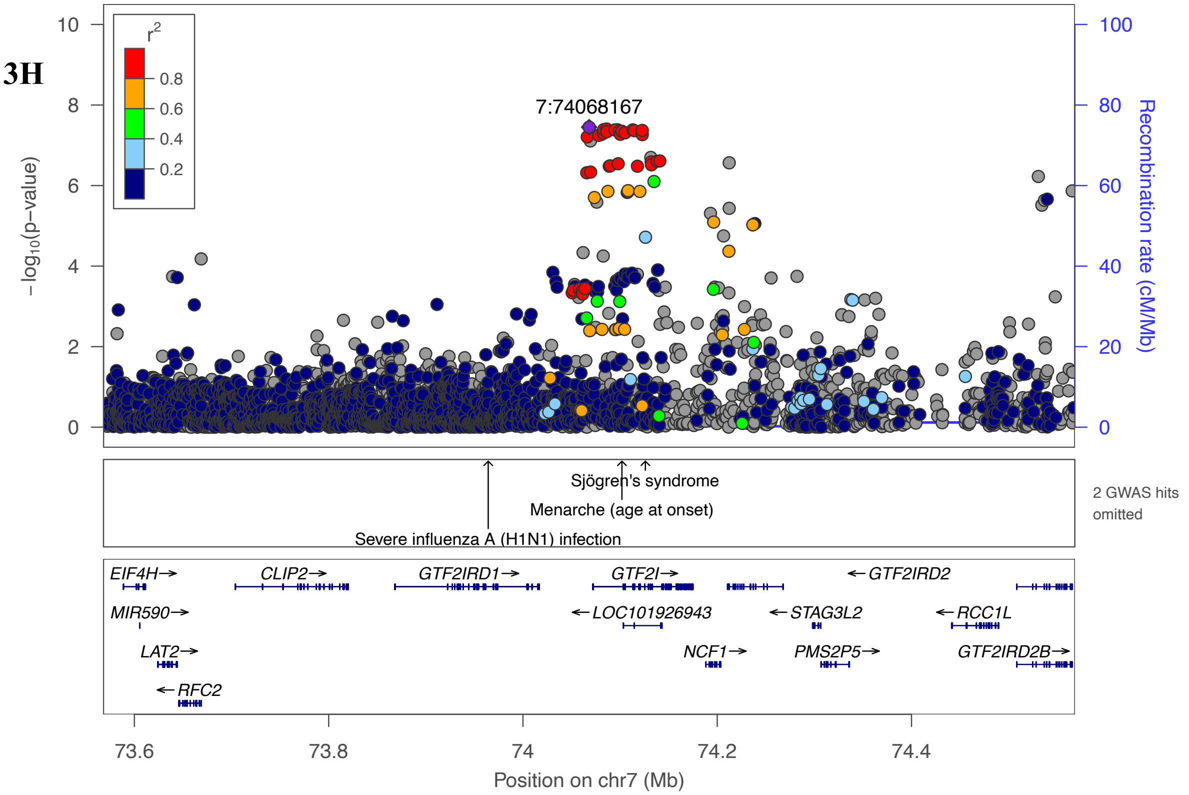
7:74068167
−log10(p−value)
Recombination rate (cM/Mb)
r2
0.8
0.6
0.4
0.2
Sjögren's syndrome
Menarche (age at onset)
Severe influenza A (H1N1) infection
2 GWAS hits omitted
EIF4H
CLIP2
GTF2IRD1
GTF2I
GTF2IRD2
MIR590
LOC101926943
STAG3L2
RCC1L
LAT2
NCF1
PMS2P5
GTF2IRD2B
RFC2
73.6
73.8
74
74.2
74.4
Position on chr7 (Mb)

3I

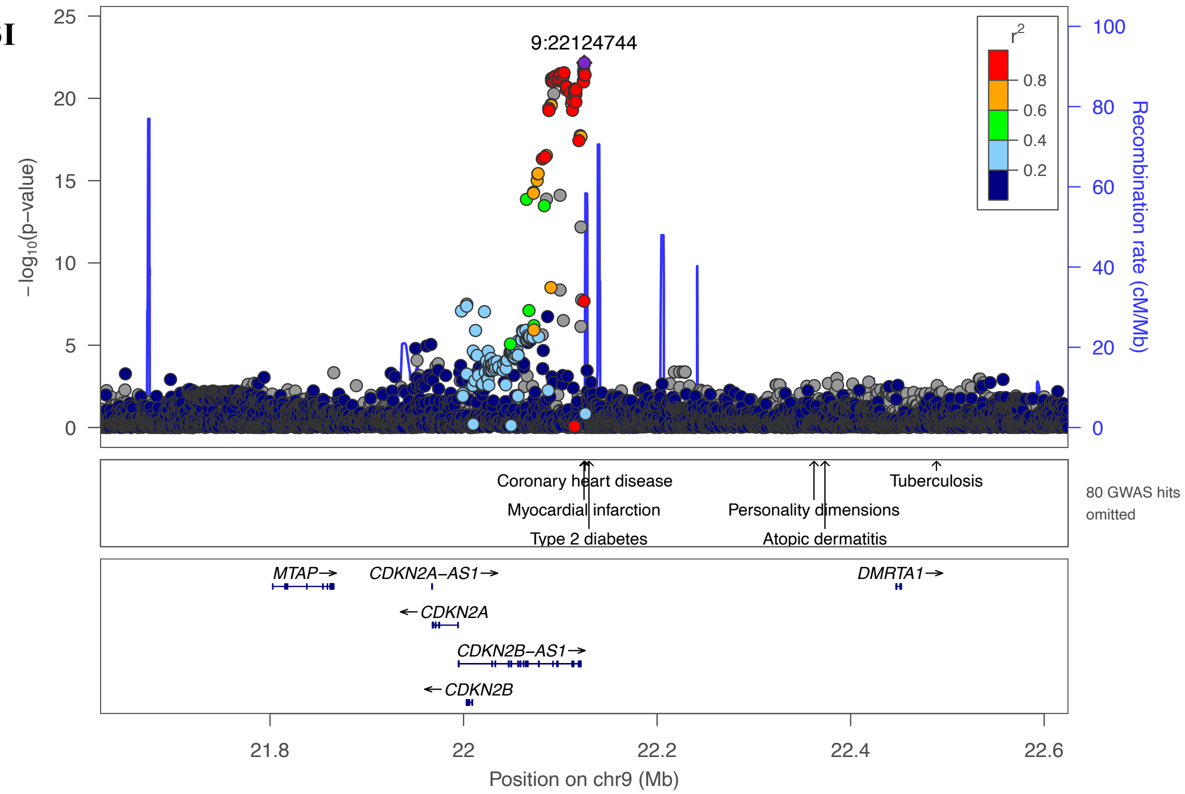
9:22124744
r²
0.8
0.6
0.4
0.2
−log10(p−value)
Recombination rate (cM/Mb)
Coronary heart disease
Myocardial infarction
Type 2 diabetes
Personality dimensions
Atopic dermatitis
Tuberculosis
80 GWAS hits omitted
MTAP
CDKN2A−AS1
CDKN2A
CDKN2B−AS1
CDKN2B
DMRTA1
Position on chr9 (Mb)

3J

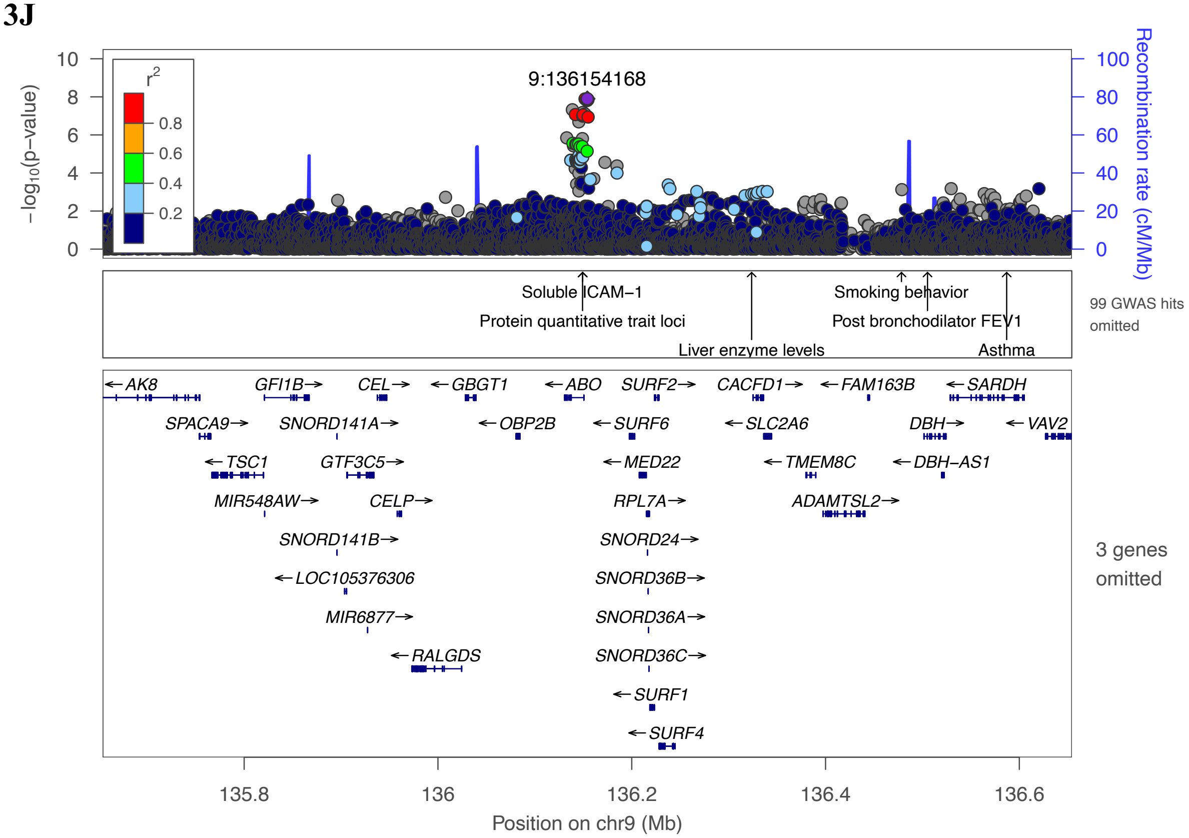
9:136154168
r²
0.8
0.6
0.4
0.2
−log10(p−value)
Recombination rate (cM/Mb)
Soluble ICAM−1
Protein quantitative trait loci
Liver enzyme levels
Smoking behavior
Post bronchodilator FEV1
Asthma
99 GWAS hits omitted
AK8
GFI1B
CEL
GBGT1
ABO
SURF2
CACFD1
FAM163B
SARDH
SPACA9
SNORD141A
OBP2B
SURF6
SLC2A6
DBH
VAV2
TSC1
GTF3C5
MED22
TMEM8C
DBH−AS1
MIR548AW
CELP
RPL7A
ADAMTSL2
SNORD141B
SNORD24
LOC105376306
SNORD36B
MIR6877
SNORD36A
RALGDS
SNORD36C
SURF1
SURF4
3 genes omitted
Position on chr9 (Mb)

3K

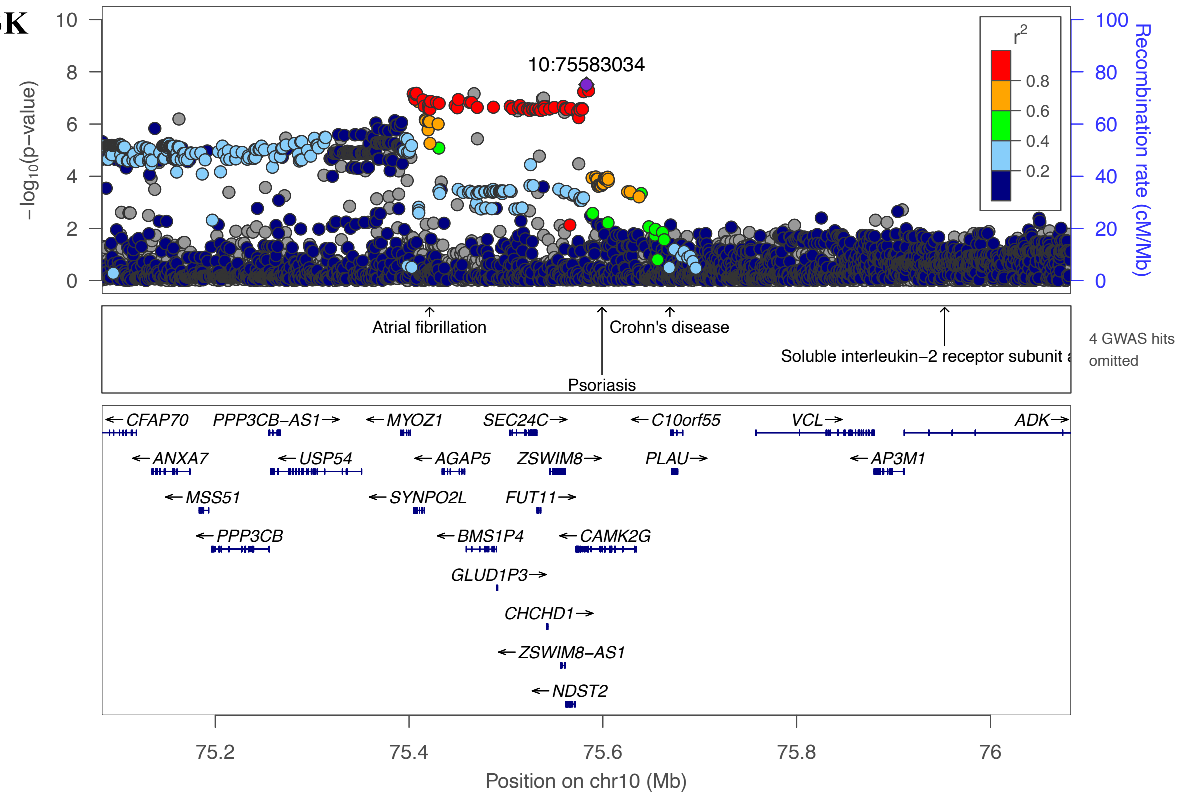

10:75583034
r2
0.8
0.6
0.4
0.2
− log10(p−value)
Recombination rate (cM/Mb)
Atrial fibrillation
Crohn's disease
Soluble interleukin−2 receptor subunit a
Psoriasis
4 GWAS hits omitted
CFAP70
PPP3CB−AS1
MYOZ1
SEC24C
C10orf55
VCL
ADK
ANXA7
USP54
AGAP5
ZSWIM8
PLAU
AP3M1
MSS51
SYNPO2L
FUT11
PPP3CB
BMS1P4
CAMK2G
GLUD1P3
CHCHD1
ZSWIM8−AS1
NDST2
Position on chr10 (Mb)


3L

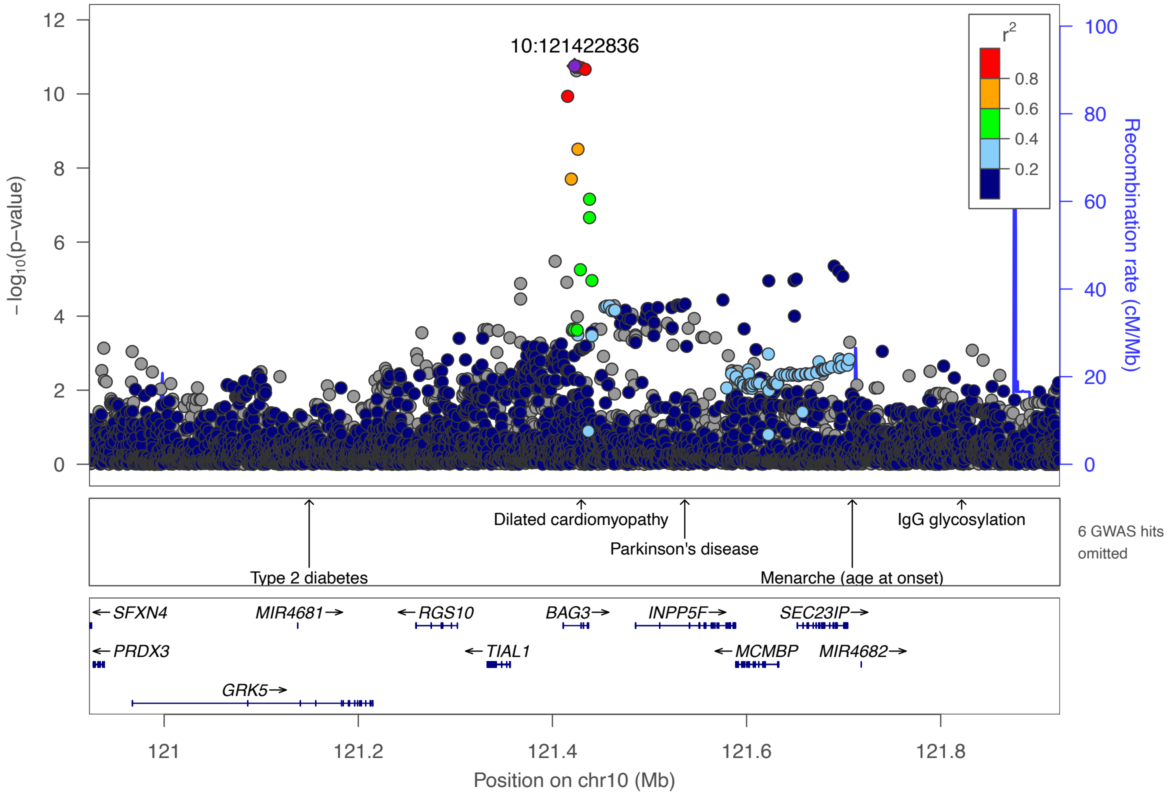

10:121422836
r2
0.8
0.6
0.4
0.2
− log10(p−value)
Recombination rate (cM/Mb)
Dilated cardiomyopathy
IgG glycosylation
Parkinson's disease
Type 2 diabetes
Menarche (age at onset)
6 GWAS hits omitted
SFXN4
MIR4681
RGS10
BAG3
INPP5F
SEC23IP
PRDX3
TIAL1
MCMBP
MIR4682
GRK5
Position on chr10 (Mb)

3M

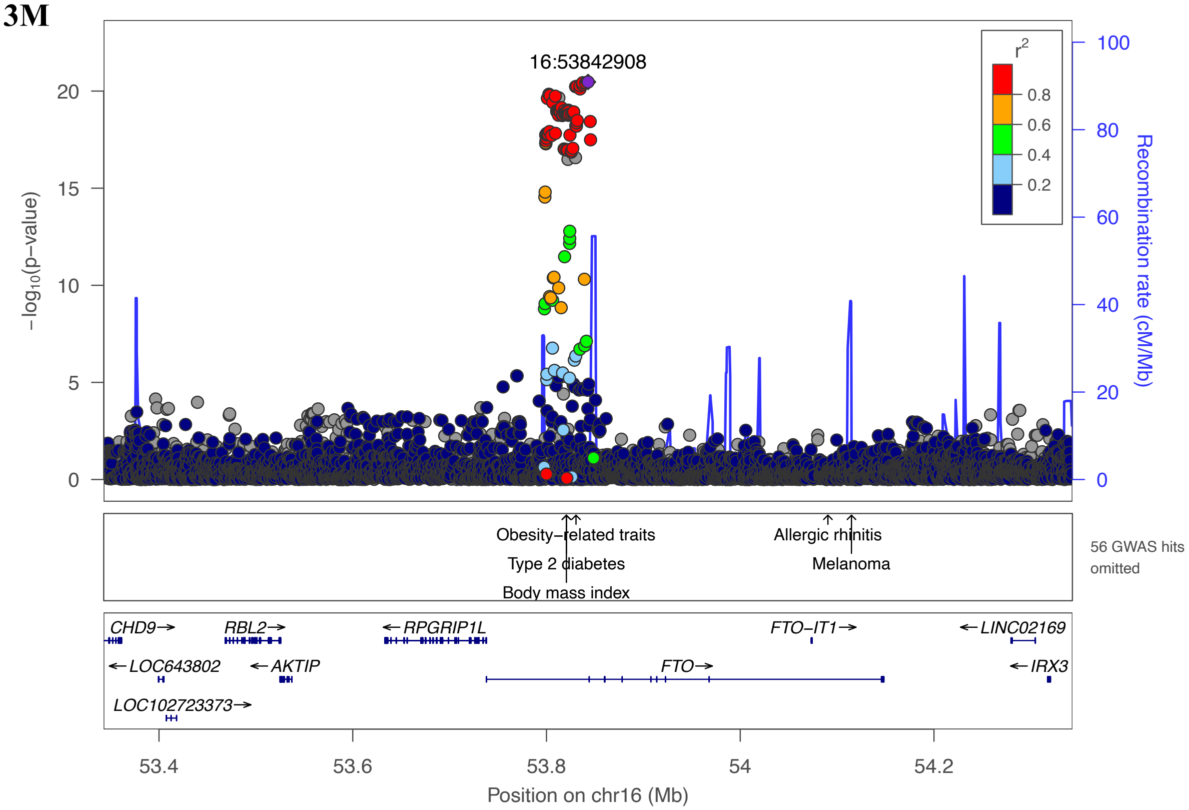
16:53842908
r2
0.8
0.6
0.4
0.2
−log10(p−value)
Recombination rate (cM/Mb)
Obesity−related traits
Type 2 diabetes
Body mass index
Allergic rhinitis
Melanoma
56 GWAS hits omitted
CHD9
RBL2
RPGRIP1L
FTO−IT1
LINC02169
LOC643802
AKTIP
FTO
IRX3
LOC102723373
Position on chr16 (Mb)

3N

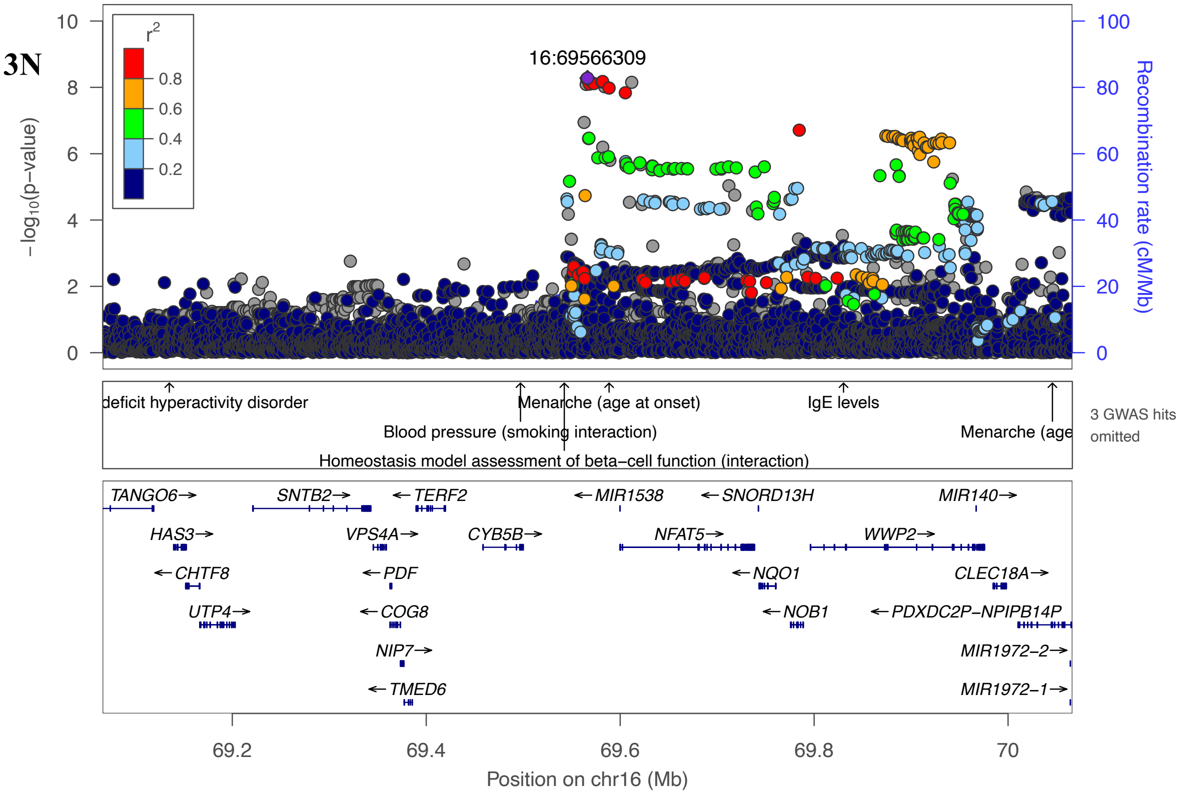
16:69566309
r2
0.8
0.6
0.4
0.2
−log10(p−value)
Recombination rate (cM/Mb)
deficit hyperactivity disorder
Menarche (age at onset)
IgE levels
Blood pressure (smoking interaction)
Menarche (age
Homeostasis model assessment of beta−cell function (interaction)
3 GWAS hits omitted
TANGO6
SNTB2
TERF2
MIR1538
SNORD13H
MIR140
HAS3
VPS4A
CYB5B
NFAT5
WWP2
CHTF8
PDF
NQO1
CLEC18A
UTP4
COG8
NOB1
PDXDC2P−NPIPB14P
NIP7
MIR1972−2
TMED6
MIR1972−1
Position on chr16 (Mb)

3O

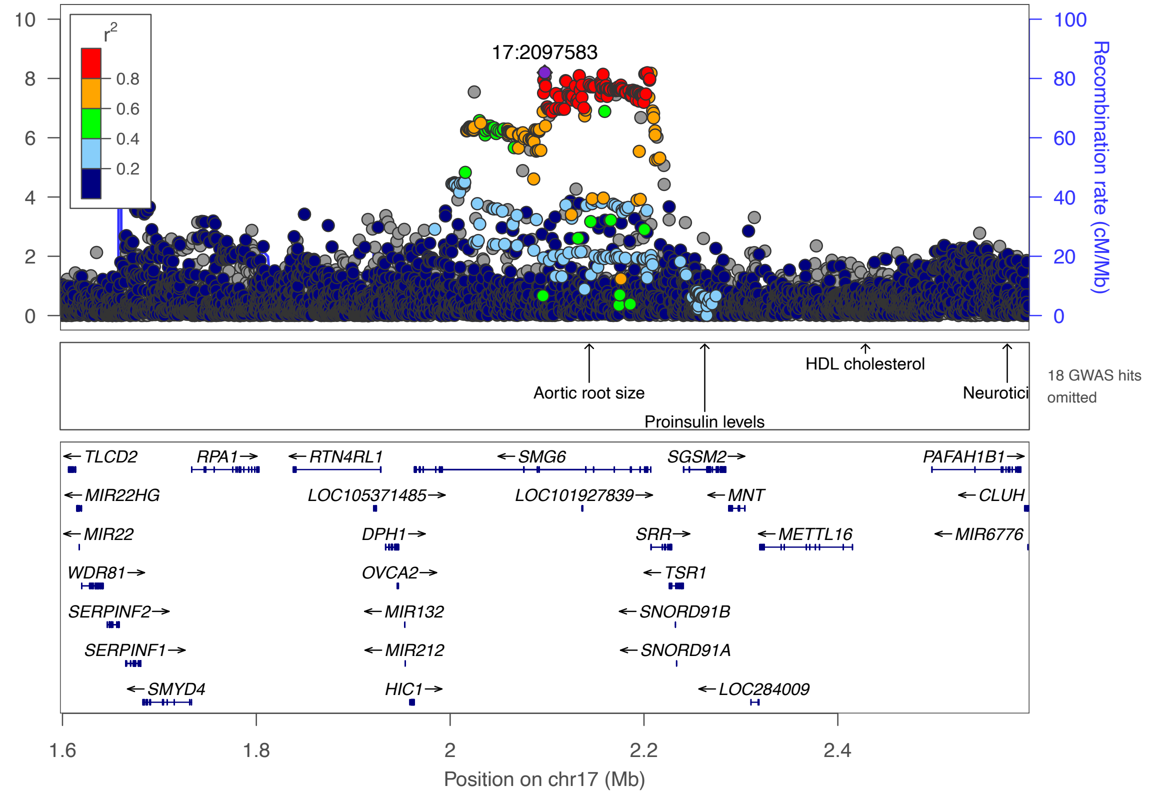

17:2097583
r2
0.8
0.6
0.4
0.2
−log10(p−value)
Recombination rate (cM/Mb)
Aortic root size
Proinsulin levels
HDL cholesterol
Neurotici
18 GWAS hits omitted
TLCD2
RPA1
RTN4RL1
SMG6
SGSM2
PAFAH1B1
MIR22HG
LOC105371485
LOC101927839
MNT
CLUH
MIR22
DPH1
SRR
METTL16
MIR6776
WDR81
OVCA2
TSR1
SERPINF2
MIR132
SNORD91B
SERPINF1
MIR212
SNORD91A
SMYD4
HIC1
LOC284009
Position on chr17 (Mb)


3P

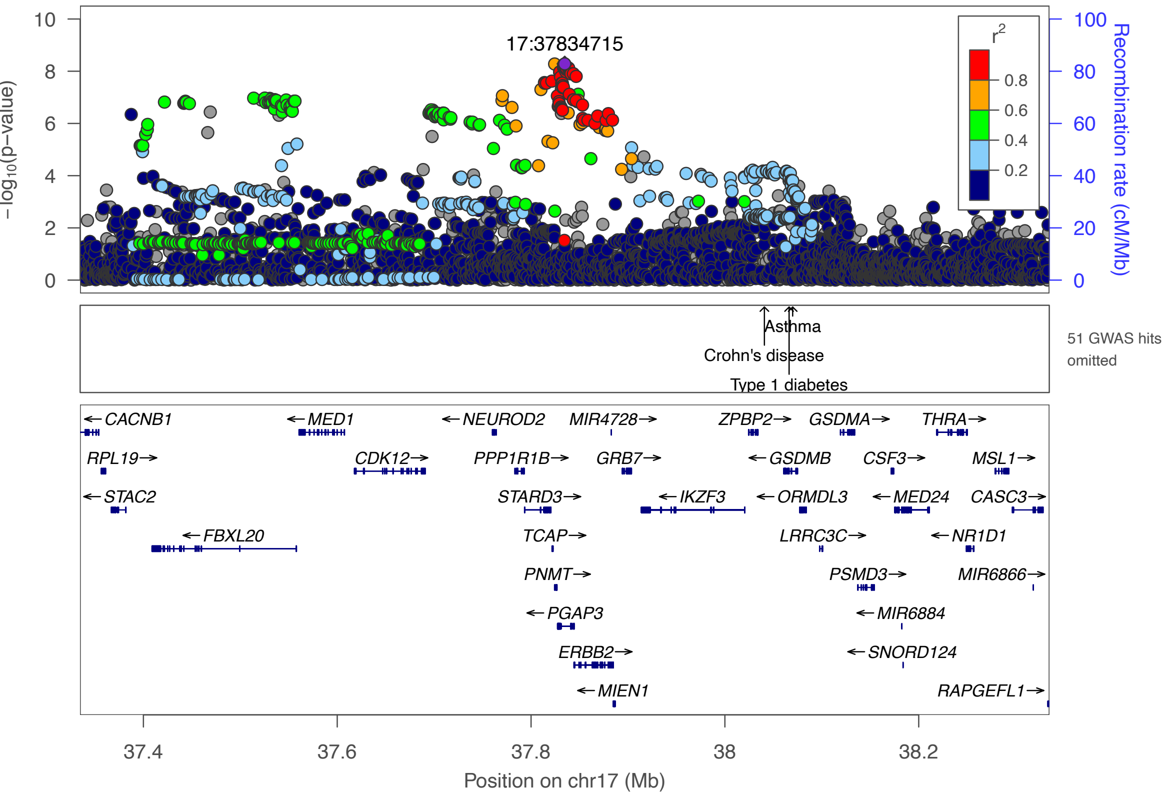

17:37834715
r2
0.8
0.6
0.4
0.2
−log10(p−value)
Recombination rate (cM/Mb)
Asthma
Crohn's disease
Type 1 diabetes
51 GWAS hits omitted
CACNB1
MED1
NEUROD2
MIR4728
ZPBP2
GSDMA
THRA
RPL19
CDK12
PPP1R1B
GRB7
GSDMB
CSF3
MSL1
STAC2
STARD3
IKZF3
ORMDL3
MED24
CASC3
FBXL20
TCAP
LRRC3C
NR1D1
PNMT
PSMD3
MIR6866
PGAP3
MIR6884
ERBB2
SNORD124
MIEN1
RAPGEFL1
Position on chr17 (Mb)

**3Q**

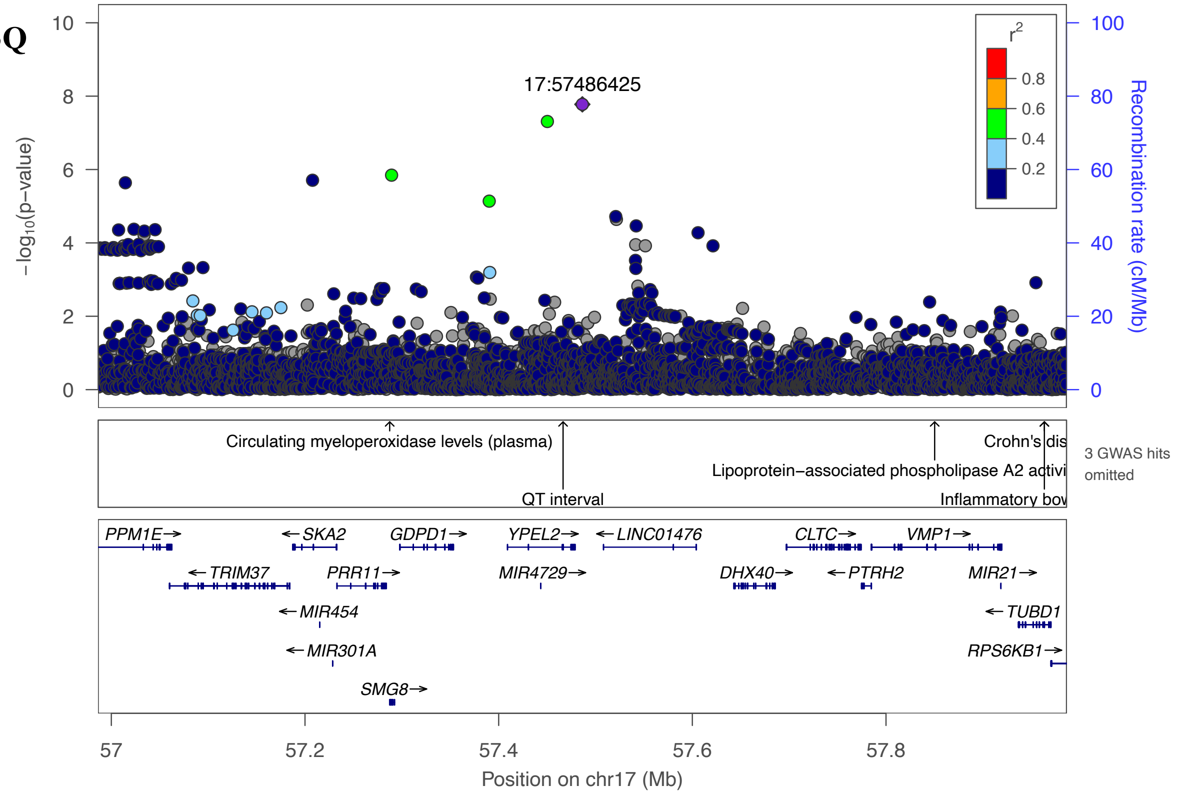
17:57486425
−log10(p−value)
Recombination rate (cM/Mb)
r2
0.8
0.6
0.4
0.2
Circulating myeloperoxidase levels (plasma)
QT interval
Lipoprotein−associated phospholipase A2 activi
Crohn's dis
Inflammatory bov
3 GWAS hits omitted
PPM1E
TRIM37
SKA2
MIR454
MIR301A
PRR11
GDPD1
SMG8
YPEL2
MIR4729
LINC01476
DHX40
CLTC
PTRH2
VMP1
MIR21
TUBD1
RPS6KB1
Position on chr17 (Mb)

**3R**

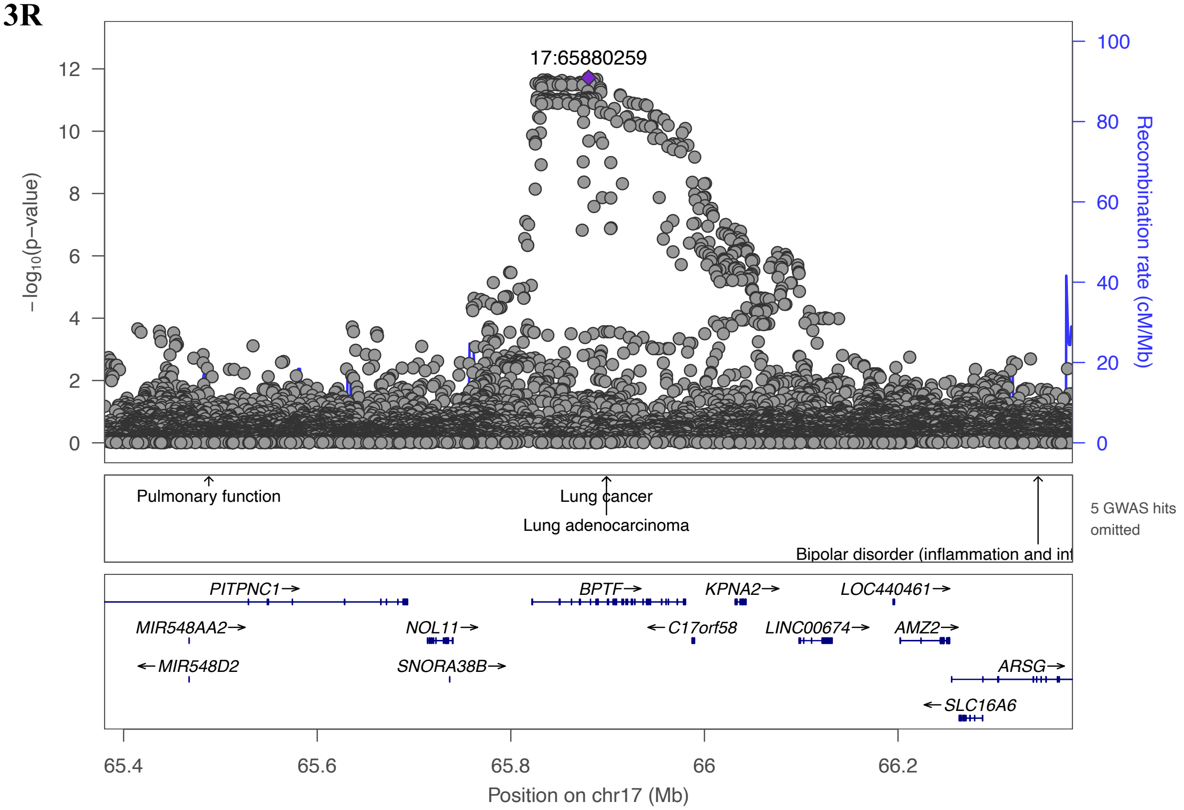
17:65880259
−log10(p−value)
Recombination rate (cM/Mb)
Pulmonary function
Lung cancer
Lung adenocarcinoma
Bipolar disorder (inflammation and inf
5 GWAS hits omitted
PITPNC1
MIR548AA2
MIR548D2
NOL11
SNORA38B
BPTF
C17orf58
KPNA2
LINC00674
LOC440461
AMZ2
ARSG
SLC16A6
Position on chr17 (Mb)

**3S**

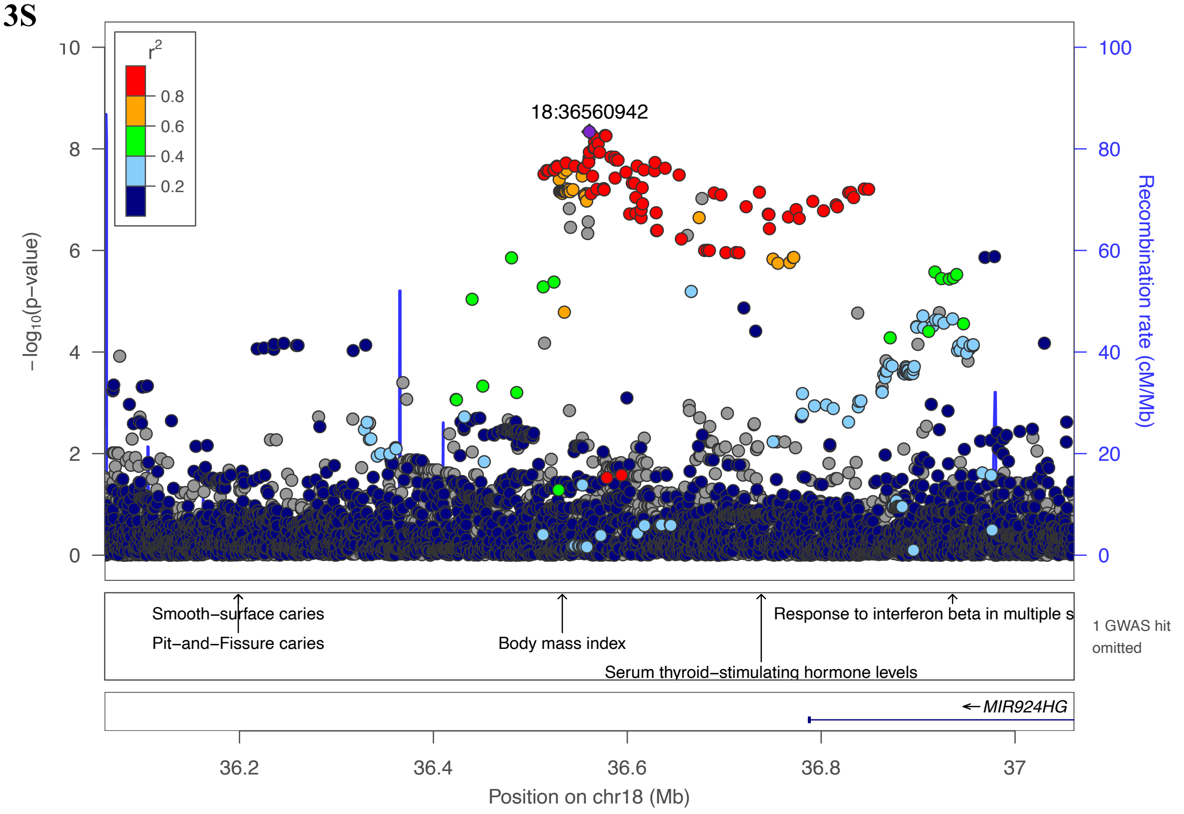
18:36560942
r2
0.8
0.6
0.4
0.2
−log10(p−value)
Recombination rate (cM/Mb)
Smooth−surface caries
Pit−and−Fissure caries
Body mass index
Response to interferon beta in multiple s
Serum thyroid−stimulating hormone levels
1 GWAS hit omitted
MIR924HG
Position on chr18 (Mb)

**3T**

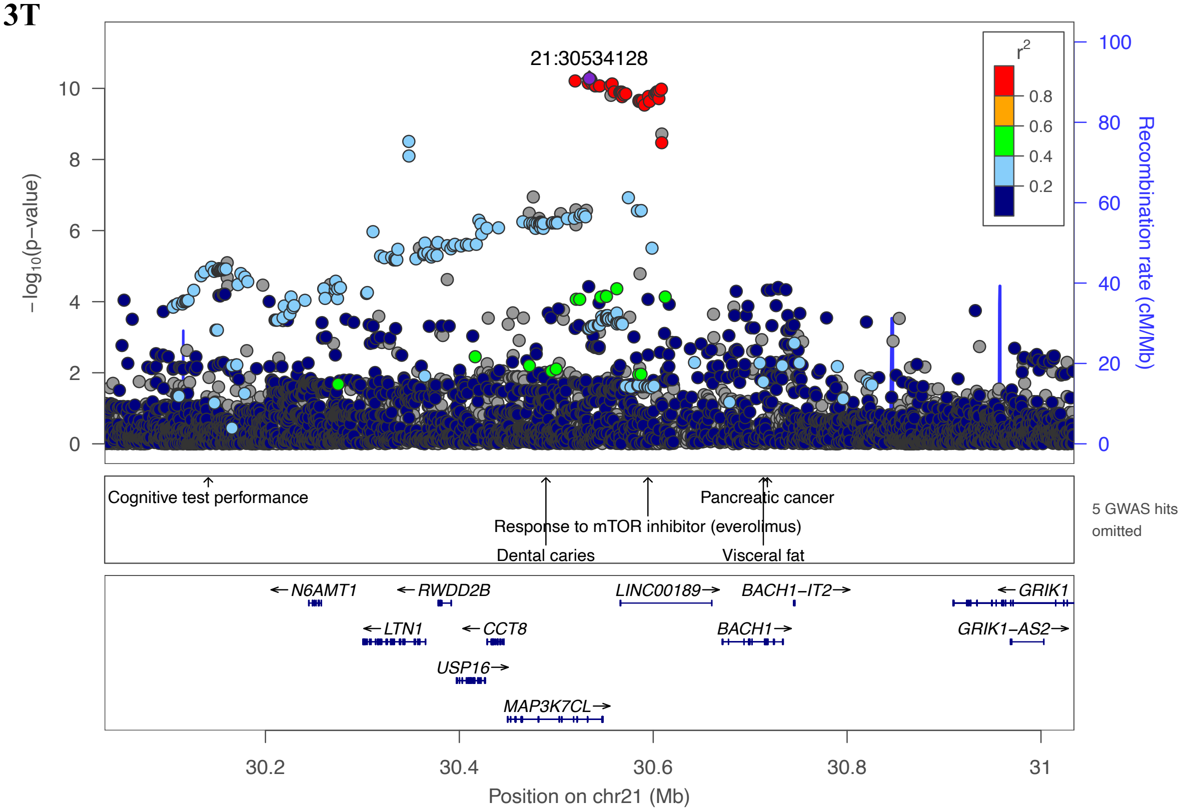
21:30534128
r2
0.8
0.6
0.4
0.2
−log10(p−value)
Recombination rate (cM/Mb)
Cognitive test performance
Pancreatic cancer
Response to mTOR inhibitor (everolimus)
Dental caries
Visceral fat
5 GWAS hits omitted
N6AMT1
RWDD2B
LINC00189
BACH1−IT2
GRIK1
LTN1
CCT8
BACH1
GRIK1−AS2
USP16
MAP3K7CL
Position on chr21 (Mb)

**Supplementary Figure 4.** Comparison of top HFpEF-associated between GWAS results from two HFpEF definitions - a more restrictive definition (n=12,119 cases) and the definition used for the primary HFpEF GWAS (n=19,598 cases).

Similar to the primary HFpEF GWAS, only one genome-wide significant locus (*FTO*) was identified. Using LDSC and the GWAS summary statistics, we found that the genetic correlation between the two HFpEF definitions was very high ($r=0.981$, $p<2\times10^{-16}$). Among top 110 HFpEF-associated common SNPs ($p<10^{-6}$, MAF>1%), the genetic effects between the two HFpEF GWAS were highly correlated ($r=0.995$, $p<2\times10^{-16}$). Mostly driven by a larger number of HFpEF cases in the original definition (19,598 vs. 12,119), the p-values of 109 out of 110 SNPs were lower in the original HFpEF GWAS conducted in the less restrictive cohort.

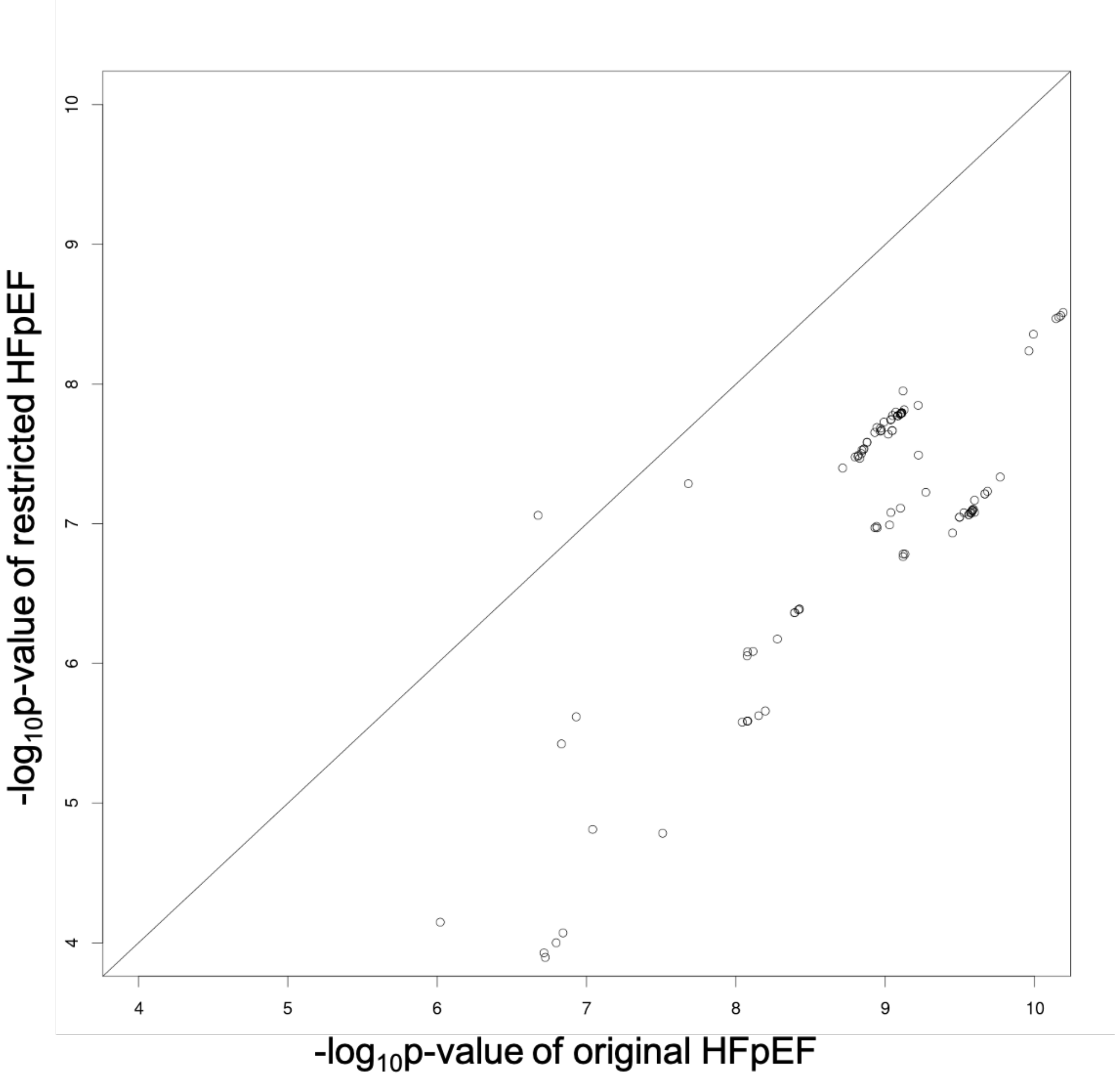

**Supplementary Figure 5A.** Quantile-quantile plot of genome-wide association study of heart failure with reduced ejection fraction (HFrEF).

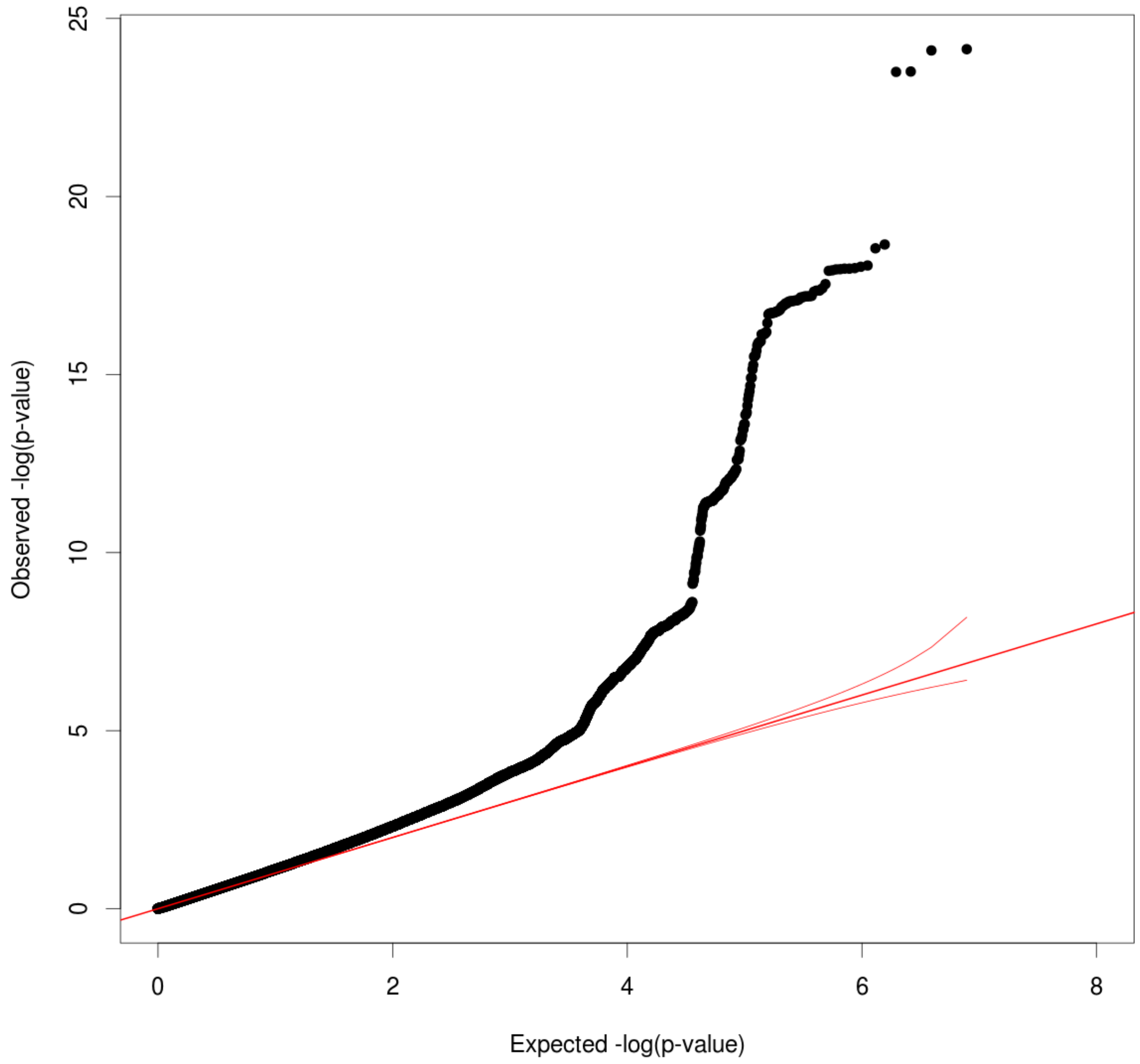

**Supplementary Figure 5B.** Quantile-quantile plot of genome-wide association study of heart failure with preserved ejection fraction (HFpEF).

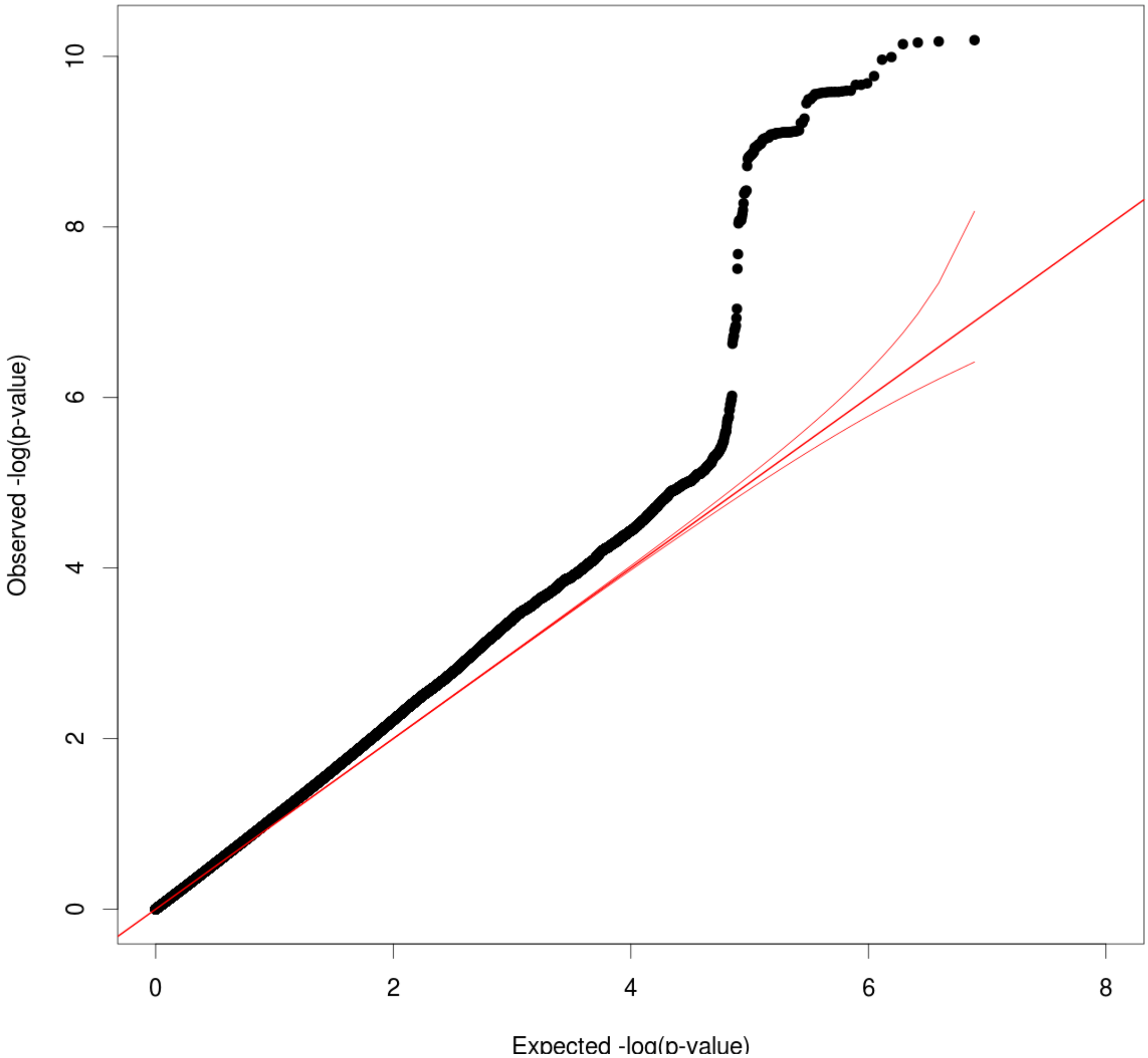

**Supplementary Figure 6 A-M**. Genome-wide significant loci associated with HFrEF/HFpEF

6A

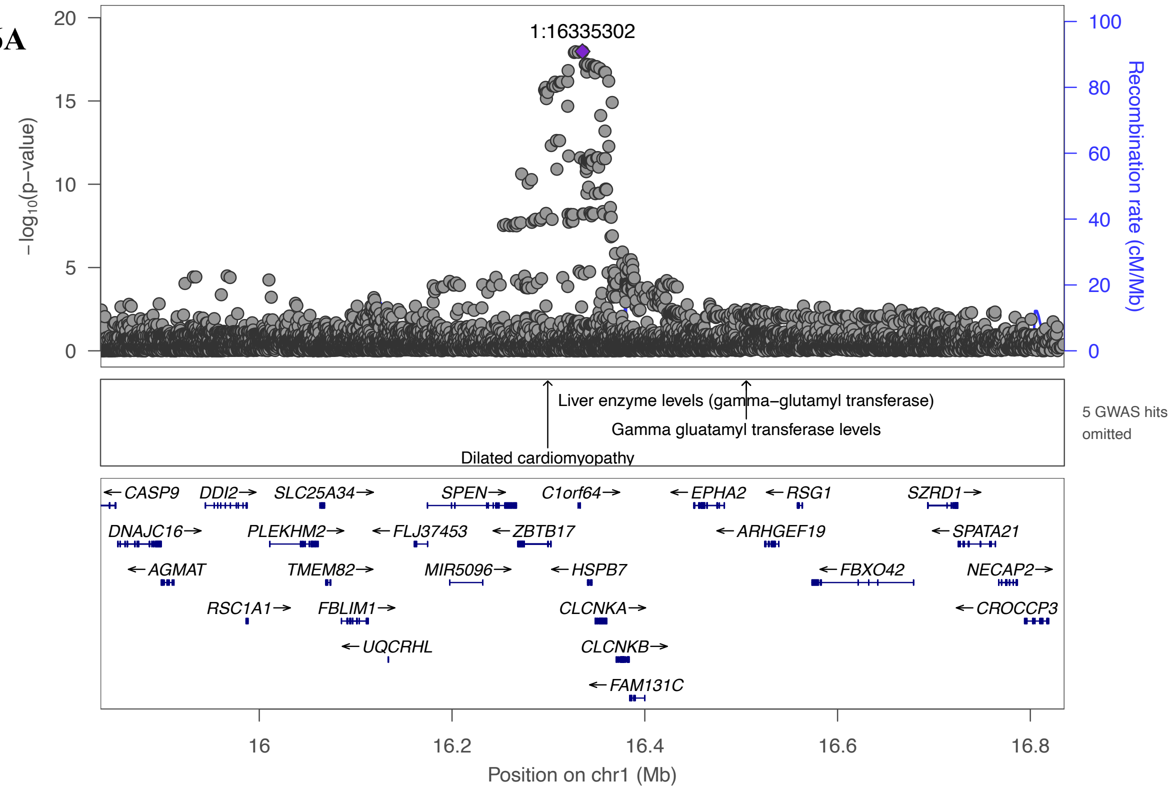


6B

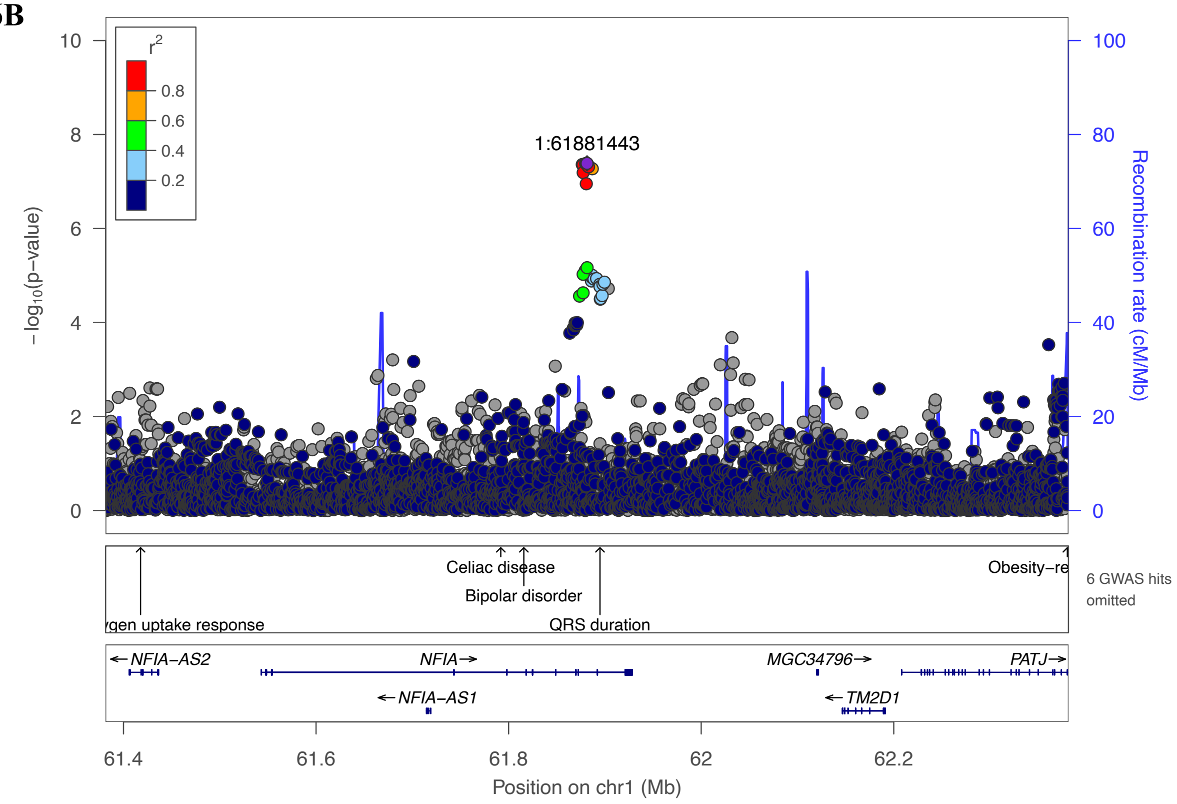

6C

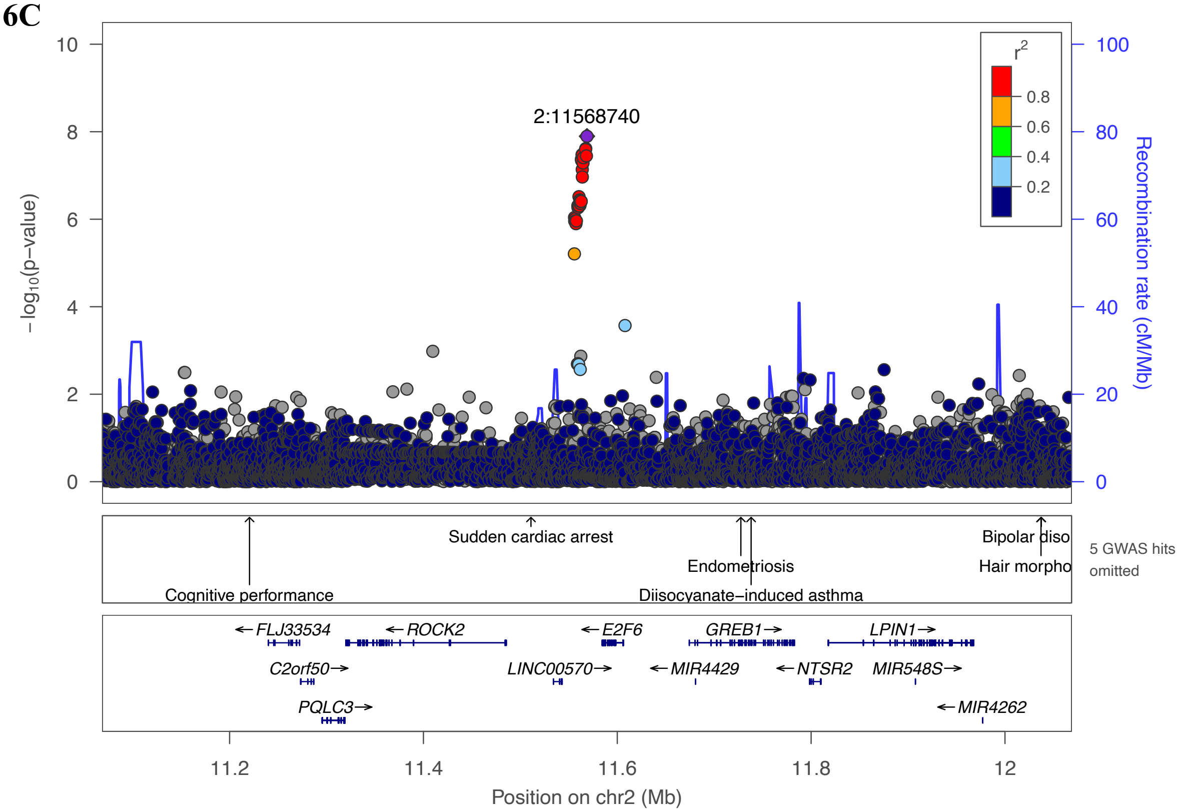

2:11568740
r2
0.8
0.6
0.4
0.2
−log10(p−value)
Recombination rate (cM/Mb)
Sudden cardiac arrest
Endometriosis
Bipolar diso
Hair morpho
Cognitive performance
Diisocyanate−induced asthma
5 GWAS hits omitted
FLJ33534
ROCK2
E2F6
GREB1
LPIN1
C2orf50
LINC00570
MIR4429
NTSR2
MIR548S
PQLC3
MIR4262
Position on chr2 (Mb)


6D

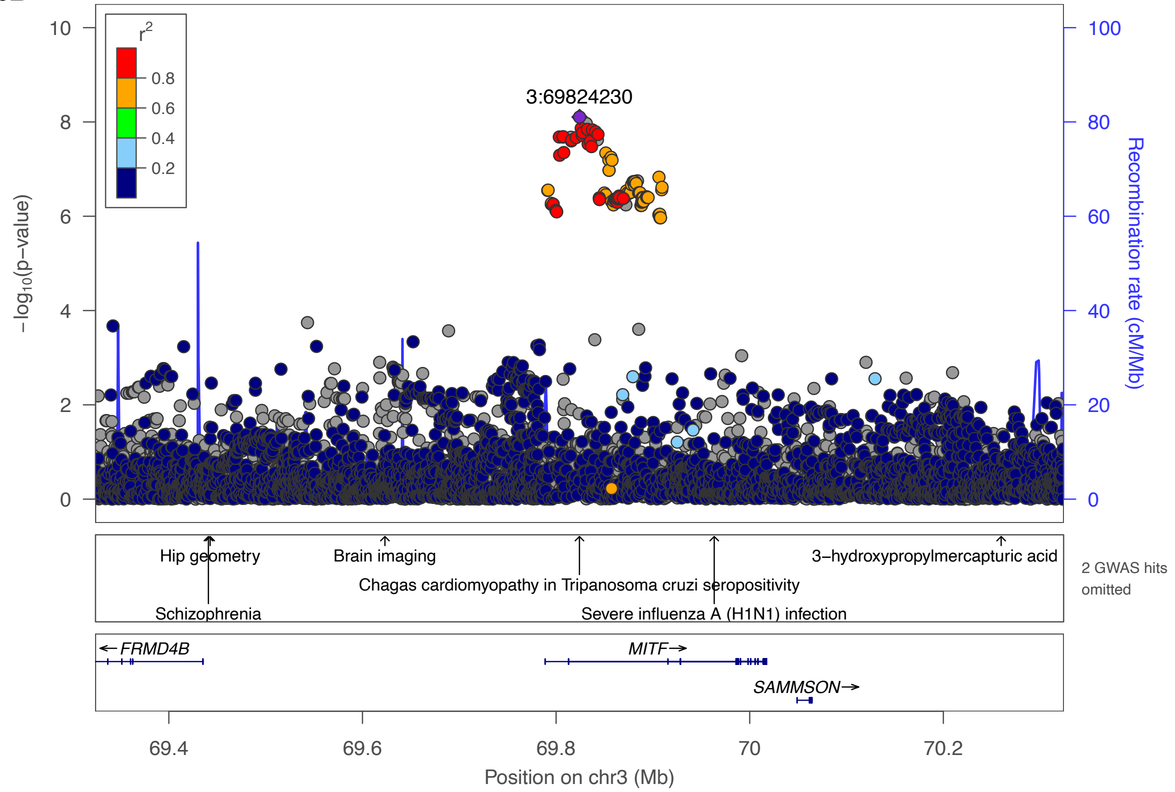

3:69824230
r2
0.8
0.6
0.4
0.2
−log10(p−value)
Recombination rate (cM/Mb)
Hip geometry
Brain imaging
3−hydroxypropylmercapturic acid
Chagas cardiomyopathy in Tripanosoma cruzi seropositivity
Schizophrenia
Severe influenza A (H1N1) infection
2 GWAS hits omitted
FRMD4B
MITF
SAMMSON
Position on chr3 (Mb)

6E

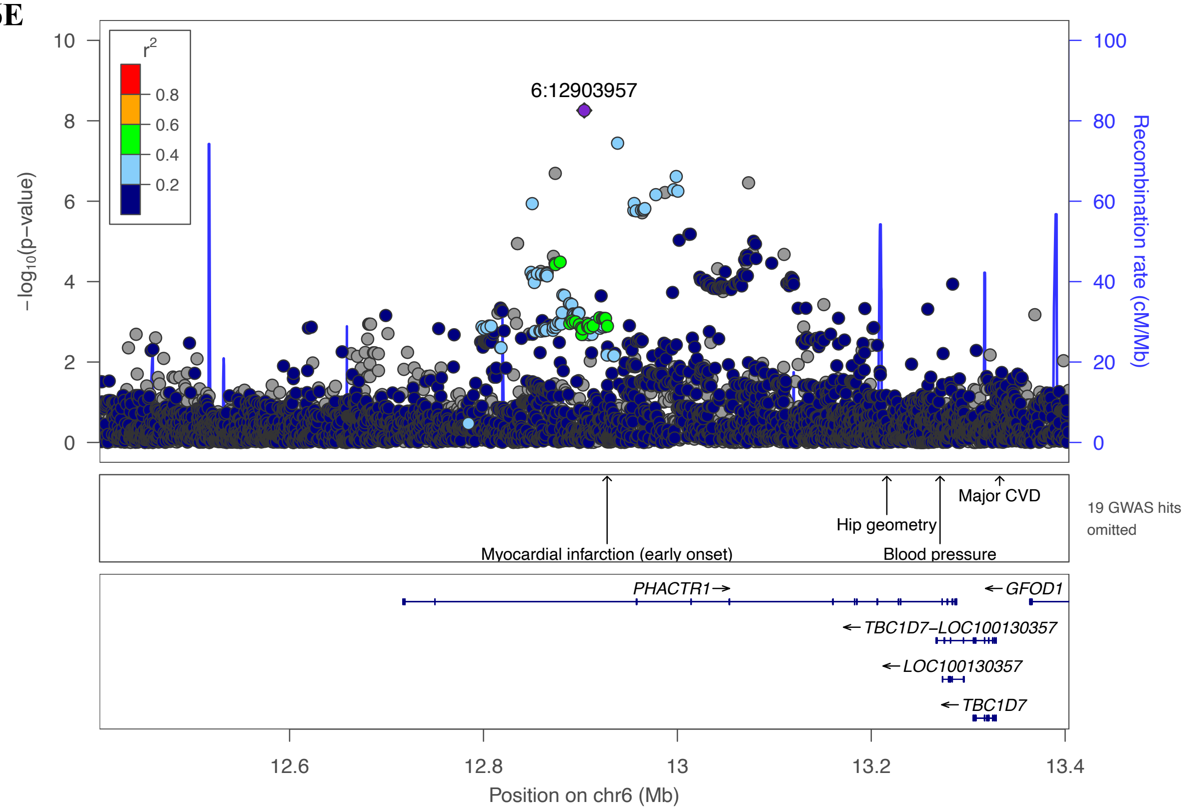
6:12903957
r2
0.8
0.6
0.4
0.2
−log10(p−value)
Recombination rate (cM/Mb)
Myocardial infarction (early onset)
Hip geometry
Blood pressure
Major CVD
19 GWAS hits omitted
PHACTR1→
←GFOD1
←TBC1D7−LOC100130357
←LOC100130357
←TBC1D7
12.6
12.8
13
13.2
13.4
Position on chr6 (Mb)

6F

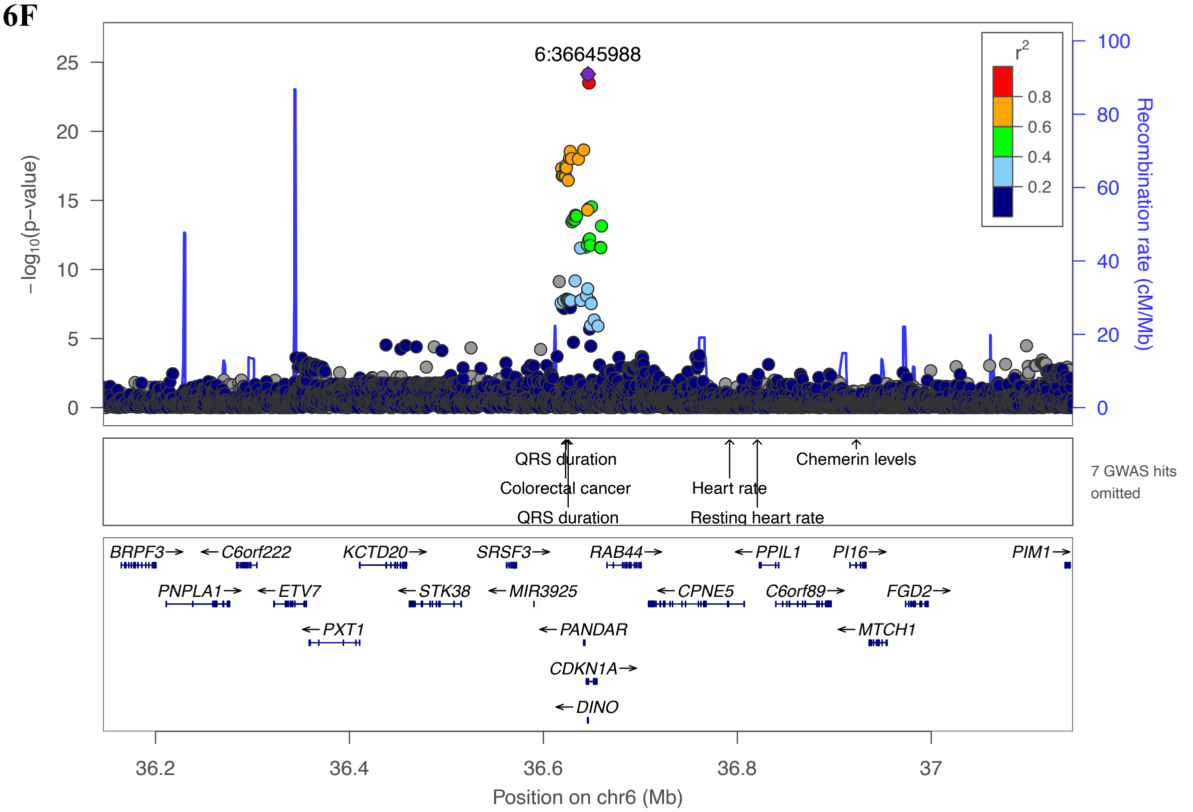
6:36645988
r2
0.8
0.6
0.4
0.2
−log10(p−value)
Recombination rate (cM/Mb)
QRS duration
Colorectal cancer
QRS duration
Heart rate
Resting heart rate
Chemerin levels
7 GWAS hits omitted
BRPF3→
←C6orf222
KCTD20→
SRSF3→
RAB44→
←PPIL1
PI16→
PIM1→
PNPLA1→
←ETV7
←STK38
←MIR3925
←CPNE5
C6orf89→
FGD2→
←PXT1
←PANDAR
←MTCH1
CDKN1A→
←DINO
36.2
36.4
36.6
36.8
37
Position on chr6 (Mb)

6G

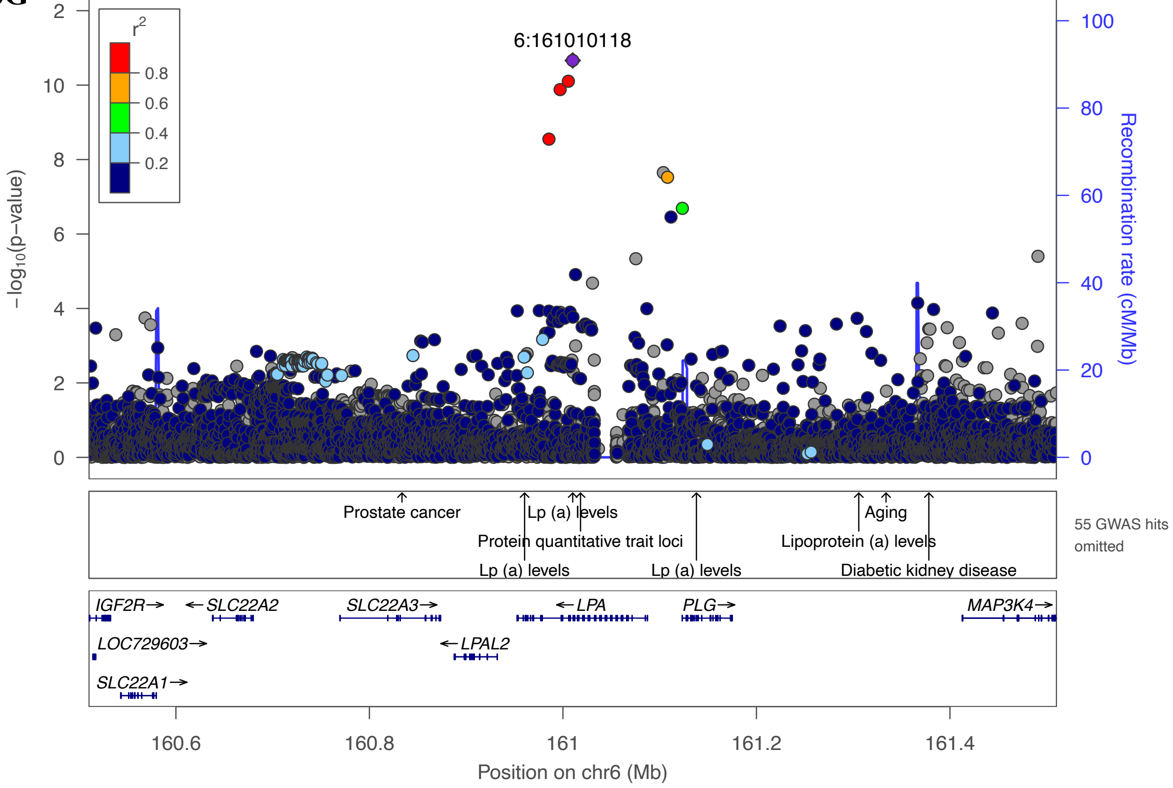

6:161010118
r2
0.8
0.6
0.4
0.2
−log10(p−value)
Recombination rate (cM/Mb)
Prostate cancer
Lp (a) levels
Protein quantitative trait loci
Lp (a) levels
Lp (a) levels
Aging
Lipoprotein (a) levels
Diabetic kidney disease
55 GWAS hits omitted
IGF2R→
←SLC22A2
SLC22A3→
←LPA
PLG→
MAP3K4→
LOC729603→
←LPAL2
SLC22A1→
160.6
160.8
161
161.2
161.4
Position on chr6 (Mb)


6H

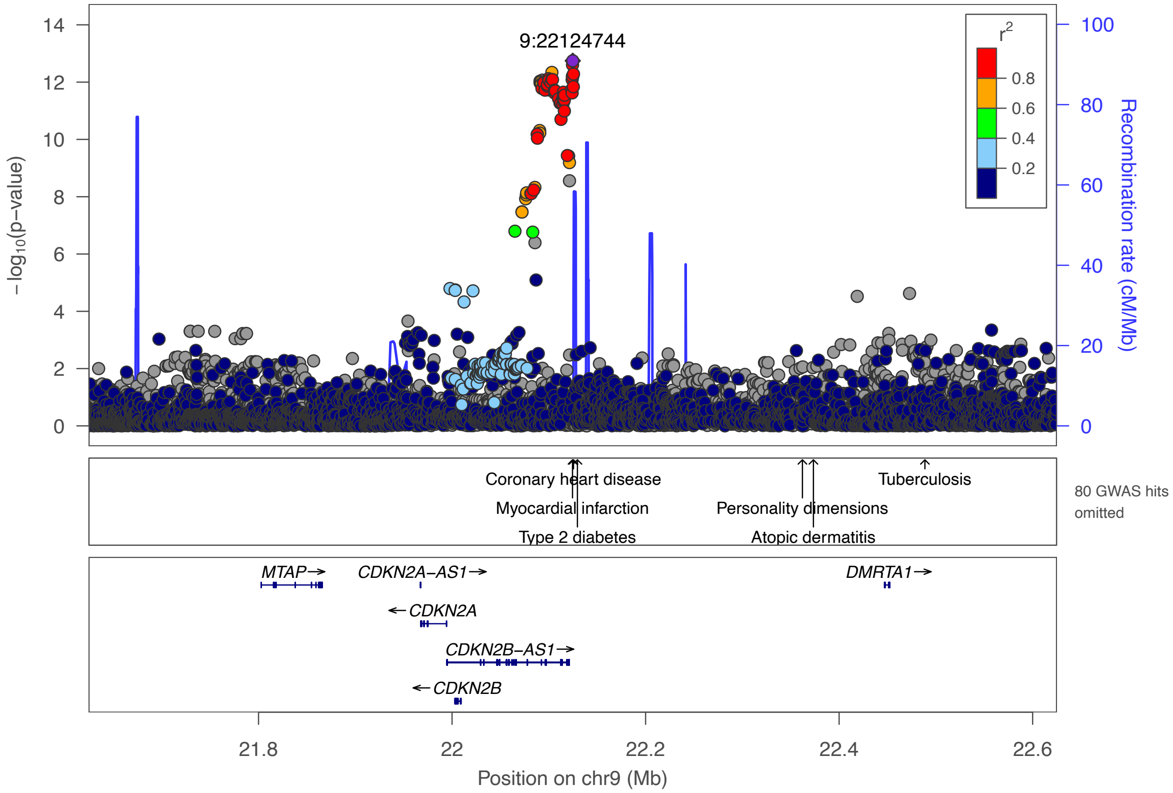

9:22124744
r2
0.8
0.6
0.4
0.2
−log10(p−value)
Recombination rate (cM/Mb)
Coronary heart disease
Myocardial infarction
Type 2 diabetes
Personality dimensions
Atopic dermatitis
Tuberculosis
80 GWAS hits omitted
MTAP→
CDKN2A−AS1→
DMRTA1→
←CDKN2A
CDKN2B−AS1→
←CDKN2B
21.8
22
22.2
22.4
22.6
Position on chr9 (Mb)

6I

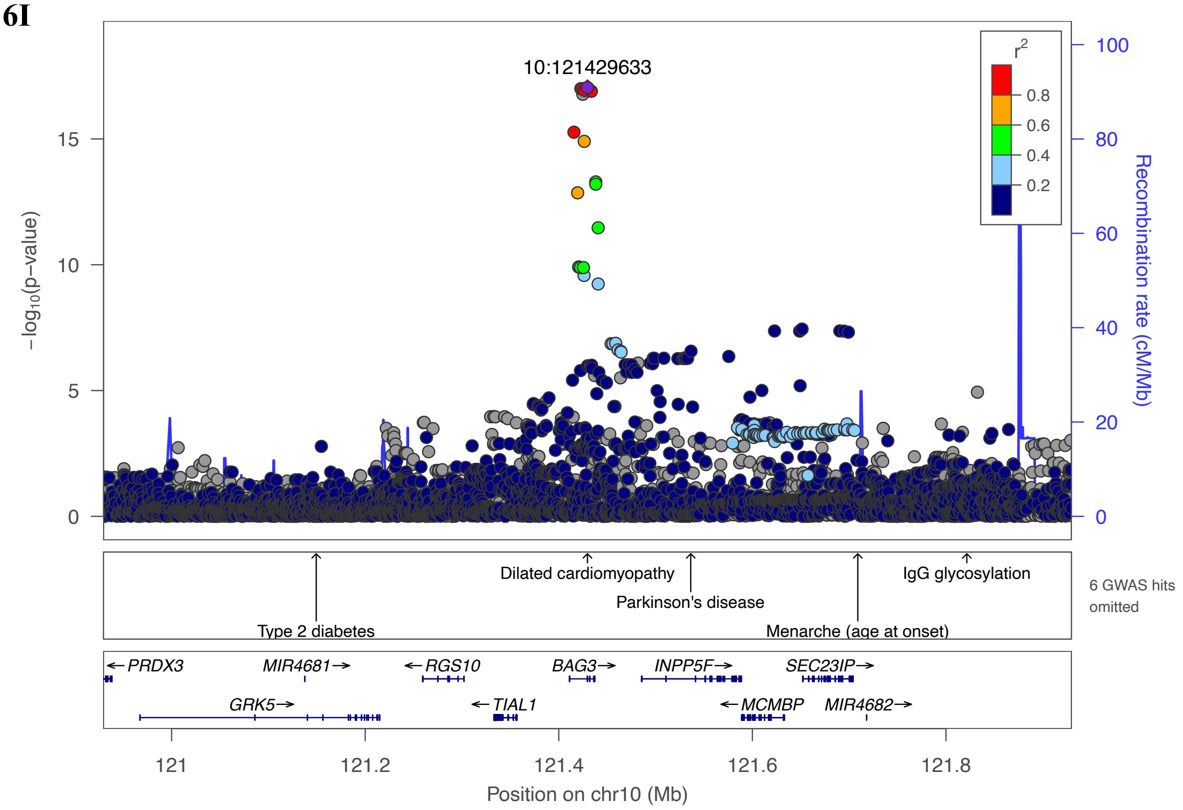
10:121429633
r²
0.8
0.6
0.4
0.2
−log10(p−value)
Recombination rate (cM/Mb)
Dilated cardiomyopathy
Parkinson's disease
IgG glycosylation
Type 2 diabetes
Menarche (age at onset)
6 GWAS hits omitted
PRDX3
MIR4681
RGS10
BAG3
INPP5F
SEC23IP
GRK5
TIAL1
MCMBP
MIR4682
121
121.2
121.4
121.6
121.8
Position on chr10 (Mb)

6J

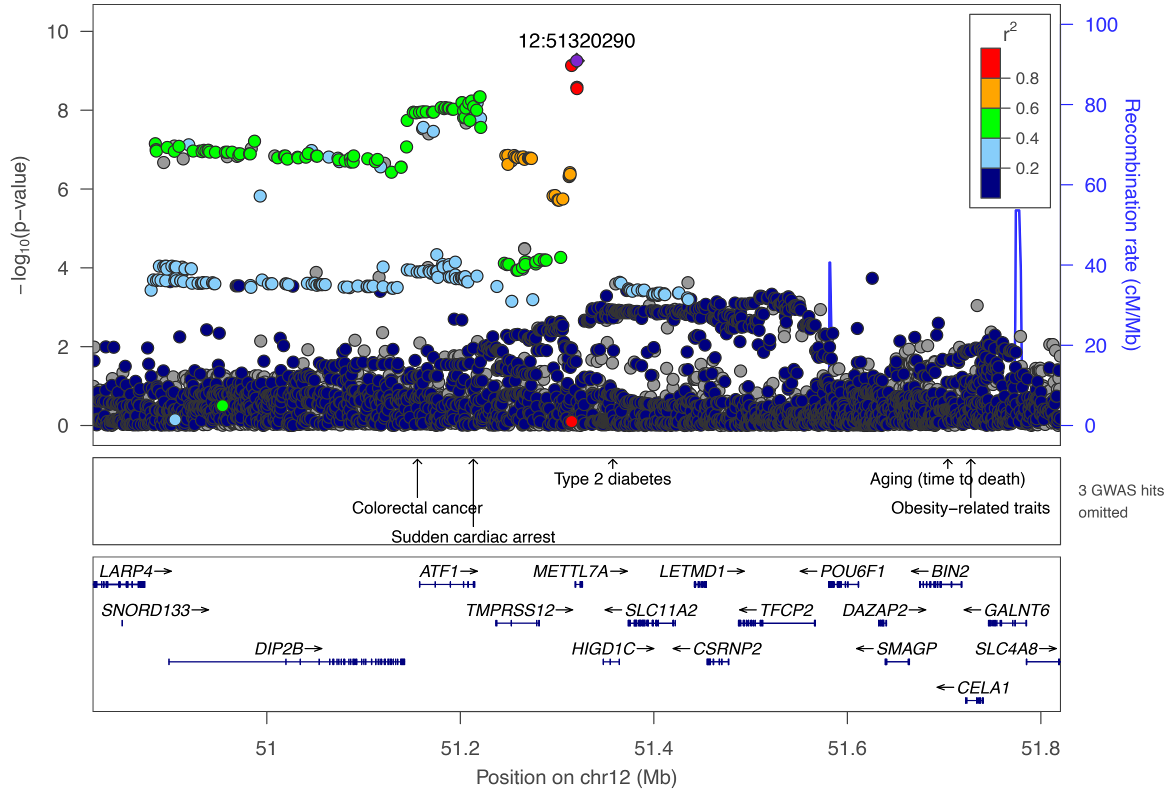
12:51320290
r²
0.8
0.6
0.4
0.2
−log10(p−value)
Recombination rate (cM/Mb)
Colorectal cancer
Sudden cardiac arrest
Type 2 diabetes
Aging (time to death)
Obesity−related traits
3 GWAS hits omitted
LARP4
ATF1
METTL7A
LETMD1
POU6F1
BIN2
SNORD133
TMPRSS12
SLC11A2
TFCP2
DAZAP2
GALNT6
DIP2B
HIGD1C
CSRNP2
SMAGP
SLC4A8
CELA1
51
51.2
51.4
51.6
51.8
Position on chr12 (Mb)

**6K**

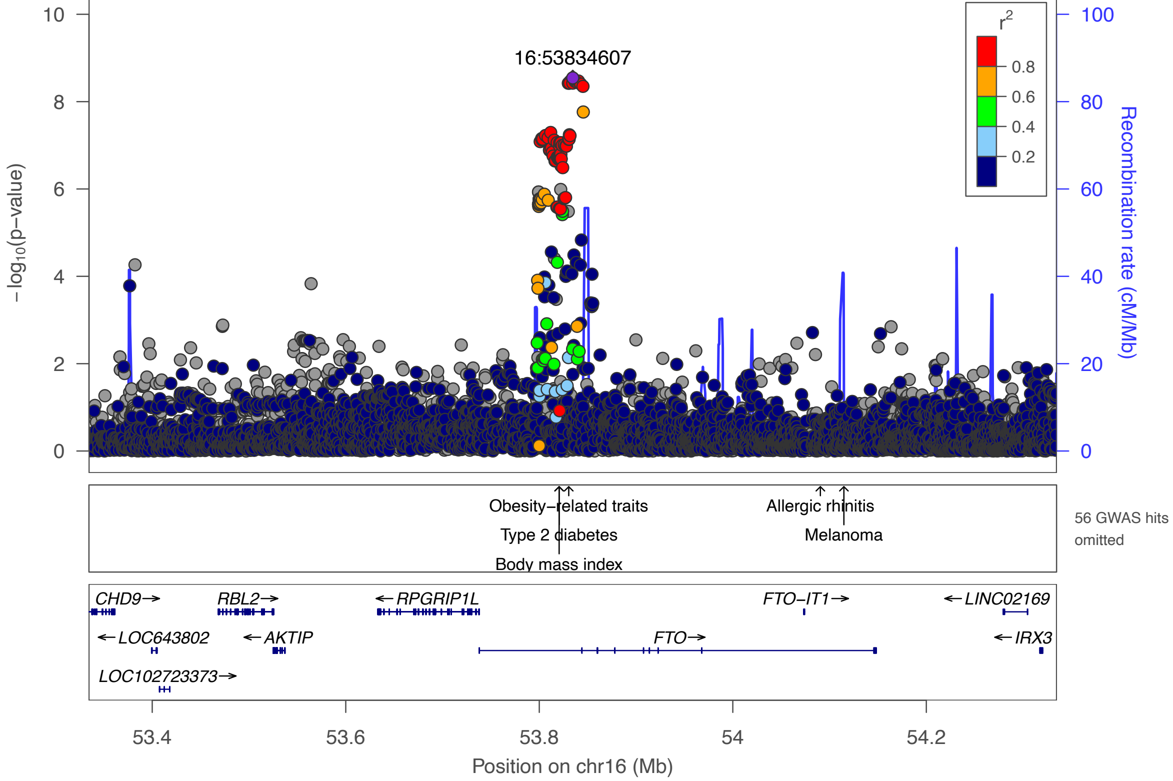

16:53834607
r²
0.8
0.6
0.4
0.2
−log₁₀(p−value)
Recombination rate (cM/Mb)
Obesity−related traits
Type 2 diabetes
Body mass index
Allergic rhinitis
Melanoma
56 GWAS hits omitted
CHD9→
RBL2→
←RPGRIP1L
FTO−IT1→
←LINC02169
←LOC643802
←AKTIP
FTO→
←IRX3
LOC102723373→
53.4
53.6
53.8
54
54.2
Position on chr16 (Mb)


**5L**

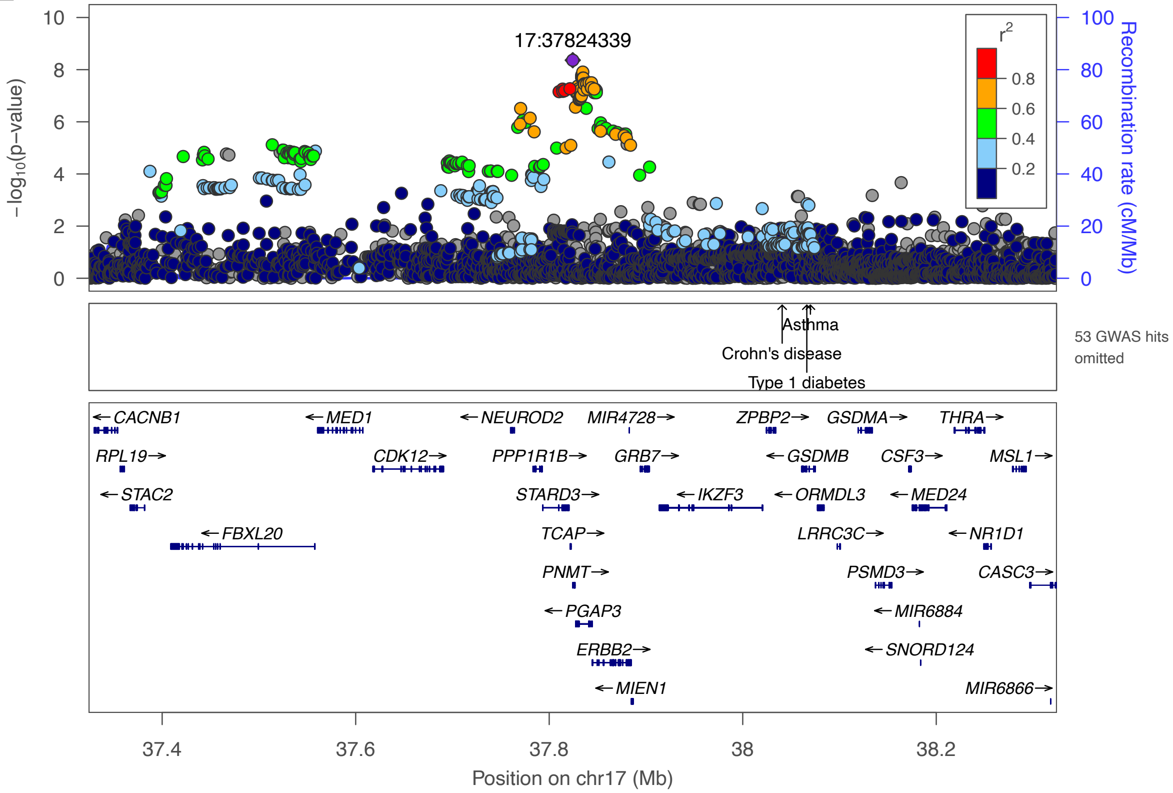

17:37824339
r²
0.8
0.6
0.4
0.2
−log₁₀(p−value)
Recombination rate (cM/Mb)
Asthma
Crohn's disease
Type 1 diabetes
53 GWAS hits omitted
←CACNB1
←MED1
←NEUROD2
MIR4728→
ZPBP2→
GSDMA→
THRA→
RPL19→
CDK12→
PPP1R1B→
GRB7→
←GSDMB
CSF3→
MSL1→
←STAC2
STARD3→
←IKZF3
←ORMDL3
←MED24
←FBXL20
TCAP→
LRRC3C→
←NR1D1
PNMT→
PSMD3→
CASC3→
←PGAP3
←MIR6884
ERBB2→
←SNORD124
←MIEN1
MIR6866→
37.4
37.6
37.8
38
38.2
Position on chr17 (Mb)

6M

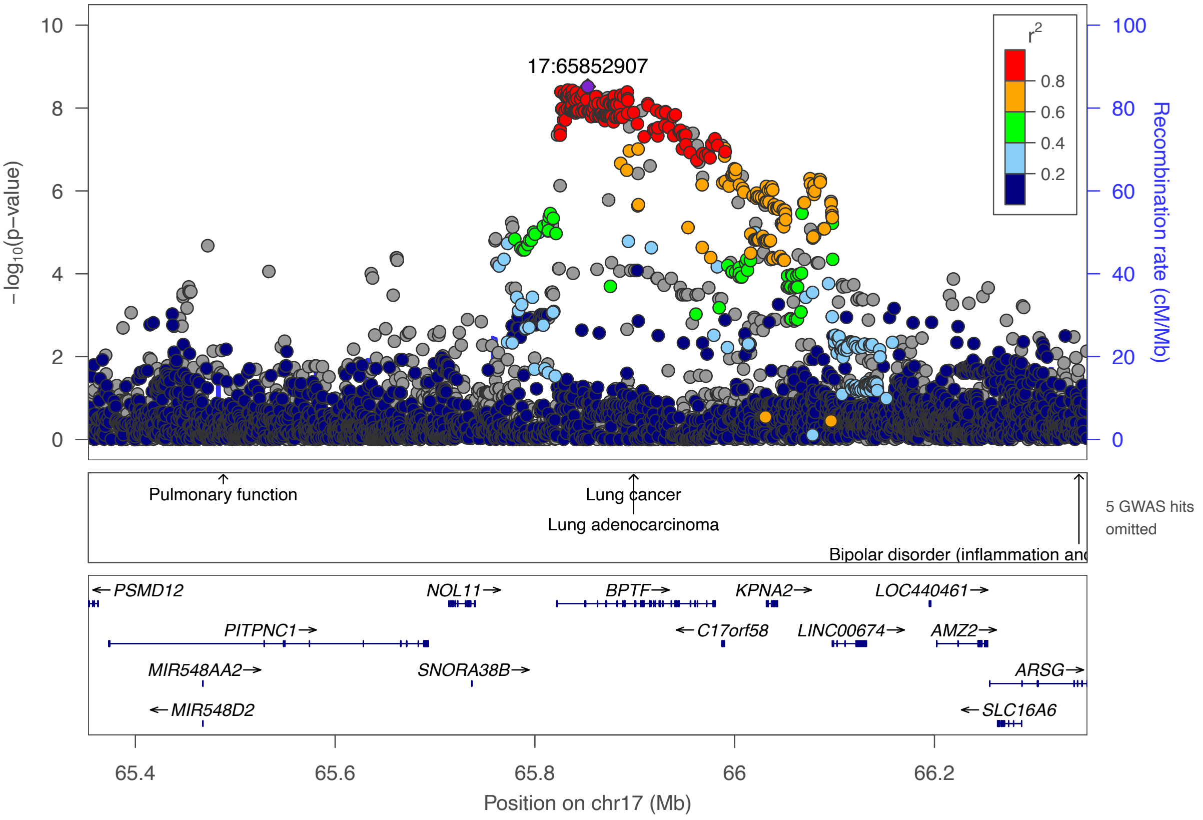

17:65852907
r2
0.8
0.6
0.4
0.2
−log10(p−value)
Recombination rate (cM/Mb)
Pulmonary function
Lung cancer
Lung adenocarcinoma
Bipolar disorder (inflammation and
5 GWAS hits omitted
PSMD12
NOL11
BPTF
KPNA2
LOC440461
PITPNC1
C17orf58
LINC00674
AMZ2
MIR548AA2
SNORA38B
ARSG
MIR548D2
SLC16A6
65.4
65.6
65.8
66
66.2
Position on chr17 (Mb)

**Supplementary Figure 6N**. Genome-wide significant locus associated with HFpEF

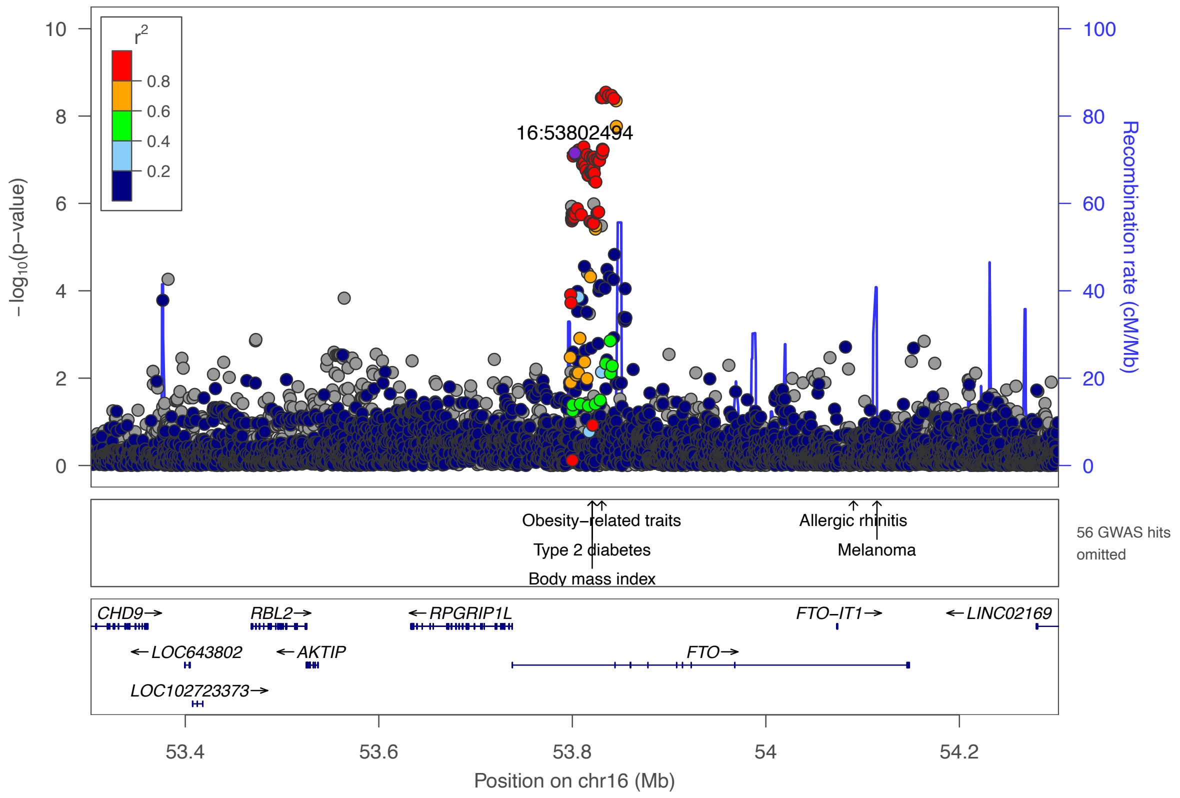

**Supplementary Figure 7.** Genetic associations of HFrEF in the *TMEM43* region.

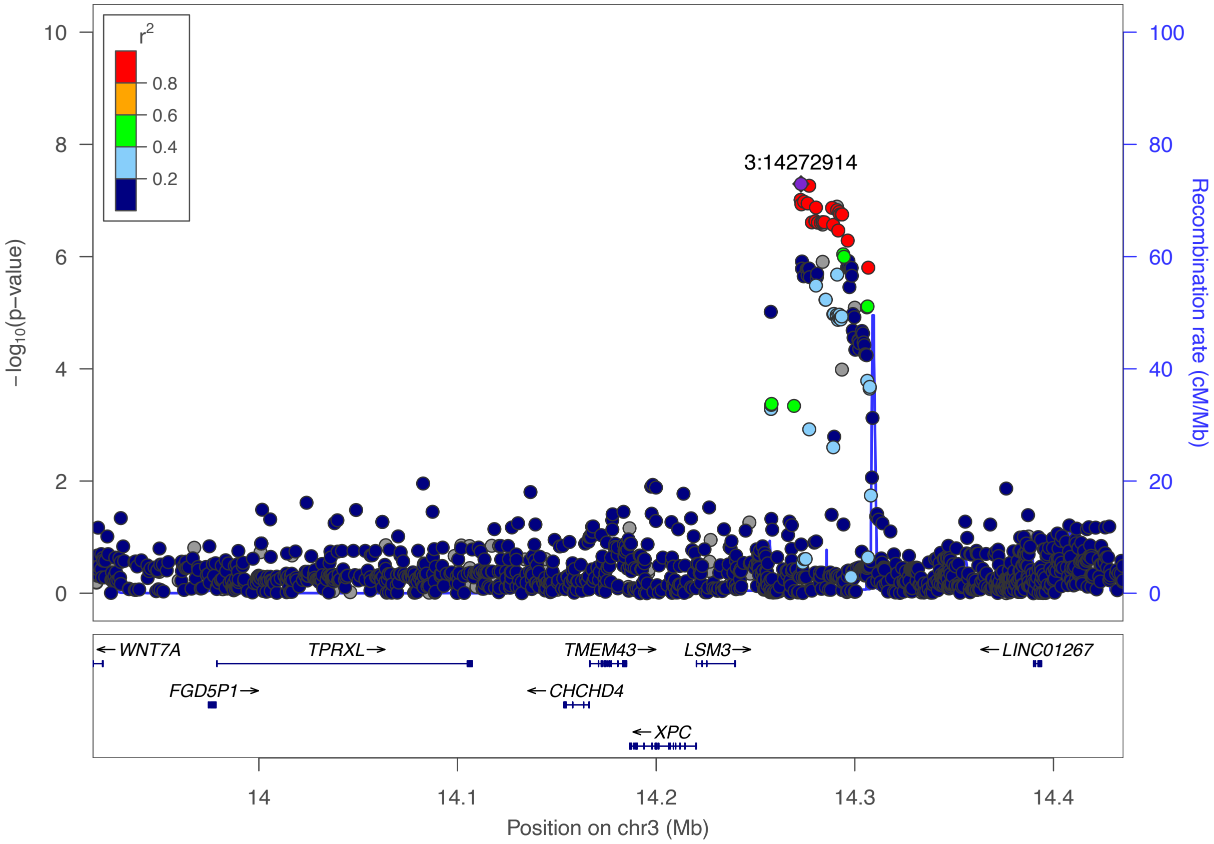

**Consortium Authors: VA Million Veteran Program**

### MVP Executive Committee

- Co-Chair: J. Michael Gaziano, M.D., M.P.H.
  VA Boston Healthcare System, 150 S. Huntington Avenue, Boston, MA 02130
- Co-Chair: Sumitra Muralidhar, Ph.D.
  US Department of Veterans Affairs, 810 Vermont Avenue NW, Washington, DC 20420
- Rachel Ramoni, D.M.D., Sc.D., Chief VA Research and Development Officer
  US Department of Veterans Affairs, 810 Vermont Avenue NW, Washington, DC 20420
- Jean Beckham, Ph.D.
  Durham VA Medical Center, 508 Fulton Street, Durham, NC 27705
- Kyong-Mi Chang, M.D.
  Philadelphia VA Medical Center, 3900 Woodland Avenue, Philadelphia, PA 19104
- Christopher J. O'Donnell, M.D., M.P.H.
  VA Boston Healthcare System, 150 S. Huntington Avenue, Boston, MA 02130
- Philip S. Tsao, Ph.D.
  VA Palo Alto Health Care System, 3801 Miranda Avenue, Palo Alto, CA 94304
- James Breeling, M.D., Ex-Officio
  US Department of Veterans Affairs, 810 Vermont Avenue NW, Washington, DC 20420
- Grant Huang, Ph.D., Ex-Officio
  US Department of Veterans Affairs, 810 Vermont Avenue NW, Washington, DC 20420
- JP Casas Romero, M.D., Ph.D., Ex-Officio
  VA Boston Healthcare System, 150 S. Huntington Avenue, Boston, MA 02130

### MVP Program Office

- Sumitra Muralidhar, Ph.D.
  US Department of Veterans Affairs, 810 Vermont Avenue NW, Washington, DC 20420
- Jennifer Moser, Ph.D.
  US Department of Veterans Affairs, 810 Vermont Avenue NW, Washington, DC 20420

### MVP Recruitment/Enrollment

- Recruitment/Enrollment Director/Deputy Director, Boston – Stacey B. Whitbourne, Ph.D.; Jessica V. Brewer, M.P.H.
  VA Boston Healthcare System, 150 S. Huntington Avenue, Boston, MA 02130
- MVP Coordinating Centers
  - Clinical Epidemiology Research Center (CERC), West Haven – Mihaela Aslan, Ph.D.
    West Haven VA Medical Center, 950 Campbell Avenue, West Haven, CT 06516
  - Cooperative Studies Program Clinical Research Pharmacy Coordinating Center, Albuquerque – Todd Connor, Pharm.D.; Dean P. Argyres, B.S., M.S.

New Mexico VA Health Care System, 1501 San Pedro Drive SE, Albuquerque, NM 87108

- Genomics Coordinating Center, Palo Alto – Philip S. Tsao, Ph.D.
  VA Palo Alto Health Care System, 3801 Miranda Avenue, Palo Alto, CA 94304
- MVP Boston Coordinating Center, Boston - J. Michael Gaziano, M.D., M.P.H.
  VA Boston Healthcare System, 150 S. Huntington Avenue, Boston, MA 02130
- MVP Information Center, Canandaigua – Brady Stephens, M.S.
  Canandaigua VA Medical Center, 400 Fort Hill Avenue, Canandaigua, NY 14424

- VA Central Biorepository, Boston – Mary T. Brophy M.D., M.P.H.; Donald E. Humphries, Ph.D.; Luis E. Selva, Ph.D.
  VA Boston Healthcare System, 150 S. Huntington Avenue, Boston, MA 02130
- MVP Informatics, Boston – Nhan Do, M.D.; Shahpoor Shayan
  VA Boston Healthcare System, 150 S. Huntington Avenue, Boston, MA 02130
- MVP Data Operations/Analytics, Boston – Kelly Cho, Ph.D.
  VA Boston Healthcare System, 150 S. Huntington Avenue, Boston, MA 02130

**MVP Science**

- Science Operations – Christopher J. O'Donnell, M.D., M.P.H.
  VA Boston Healthcare System, 150 S. Huntington Avenue, Boston, MA 02130
- Genomics Core - Christopher J. O'Donnell, M.D., M.P.H.; Saiju Pyarajan Ph.D.
  VA Boston Healthcare System, 150 S. Huntington Avenue, Boston, MA 02130
  Philip S. Tsao, Ph.D.
  VA Palo Alto Health Care System, 3801 Miranda Avenue, Palo Alto, CA 94304
- Phenomics Core- Kelly Cho, M.P.H, Ph.D.
  VA Boston Healthcare System, 150 S. Huntington Avenue, Boston, MA 02130
- Data and Computational Sciences – Saiju Pyarajan, Ph.D.
  VA Boston Healthcare System, 150 S. Huntington Avenue, Boston, MA 02130
- Statistical Genetics – Elizabeth Hauser, Ph.D.
  Durham VA Medical Center, 508 Fulton Street, Durham, NC 27705
  Yan V. Sun, Ph.D.
  Atlanta VA Medical Center, 1670 Clairmont Road, Decatur, GA 30033
  Hongyu Zhao, Ph.D.
  West Haven VA Medical Center, 950 Campbell Avenue, West Haven, CT 06516

**Current MVP Local Site Investigators**

- Atlanta VA Medical Center (Peter Wilson, M.D.)
  1670 Clairmont Road, Decatur, GA 30033
- Bay Pines VA Healthcare System (Rachel McArdle, Ph.D.)
  10,000 Bay Pines Blvd Bay Pines, FL 33744
- Birmingham VA Medical Center (Louis Dellitalia, M.D.)
  700 S. 19th Street, Birmingham AL 35233
- Central Western Massachusetts Healthcare System (Kristin Mattocks, Ph.D., M.P.H.)

421 North Main Street, Leeds, MA 01053

- Cincinnati VA Medical Center (John Harley, M.D., Ph.D.)
  3200 Vine Street, Cincinnati, OH 45220
- Clement J. Zablocki VA Medical Center (Jeffrey Whittle, M.D., M.P.H.)
  5000 West National Avenue, Milwaukee, WI 53295
- VA Northeast Ohio Healthcare System (Frank Jacono, M.D.)
  10701 East Boulevard, Cleveland, OH 44106
- Durham VA Medical Center (Jean Beckham, Ph.D.)
  508 Fulton Street, Durham, NC 27705
- Edith Nourse Rogers Memorial Veterans Hospital (John Wells., Ph.D.)
  200 Springs Road, Bedford, MA 01730
- Edward Hines, Jr. VA Medical Center (Salvador Gutierrez, M.D.)
  5000 South 5th Avenue, Hines, IL 60141
- Veterans Health Care System of the Ozarks (Gretchen Gibson, D.D.S., M.P.H.)
  1100 North College Avenue, Fayetteville, AR 72703
- Fargo VA Health Care System (Kimberly Hammer, Ph.D.)
  2101 N. Elm, Fargo, ND 58102
- VA Health Care Upstate New York (Laurence Kaminsky, Ph.D.)
  113 Holland Avenue, Albany, NY 12208
- New Mexico VA Health Care System (Gerardo Villareal, M.D.)
  1501 San Pedro Drive, S.E. Albuquerque, NM 87108
- VA Boston Healthcare System (Scott Kinlay, M.B.B.S., Ph.D.)
  150 S. Huntington Avenue, Boston, MA 02130
- VA Western New York Healthcare System (Junzhe Xu, M.D.)
  3495 Bailey Avenue, Buffalo, NY 14215-1199
- Ralph H. Johnson VA Medical Center (Mark Hamner, M.D.)
  109 Bee Street, Mental Health Research, Charleston, SC 29401
- Columbia VA Health Care System (Roy Mathew, M.D.)
  6439 Garners Ferry Road, Columbia, SC 29209
- VA North Texas Health Care System (Sujata Bhushan, M.D.)
  4500 S. Lancaster Road, Dallas, TX 75216
- Hampton VA Medical Center (Pran Iruvanti, D.O., Ph.D.)
  100 Emancipation Drive, Hampton, VA 23667
- Richmond VA Medical Center (Michael Godschalk, M.D.)
  1201 Broad Rock Blvd., Richmond, VA 23249
- Iowa City VA Health Care System (Zuhair Ballas, M.D.)
  601 Highway 6 West, Iowa City, IA 52246-2208
- Eastern Oklahoma VA Health Care System (Douglas Ivins, M.D.)
  1011 Honor Heights Drive, Muskogee, OK 74401
- James A. Haley Veterans’ Hospital (Stephen Mastorides, M.D.)
  13000 Bruce B. Downs Blvd, Tampa, FL 33612

- James H. Quillen VA Medical Center (Jonathan Moorman, M.D., Ph.D.)
  Corner of Lamont & Veterans Way, Mountain Home, TN 37684
- John D. Dingell VA Medical Center (Saib Gappy, M.D.)
  4646 John R Street, Detroit, MI 48201
- Louisville VA Medical Center (Jon Klein, M.D., Ph.D.)
  800 Zorn Avenue, Louisville, KY 40206
- Manchester VA Medical Center (Nora Ratcliffe, M.D.)
  718 Smyth Road, Manchester, NH 03104
- Miami VA Health Care System (Hermes Florez, M.D., Ph.D.)
  1201 NW 16th Street, 11 GRC, Miami FL 33125
- Michael E. DeBakey VA Medical Center (Olaoluwa Okusaga, M.D.)
  2002 Holcombe Blvd, Houston, TX 77030
- Minneapolis VA Health Care System (Maureen Murdoch, M.D., M.P.H.)
  One Veterans Drive, Minneapolis, MN 55417
- N. FL/S. GA Veterans Health System (Peruvemba Sriram, M.D.)
  1601 SW Archer Road, Gainesville, FL 32608
- Northport VA Medical Center (Shing Shing Yeh, Ph.D., M.D.)
  79 Middleville Road, Northport, NY 11768
- Overton Brooks VA Medical Center (Neeraj Tandon, M.D.)
  510 East Stoner Ave, Shreveport, LA 71101
- Philadelphia VA Medical Center (Darshana Jhala, M.D.)
  3900 Woodland Avenue, Philadelphia, PA 19104
- Phoenix VA Health Care System (Samuel Aguayo, M.D.)
  650 E. Indian School Road, Phoenix, AZ 85012
- Portland VA Medical Center (David Cohen, M.D.)
  3710 SW U.S. Veterans Hospital Road, Portland, OR 97239
- Providence VA Medical Center (Satish Sharma, M.D.)
  830 Chalkstone Avenue, Providence, RI 02908
- Richard Roudebush VA Medical Center (Suthat Liangpunsakul, M.D., M.P.H.)
  1481 West 10th Street, Indianapolis, IN 46202
- Salem VA Medical Center (Kris Ann Oursler, M.D.)
  1970 Roanoke Blvd, Salem, VA 24153
- San Francisco VA Health Care System (Mary Whooley, M.D.)
  4150 Clement Street, San Francisco, CA 94121
- South Texas Veterans Health Care System (Sunil Ahuja, M.D.)
  7400 Merton Minter Boulevard, San Antonio, TX 78229
- Southeast Louisiana Veterans Health Care System (Joseph Constans, Ph.D.)
  2400 Canal Street, New Orleans, LA 70119
- Southern Arizona VA Health Care System (Paul Meyer, M.D., Ph.D.)
  3601 S 6th Avenue, Tucson, AZ 85723
- Sioux Falls VA Health Care System (Jennifer Greco, M.D.)

2501 W 22nd Street, Sioux Falls, SD 57105
- St. Louis VA Health Care System (Michael Rauchman, M.D.)
  915 North Grand Blvd, St. Louis, MO 63106
- Syracuse VA Medical Center (Richard Servatius, Ph.D.)
  800 Irving Avenue, Syracuse, NY 13210
- VA Eastern Kansas Health Care System (Melinda Gaddy, Ph.D.)
  4101 S 4th Street Trafficway, Leavenworth, KS 66048
- VA Greater Los Angeles Health Care System (Agnes Wallbom, M.D., M.S.)
  11301 Wilshire Blvd, Los Angeles, CA 90073
- VA Long Beach Healthcare System (Timothy Morgan, M.D.)
  5901 East 7th Street Long Beach, CA 90822
- VA Maine Healthcare System (Todd Stapley, D.O.)
  1 VA Center, Augusta, ME 04330
- VA New York Harbor Healthcare System (Scott Sherman, M.D., M.P.H.)
  423 East 23rd Street, New York, NY 10010
- VA Pacific Islands Health Care System (George Ross, M.D.)
  459 Patterson Rd, Honolulu, HI 96819
- VA Palo Alto Health Care System (Philip Tsao, Ph.D.)
  3801 Miranda Avenue, Palo Alto, CA 94304-1290
- VA Pittsburgh Health Care System (Patrick Strollo, Jr., M.D.)
  University Drive, Pittsburgh, PA 15240
- VA Puget Sound Health Care System (Edward Boyko, M.D.)
  1660 S. Columbian Way, Seattle, WA 98108-1597
- VA Salt Lake City Health Care System (Laurence Meyer, M.D., Ph.D.)
  500 Foothill Drive, Salt Lake City, UT 84148
- VA San Diego Healthcare System (Samir Gupta, M.D., M.S.C.S.)
  3350 La Jolla Village Drive, San Diego, CA 92161
- VA Sierra Nevada Health Care System (Mostaqul Huq, Pharm.D., Ph.D.)
  975 Kirman Avenue, Reno, NV 89502
- VA Southern Nevada Healthcare System (Joseph Fayad, M.D.)
  6900 North Pecos Road, North Las Vegas, NV 89086
- VA Tennessee Valley Healthcare System (Adriana Hung, M.D., M.P.H.)
  1310 24th Avenue, South Nashville, TN 37212
- Washington DC VA Medical Center (Jack Lichy, M.D., Ph.D.)
  50 Irving St, Washington, D. C. 20422
- W.G. (Bill) Hefner VA Medical Center (Robin Hurley, M.D.)
  1601 Brenner Ave, Salisbury, NC 28144
- White River Junction VA Medical Center (Brooks Robey, M.D.)
  163 Veterans Drive, White River Junction, VT 05009
- William S. Middleton Memorial Veterans Hospital (Robert Striker, M.D., Ph.D.)
  2500 Overlook Terrace, Madison, WI 53705
